# Evaluation of the Contribution of Natural Selection to Greater Cardiometabolic Disease Risk in South Asian Populations

**DOI:** 10.64898/2026.05.15.26353234

**Authors:** Daniel Searby, Amanda Chong, George Davey-Smith, Nish Chaturvedi, Daniel Lawson, Gibran Hemani

## Abstract

A greater genetic susceptibility has been proposed as an explanation of the greater rates of cardiovascular and metabolic disease in South Asian relative to European populations. We first demonstrate that after accounting for technical artefacts the genetic effects for related traits are largely consistent between ancestral groups, which downplays the role of GxG or GxE interactions driving differential prevalence. If higher genetic susceptibility in South Asians is due to selective pressures acting through adiposity-related traits in the evolutionary past, signatures of selection should be evident at loci associated with cardiometabolic disease and other causally related traits (e.g. fat distribution). We tested for enrichment of several selection statistics (F_ST,_ XP-EHH and XP-nSL) at loci associated with a range of traits related to cardiometabolic disease, in comparison to a null distribution of linkage disequilibrium (LD) score and minor allele frequency (MAF) matched SNPs. Loci associated with a subset of these traits (Type 2 diabetes mellitus, trunk fat percentage, body fat percentage and trunk fat mass) exhibited enrichment for F_ST_, consistent with a moderate adaptive explanation for their cross-population differentiation. In contrast, none of the studied traits were enriched for haplotype-based statistics, indicative that cross population genetic divergence is unlikely to have been driven by recent selective sweeps but has rather likely arisen from either ancient selection or recent polygenic selection acting on standing variation.

## Introduction

South Asian diaspora populations within western countries generally exhibit far greater risk for a range of cardiometabolic diseases compared to European populations within the same countries, with the risk of cardiovascular disease and Type 2 diabetes mellitus (T2D) being 1.5-2 and 2-4 fold [1,2] greater respectively than Europeans. Additionally, South Asians generally develop T2D and cardiovascular disease earlier in life and at a lower body mass index (BMI) compared to Europeans, [3, 4] and convert from a prediabetic to diabetic state far more rapidly [5]. Given the striking consistency of these diagnostic and prognostic patterns despite the heterogeneity in exposure to environmental risk factors (e.g. cultural practices, socioeconomic and occupational status) amongst South Asian populations, it has been proposed that this increased risk may arise in part due to a greater genetic susceptibility [6]. This claim is also supported by evidence from intergenerational migration studies, in which it has been observed that excess diabetes risk persists in second generation migrants as well as those of mixed ancestry, and cannot be fully explained by known environmental risk factors [7].

Potential mechanisms by which a greater genetic susceptibility could manifest are evident in observed differences in metabolic and adipose biology between South Asian and European populations. From early life and throughout adulthood, South Asians in general have greater visceral[8] and hepatic fat deposition[9]],increased insulin resistance [10] and impaired insulin secretion capacity[11], as well as lower lean mass [12]. Studies seeking a role of a greater genetic susceptibility for cardiometabolic disease in South Asian populations, comparing either frequency or effect sizes of known risk alleles, have generally found little evidence in support of an important contribution of known loci to either greater T2D risk [13] or greater waist-hip ratio [14]. Nonetheless, it remains plausible that a greater genetic susceptibility may arise through variants that would go undiscovered given current sample sizes in South Asian specific genome-wide association studies (GWAS) [15]. Additionally, population specific LD patterns diminish the extent to which loci ascertained from predominantly European discovery GWAS sufficiently tag causal variants in non-European populations [16].

Under the assumption that the greater susceptibility does have a genetic basis, several evolutionary hypotheses have been proposed to explain the selective pressures that may have shaped the genetic architecture of fat distribution traits in South Asian populations [17–24]. Whilst until recently empirical evaluation of these hypotheses was not feasible, it is now possible to leverage the abundance of genetic associations for complex traits to evaluate evidence of the role of natural selection in shaping their genetic architecture [25]. Application of these methods makes it possible to infer the evolutionary history of fat distribution traits and cardiometabolic disease across global populations and its potential contribution to present day disease risk.

Detection of polygenic adaptation is a considerable challenge, especially when considering evidence of differential selection across ancestry groups, mainly due to the sparsity of genetic data available for non-European populations, in addition to observed biases arising from stratification when applying polygenic scores (PS) or trait SNP associations ascertained from predominantly European discovery datasets [26]. In particular, studies testing for overdispersion or directional changes in the frequency of trait associated variants have been demonstrated to suffer from pervasive bias arising from residual population stratification with the estimated magnitude of selection being highly sensitive to the choice of discovery cohort [27]. Whilst recent studies limit the scale of variation in genetic effects between populations [28], phenotypic effects of alleles may not be consistent between populations due to gene-gene interaction (epistasis), gene environment interaction (GxE), and associations differ due to linkage disequilibrium (LD) patterns between ascertained and causal variants across populations [29]. Differential demographic history, admixture, or population structure can all obscure or overaccentuate signatures of selection at trait associated loci [30–33].

With these caveats in mind, more robust approaches to quantifying the role of natural selection in shaping cross population genetic differentiation in complex traits have been devised. Guo et al [34] present an approach to quantifying the extent to which cross population genetic differentiation of complex traits is attributable to natural selection by comparing the mean fixation index (F_ST_) of trait associated SNPs to a background composed of LD score and minor allele frequency (MAF) matched SNPs, with the underlying assumption being that under genetic drift, the mean F_ST_ of trait associated SNPs should not depart significantly from the null distribution. This approach has the advantage of not requiring direct use of effect sizes or directions at trait associated loci (since F_ST_ is an unsigned statistic), theoretically minimising bias arising from residual stratification.

We applied this method to several traits relevant to cardiometabolic disease and body fat distribution; Trunk Fat Mass (TFM), Trunk Fat Percentage (TFP), Body Fat Percentage (BFP), BMI, Appendicular Lean Mass (ALM) and T2D, as well as two positive controls for selection, height and black hair colour, in order to assess evidence for cross population genetic differentiation of these traits between South Asian and European populations having been contributed by natural selection (see Methods – Data Sources).

Height was included as a positive control given its status as one of the most extensively studied complex traits in human evolutionary genetics, for which signatures of selection have repeatedly been reported [35–39]. Recent studies have reappraised the findings of strong directional selection for height, finding many of the original results to be affected by population stratification and other confounders [40, 41]. Nonetheless, results still largely indicate that (although attenuated from earlier estimates) directional selection has acted on height, and it therefore remains a valid comparison trait to fat distribution and cardiometabolic disease traits of similar polygenicity.

Black hair colour was included as a contrasting positive control due to the unambiguous phenotypic differentiation between European and South Asian populations for this trait, and the extensive evidence for both European-specific selection at pigmentation-associated loci [37, 42]. Pigmentation traits have repeatedly shown enrichment for both F_ST_ and haplotype-based selection statistics, reflecting the action of relatively strong and geographically structured selection pressures [43]. Accordingly, enrichment of evolutionary statistics at black hair–associated loci provide a more robust validation of the enrichment framework applied here, and contextualise the magnitude of selection that may be acting on other studied traits.

A major limitation of the original study that applied this method was that all trait SNP associations were ascertained from European discovery datasets, due to a lack of larger sample sizes for GWAS’s for the traits under study from alternative populations at the time of publication. Therefore, in the present study we made use of recently available large meta-analyses combining both South Asian and European discovery GWAS, theoretically minimising bias arising from differences in effect size or frequency of trait associated SNPs between populations. We additionally extended the F_ST_ enrichment test by also integrating two statistics that quantify evidence for cross population selection based on leveraging the reduction in haplotype diversity characteristic of a selective sweep around selected loci – cross population extended haplotype homozygosity (XP-EHH)[44] and cross population number of segregating lengths by site (XP-nSL)[45]. Whilst the F_ST_ statistic is generally interpreted as allowing detection of selection on the timescale of very recently to many tens of thousands of years (>50000-75000), both of these statistics are better powered to detect selective sweeps that occurred over the last 10-30 thousand years [46]. XP-EHH is better powered to detect classical ‘hard’ sweeps in which a newly arising allele rapidly rises from low to high frequency, typically on a single haplotypic background carrying the original mutation. Conversely, XP-nSL is better powered to detect ‘soft’ sweeps on standing variation, in which the favoured allele may rise in frequency on multiple haplotypic backgrounds simultaneously, as well as incomplete sweeps [45]. Both methods are best powered when the sweep is near (>80%) fixation in either the target or reference population [47].

**Figure 1:** Schematic overview of the study design Yellow: input data, Blue: processing steps, Purple: analysis steps

## Results

### Consistency of adiposity related genetic effects between European and South Asian populations

We first aimed to establish if genetic influences on height, black hair colour and adiposity related traits were consistent across ancestral groups. To select variants, we performed cross-ancestry fixed effects meta-analysis which prioritises variant exhibiting consistent effects between populations and diminishes the likelihood that chosen variants represent ancestry-specific LD tagging. To confirm the consistency of effect sizes for sentinel variants chosen from the fixed effect meta-analysis between populations, we then implemented a heterogeneity test that generates a per variant Cochran’s Q statistic, based on its effect size and standard error in the two populations. After adjusting for the false discovery rate and multiple tests, none of the variants associated with any of the fat distribution traits or T2D were found to be significantly heterogenous, with the exception of one BMI associated SNP (rs1229984), (heterogeneity p value = 4.12×10^−5^, South Asian beta: −0.076 ± 0.033, European beta: 0.065 ± 0.009). This variant is located within the *ADH1B* alcohol dehydrogenase gene and is strongly associated with alcohol intake [48]. Therefore, to assess whether the observed cross-ancestry heterogeneity could be explained by cultural patterning of alcohol consumption, we used UK Biobank data accessed via the UK Biobank Research Analysis Platform to fit a regression model including a genotype–ancestry interaction (BMI ∼ genotype × ancestry), as well as a model additionally including a genotype–alcohol consumption interaction term (BMI ∼ genotype × ancestry + genotype × alcohol consumption), in both cases restricting analyses to participants with available alcohol consumption data. After inclusion of the genotype–alcohol consumption interaction term, the ancestry-specific genotype effect (South Asian interaction) was modestly attenuated (from −1.35 to −1.29) but remained highly associated (p = 2.4 × 10⁻⁴). Consistent with strong alcohol-dependent genetic effects, a nested model comparison showed that inclusion of the genotype–alcohol interaction term substantially improved model fit (ΔRSS = 1.8 × 10⁵, F = 4006, p < 10⁻¹⁶). Nonetheless, we cannot conclude that between ancestry heterogeneity is accounted for by alcohol consumption alone.

In contrast to the fat distribution traits and T2D, a total of 7 variants associated with height and 9 associated with black hair colour were found to have heterogenous effects in the two populations after adjusting for multiple tests (Supplementary Tables 4 and 5). Supplementary Figures 49-56 visualise effect estimates for the clumped SNPs in both ancestries, with those that were heterogenous after adjusting for multiple tests highlighted.

Treating the European GWAS as the discovery dataset and the South Asian GWAS as the replication dataset, we then compared the observed replication rate (sign concordance and nominal p-value replication) of sentinel variants from the fixed-effects meta-analysis with the expected rate assuming normally distributed effect estimates.

Replication was generally high for adiposity-related traits (TFM, TFP, BFP, BMI, ALM) as well as for T2D, with observed rates typically in the range of ∼80–90% for both sign concordance and p-value replication. Height also showed strong replication, with around 90% sign concordance and nearly 90% p-value replication. By contrast, replication was weaker for black hair colour, which showed reduced sign concordance (∼75%) and substantially lower p-value replication (∼47%) (Supplementary Table 6).

To further assess the consistency of genetic effects between populations, we calculated the correlation of effect size estimates (beta coefficients) between the European discovery GWAS and the South Asian replication GWAS for each trait. Correlations were generally moderate to high for most traits, reflecting good concordance in the direction and magnitude of genetic effects. Height showed the strongest correlation (r ≈ 0.69), consistent with its high replication rates. Adiposity-related traits including ALM (r ≈ 0.54), BFP (r ≈ 0.44), TFP (r ≈ 0.42), and BMI (r ≈ 0.32) exhibited moderate correlations, while T2D also showed a moderate correlation (r ≈ 0.59). TFM (r ≈ 0.27) and black hair colour (r ≈ 0.27) showed weaker correlations, consistent with their lower replication rates. Overall, these results indicate that effect sizes are largely concordant across populations for most traits, particularly for height and adiposity-related phenotypes.

The low rate of p-value replication for black hair colour may be due to allele frequency differences between the two populations, or potentially epistatic effects, which have been documented in the case of hair colour and other pigmentation traits [49].

Additional factors such as low minor allele frequency in the replication population, measurement heterogeneity, gene–environment interactions, and overestimation of effect sizes in the discovery GWAS may also contribute to reduced replication. Given the high rate of replication for all fat distribution traits, T2D and height there is little evidence for widespread GxE or GxG interaction. We are therefore satisfied that the selected sentinel variants are appropriate for examining signatures of selection at the trait level, given that any selective pressure mediated through the traits of interest should be comparable in the two populations given the concordance of effect sizes. An important caveat regarding our inference of consistency of genetic effects is that the majority of South Asian participants included in the input GWAS utilised were part of diaspora communities resident in western countries, and therefore we are limited in our ability to make inference about the potential for GxE interaction in South Asia itself.

More pertinently, it is ultimately impossible to confirm the consistency of genetic effects across evolutionary timescales, which is a fundamental limitation of all studies of polygenic adaptation [50]. We are also unable to confirm consistency of genetic effects that would be primarily discovered in South Asian samples should large sample sizes have been available.

### F_ST_ enrichment amongst trait associated SNPs

Effect size estimates were largely consistent between ancestral groups, they are unlikely to explain difference in genetic susceptibility to T2D, which indicates systematic differences of allele frequencies at causal variants as an alternative explanation.

Therefore, we next performed an F_ST_ enrichment test, comparing the mean F_ST_ of trait-associated SNPs to a null distribution of LD and MAF matched SNPs representing expectations under genetic drift, and derived empirical two-tailed p-values. T2D, TFM, TFP, BFP and black hair colour exhibited departures from the null distribution, although only T2D, TFM and BH survived multiple testing correction (Holm-adjusted empirical p-values: pT2D = 0.026, pTFM = 0.008, pBH = 0.0008, pBFP = 0.102, pTFP = 0.063), whereas BMI, ALM, and height did not (Holm-adjusted empirical p-values: pBMI = 0.257, pALM = 0.257, pHeight = 0.257) (Supplementary Table 7). As a sensitivity analysis, this test was also repeated matching using LD scores and MAFs generated using the South Asian 1000 Genomes PLINK files, results being generally consistent with the patterns observed using LD scores and MAFs generated from the European PLINK files, although T2D became marginally non-significant after multiple testing correction (Holm-adjusted pT2D = 0.059) (Supplementary Table 8).

**Figure 2:** Beta (departure from the null distribution of LD score and MAF matched SNP sets) ± 1.96 standard errors (derived by dividing the beta by the z score) for all traits when applying the F_ST_ enrichment test to trait associated SNPs clumped at p < 5 × 10⁻⁸ and LD 0.001, within 1000kb windows. Traits with Holm-adjusted significant enrichment are marked with an asterisk.

### XP-EHH and XP-nSL enrichment amongst trait associated SNPs

We next tested for enrichment of haplotype-based selection statistics at trait-associated SNPs relative to matched backgrounds. Considering enrichment of XP-EHH scores amongst trait associated SNPs, none of the traits exhibited departures from the null distribution after adjusting for multiple tests (Holm-adjusted empirical p-values: pHeight = 0.436, pBH = 0.436, pBFP = 0.914, pTFP = 0.855, pTFM = 0.953, pBMI = 0.912, pT2D = 0.855, pALM = 0.489) (Supplementary Table 9).

Whilst both height and black hair colour did exhibit enrichment of XP-nSL scores, which remained after multiple testing correction (Holm-adjusted empirical p-values: pHeight = 0.013, pBH = 0.0008), none of the other traits exhibited enrichment, with all standard errors overlapping the null (Holm-Adjusted empirical p-values: pT2D = 1, pBMI = 0.550, pTFM = 0.550, pTFP = 1, pBFP = 1, pALM = 0.671) (Supplementary Table 11).

As a sensitivity analysis, the procedure was repeated using South Asian MAF and LD scores. Results were generally consistent, with the only change being that the height result in the XP-nSL analysis attenuating (pHeight = 0.365), while black hair colour remained strongly associated (pBH = 0.0008) (Supplementary Tables 10 and 12).

**Figure 3:** Beta (departure from the null distribution of LD score and MAF matched SNP sets) ± 1.96 standard errors (derived by dividing the beta by the z score) for all traits when applying the XP-EHH enrichment test to trait associated SNPs clumped at p > 5×10^⁻8^ and LD 0.001, within 1000kb windows. Traits with Holm-adjusted significant enrichment are marked with an asterisk.

**Figure 4:** Beta (departure from the null distribution of LD score and MAF matched SNP sets) ± 1.96 standard errors (derived by dividing the beta by the z score) for all traits when applying the XP-nSL enrichment test to trait associated SNPs clumped at p < 5 × 10⁻⁸ and LD 0.001, within 1000kb windows. Traits with Holm-adjusted significant enrichment are marked with an asterisk.

### F_ST_ enrichment amongst SNPs associated with T2D clusters identified by Smith et al

To investigate whether genetic differentiation of T2D-associated SNPs (measured by F_ST_) maps to specific disease mechanisms, we leveraged the multi-ancestry T2D genetic clusters defined by Smith et al. [51]. These clusters were derived by applying Bayesian non-negative matrix factorization (bNMF) to a matrix of 650 genome-wide significant T2D variants and their associations with 110 curated traits from multi-ancestry GWAS. SNP cluster assignment is determined based on its pattern of trait associations, and SNPs with a cluster weight ≥0.7802 (the threshold recommended by the original study to maximize the signal-to-noise ratio) for any cluster were included in the set of test SNPs for that cluster.

**Figure 5:** Beta (departure from null distribution) ± 1.96 standard errors (derived by dividing the beta by the z score) for all traits when applying the F_ST_ enrichment test to SNP clusters derived from Smith et al. (see text).Clusters with Holm-adjusted significant enrichment are marked with an asterisk.

Enrichment for F_ST_ was most pronounced amongst the Lipodystrophy 1 cluster, as well as the Lipodystrophy 2 and Liver-Lipid clusters. Whilst enrichment for Lipodystrophy 2 remained after multiple testing correction (Holm-adjusted empirical p-value = 0.006), Lipodystrophy 1 and Liver-Lipid did not (Holm-adjusted empirical p-values: pLipo1 = 0.205, pLiver-Lipid = 0.1419) (Supplementary Table 13). After matching by South Asian MAFs and LD scores, the results remained unchanged (Supplementary Table 14).

### Direction of genetic differentiation

The polygenic score analysis indicated that the fat distribution traits, together with black hair colour, BMI, and T2D, showed higher mean scores in the SAS population and lower mean scores in the EUR population relative to the combined population mean, whereas height and ALM exhibited the opposite pattern, with higher scores in EUR. For several traits (BFP, BMI, T2D), the observed genetic differentiation lay outside the 95% confidence intervals derived from LD- and MAF-matched control SNP sets, exceeding what would be expected under genetic drift (i.e., in the absence of a true relationship between effect size and differentiation), and therefore consistent with adaptive differentiation. It is worth noting however that the traits which showed significant F_ST_ enrichment were not generally the same as those that exhibited differentiation outside expectations under genetic drift (with the exception of T2D and black hair colour).

Previous studies utilising the same methods have investigated potential reasons for such a discrepancy through simulation, highlighting that PS based methods are generally more sensitive to ascertainment bias [52].

**Figure 6:** Genetic differentiation in S.D. units of each population from the overall mean across the two populations. Grey bars indicate the 95% CIs of the distribution of genetic differentiation for the 1000 sets of LD score and MAF matched SNPs for each trait and population combination.

## Discussion

In summary, we demonstrate that loci associated with cardiometabolic disease and fat distribution traits exhibit significant enrichment for F_ST_ between South Asian and European populations, which can be interpreted as evidence of natural selection having driven their cross population genetic differentiation. Enrichment of F_ST_ is also most pronounced amongst regional fat distribution measures (TFM and TFP), as opposed to more general measures of total body fat (BFP and BMI), concordant with current hypotheses regarding elevated South Asian susceptibility to cardiometabolic disease being largely driven by increased visceral adiposity [53]. In contrast, the same loci do not exhibit significant enrichment for cross population haplotype-based selection statistics, as would be expected if their cross population genetic differentiation were attributable to recent strong selective sweeps [54, 55]. Considering the results of our test traits in comparison with those included as positive controls for selection, black hair colour showed robust enrichment across all considered evolutionary statistics (F_ST,_ XP-EHH, and XP-nSL), reflecting well-established population differentiation at pigmentation-associated loci driven by recent directional selection in Europeans [43].

In contrast, height did not exhibit enrichment for F_ST_ or XP-EHH, and its modest enrichment for XP-nSL did not replicate in sensitivity analyses using South Asian reference panels. It is important to note that most previous signals of selection on height reflect analyses conducted within European populations, rather than evidence of cross-population divergence [35–39]. Moreover, many of the early reports of strong selection on height have since been re-evaluated, with several shown to be artefacts of uncorrected population stratification and ascertainment bias in GWAS summary statistics [40, 41]. Consequently, while height remains a useful benchmark for selection on polygenic traits in comparison to the included fat distribution traits, within this context, it is not necessarily unsurprising that height does not exhibit pronounced F_ST_ or XP-EHH enrichment.

Whilst it is possible that the higher polygenicity of height and resultant increased number of trait associated SNPs may have led the F_ST_ enrichment test to become overconservative in comparison to black hair colour, F_ST_ enrichment was also not observed for many of the T2D clusters. These clusters have a comparably low number of test SNPs (the lowest being 2 and the highest being 78) as the clumped black hair colour associated SNPs (80).

Additionally, in our PS based analysis, we were able to infer the direction of this genetic differentiation, with all of the fat distribution traits as well as BMI and T2D showing an elevated mean in South Asian population relative to the combined European and South Asian mean. We interpret these results as complementary to the results of the F_ST_ enrichment test, and assume that the direction of genetic differentiation is correct, especially for traits with larger mean differences between the two populations. However, it is important to note that this test is qualitative and that no strong quantitative inferences can be drawn regarding the strength of selection or distribution of true genetic values in the two populations, given the numerous caveats associated with applying polygenic scores to study selection and genetic differences across populations that have been thoroughly discussed elsewhere [56–58]. Briefly, theoretical work demonstrates that pervasive negative selection can generate differing genetic architectures and causal variants for the same trait in separate populations [59], and similarly stabilizing selection (around a shared phenotypic optimum) and the resultant allelic turnover can increase the differentiation of trait associated loci between the two populations compared to expectations under drift [56]. In both cases, differences in genetic architecture underlying the trait in the two populations can in turn lead to erroneous overestimation of population differentiation, since causal loci ascertained in one population will explain a lesser extent of the variance in the alternative population. Both the F_ST_ enrichment test and the PS-based method fundamentally assume that the causal loci underlying trait variation are consistent between populations, and so are vulnerable to such biases.

Our results are generally consistent with the demonstration that, amongst global populations, high F_ST_ SNPs lack strong haplotypic patterns as expected if the favoured allele had reached fixation rapidly [54, 55]. This discrepancy has been interpretated as evidence in favour of soft sweeps on standing variation, in combination with genetic drift, as the major forces shaping cross population genetic differentiation, as opposed to hard sweeps on newly arising alleles [55]. This interpretation is important to consider in the context of studying cross population differentiation of complex traits, given that subtle coordinated allele frequency shifts characteristic of polygenic adaptation may leave too small a signal at individual loci underlying complex trait variation to be detected by conventional methods, which are better powered to detect classic sweeps [54, 55, 60, 61]. Additionally given that the F_ST_ statistic is based solely on variances in allele frequencies between populations, it is not possible to determine the timescale over which the observed differentiation may have occurred.

Whilst many recent studies have emphasised the role of rare ancestry stratified variants in contributing to differential cardiometabolic disease risk in South Asians [15], our results conversely suggest that a substantial component of excess cardiometabolic disease risk in South Asian populations may be attributable to cosmopolitan genetic variants shared across ancestries. This finding contrasts with expectations had excess cardiometabolic risk been driven predominantly by rare, ancestry stratified variants, under which common variants would not be expected to exhibit pronounced population differentiation. Differentiation among common variants further supports the view that observed genetic differences primarily reflect genetic drift and/or weak selection acting on standing variation, rather than strong selective sweeps. Nonetheless, it remains plausible that rare ancestry stratified variants also account for an appreciable component of excess risk, especially when considering the extensive genetic diversity and population structure that exist within South Asia [62, 63]. Therefore, in order to gain a comprehensive understanding of the combined contribution of common and rare genetic variation and improve prediction of cardiometabolic disease in South Asian populations, it remains crucial to increase the sample sizes of South Asian specific GWAS of cardiometabolic disease traits to those comparable with current European sample sizes [63, 64]. Moreover, the increase in the availability of high-quality whole genome sequences from South Asian populations will allow the application of methods that are more sensitive to polygenic adaptation [39, 65] and additionally ongoing advances in the understanding of the demographic and evolutionary history of South Asia could potentially allow the evolutionary timescales and ancestral populations in which selection occurred to be discovered [66].

## Methods

### Data Sources

There is substantially reduced power to detect true causal variants in non-European specific GWAS due to lower sample sizes [66], which we attempted to alleviate by leveraging statistical power from large European GWASs to identify loci that are relevant for both European and South Asian samples. We assume that differences in marginal genetic effects between populations that are estimated from GWAS will likely arise due to a combination of differential LD for tagging variants, or interaction with genetic or environmental factors that are differential distributed across the two populations. In order to minimise the impact of LD reducing replicability of GWAS discovery variants across populations we selected per-locus sentinel variants from a cross-population fixed effects meta-analysis for all per-population discovery loci, clumping using a combined ancestry panel.

All GWAS summary statistics (with the exception of appendicular lean mass) were sourced from large multi-ancestry GWAS meta-analyses. The T2D summary statistics were sourced from the latest release of the Type 2 Diabetes Global Genomics Initiative [67], whereas the height summary statistics were sourced from the latest Genetic Investigation of Anthropometric Traits (GIANT) consortium release [68]. Summary statistics for body fat distribution traits—trunk fat percentage (UKB Field ID 23128), trunk fat mass (UKB Field ID 23129), body fat percentage (UKB Field ID 23099), and body mass index (UKB Field ID 23104)—as well as for black hair colour (UKB Field ID 1747; categorical phenotype, category 5) were sourced from the Neale lab analysis of 7,221 phenotypes across six continental ancestry groups in the UK Biobank [69].

Summary statistics for the two ancestry groups (European and South Asian) were downloaded separately or split if originally in a single file.

Appendicular lean mass (ALM) was derived in UK Biobank participants as the sum of fat-free mass of all four limbs (left and right arms and legs) from impedance measures. Phenotype files were generated after quality control exclusions, including discordance between self-reported and genetically inferred sex, sex chromosome aneuploidy, heterozygosity/missingness outliers, related individuals, and other recommended genomic exclusions, retaining only participants with complete limb fat-free mass data. Covariates included age, inferred sex (binary-coded), the first 20 genetic principal components, and derived terms (age², age × sex, age² × sex). Genotype data (array-based) were merged across autosomes and subjected to variant-level quality control (MAF >0.01, MAC ≥20, missingness <10%, HWE p >1×10⁻¹⁵), yielding a high-quality SNP set for model training. GWAS was conducted using REGENIE on the UK Biobank Research Analysis Platform in two steps: (i) step 1 fitting whole-genome regression models with leave-one-chromosome-out cross-validation, and (ii) step 2 single-variant association testing across imputed genotype data (UKB v3 release), restricted to variants passing imputation and QC filters (INFO, MAF, MAC thresholds). Analyses were performed separately in South Asian (N = 9,163) and White European (448,711) ancestry groups, and chromosome-level summary statistics were merged to generate genome-wide results.

Ancestry specific summary statistics were standardised (such that the effect and non-effect allele are oriented alphabetically, and effect size also oriented accordingly) and subsequently meta-analysed using the inverse variance weighted scheme implemented in METAL [70]. Meta-analysis results generated by METAL were subsequently clumped using PLINK 1.9 [71] with a combined ancestry reference panel at a threshold of p < 5 × 10⁻⁸, at an LD threshold of 0.001 and within a clumping window of 1000 kb.

### Quality Control

Prior to meta-analysis, quality control was performed separately for each ancestry, and subsequently repeated with summary statistics generated as a result of the meta-analysis. The distribution of genome-wide test statistics showed greater inflation in European (EUR) GWAS compared with South Asian (SAS) GWAS, as reflected by higher lambda values both before and after meta-analysis (Supplementary Tables 1–2). LDSC was performed using LDSC v1.0.1 [72], in order to ascertain how much of the observed inflation was due to confounding. EUR GWAS generally exhibited LDSC intercepts above the nominal threshold of 1.1 and higher ratios, whereas SAS GWAS had intercepts closer to 1 and lower ratios (Supplementary Table 3). These findings indicate that EUR summary statistics were more affected by residual confounding, likely due to population structure or cryptic relatedness. However, given that a substantial component of inflation is nonetheless likely attributable to polygenicity alone, genomic control correction was not applied, as this can lead to overcorrection and reduced power when inflation is driven in part by true polygenic signal.

### Evaluation of consistency of association and effect size between populations

To confirm the consistency of effect sizes for sentinel variants selected from the fixed effect meta-analysis between populations, we implemented a heterogeneity test that generates a per variant Cochran’s Q statistic, based on its effect size and standard error in the two populations.

Concordance of the effect sizes and p-values of trait-associated variants was also compared by means of a binomial test, implemented through a replication analysis that estimates expected versus observed replication rates under the assumption of unbiased discovery effects.

Differences in statistical power between the discovery and replication datasets are accounted for indirectly via the standard errors of the effect estimates. These are used to model both the probability of concordant effect directions and the likelihood of achieving replication significance under the assumption that the discovery effect is true. While sample size is not explicitly modelled, it influences power through its contribution to the standard error.

### Generation of population specific MAFs and LD scores

PLINK binary files specific to each 1000 Genomes superpopulation were downloaded from: https://cncr.nl/research/magma/. Minor allele frequencies (MAFs) for all SNPs present in these PLINK binary files were computed using the --freq command in PLINK.

For all SNPs in the superpopulation specific.bim files, centimorgan (cM) positions were interpolated using the --cm-map command in PLINK 1.9, with genetic maps provided in IMPUTE-m format, available at: https://mathgen.stats.ox.ac.uk/impute/1000GP%20Phase%203%20haplotypes%206%20October%202014.html. Subsequently, superpopulation-specific LD scores were computed for all SNPs within a 1 cM window using the ldsc.py --l2 command from LDSC v1.0.1 [67].

Following the methodology of Guo et al. [34], all SNPs were then binned into 20 MAF bins (ranging from 0 to 0.5 in increments of 0.025) and 20 LD score bins based on the 20 quantiles of the LD score distribution.

### VCF download and processing

Variant Call Format (VCF) files were obtained from the 1000 Genomes Project Phase 3 release (ftp.1000genomes.ebi.ac.uk/vol1/ftp/release/20130502/). To ensure analyses were restricted to high-confidence genomic regions, VCFs were then filtered using the 1000 Genomes “pilot mask,” which defines accessible regions of the autosomes based on reliable sequencing and variant calling criteria, before subsequently filtering to retain only biallelic sites, using bcftools 1.19 [73].

### Generation of whole genome Weir and Cockerham’s F_ST_ values

Weir and Cockerhams F_ST_ [74] between the European and South Asian superpopulations were then calculated using the vcftools –weir—Fst—pop command. Negative F_ST_ values were retained, in order to avoid biasing the distribution of F_ST_ values upward.

### Generation of haplotype-based statistics

XP-EHH and XP-nSL scores were generated using selscan 2.03 [75], with default parameters in both cases, such that no SNPs with MAF below 0.05 are used as focal sites when constructing haplotypes, extending a maximum gap of 20 kb between consecutive SNPs when assembling haplotypes and truncating calculation when the extended haplotype homozygosity pooled across populations decays below 0.05 or the marker extends more than one 1Mbp from the core marker.

Per chromosome vcf files were then converted to PLINK binary files using the --make bed command in PLINK1.9, before cM positions were interpolated using the –cm-map command, using the same impute ‘-m’ format genetic maps mentioned in preceding paragraphs, in order to obtain PLINK –map format files required for haplotype construction within selscan.

Unstandardised XP-EHH and XP-nSL scores were normalised genome wide using selscans companion script ‘norm’. Haplotype based statistics were not calculated with respect to the ancestral or derived allele but rather according to the reference and alternative allele as specified in the VCFs.

### F_ST_ XP-EHH and XP-nSL enrichment tests

Following Guo et al.[34] an F_ST_ enrichment test was applied that compares the mean fixation index (F_ST_) of trait associated SNPs compared to a null distribution constructed of 10000 sets of LD score and MAF ‘matched’ SNPs. This test was also further extended to examine the enrichment of two cross population haplotype-based statistics, XP-EHH[44] and XP-nSL[45], amongst trait associated variants.

The test is performed in the following way; for each set of trait associated SNPs, the mean of the respective statistic is calculated, which is then compared to a null distribution constructed of the means of each statistic of for 10000 ‘matched’ sets which contain an equal number of SNPs of each LD bin and MAF bin combination as the original set of trait associated SNPs. By comparing the mean of the trait associated SNPs to the null distribution, an empirical two tailed p-value is derived. Before matching, the whole genome values of each statistic were also filtered to contain only common (MAF >0.01) SNPs present in both the European and South Asian PLINK files, as well as SNPs with Hardy-Weinberg p-values *p > 5×10⁻^6^*, a less stringent threshold chosen to retain SNPs under selection that may exhibit minor deviations from HWE.

### Determining the direction of genetic differentiation

In order to determine the direction of genetic differentiation, we next employed a polygenic score-based method previously used in tandem with the F_ST_ enrichment test [34], in which for each respective trait the genetic differentiation of each population was expressed as SD units of the mean of the polygenic score in each population from the overall combined mean of the two populations. In order to visualise expectations under genetic drift, we also repeated this process for 1000 sets of SNPs matched (using the same procedure as the enrichment tests) on MAF and LD scores to those used to construct the original polygenic score. As a sensitivity analysis, this process was once again repeated matching using MAFs and LD scores generated using the South Asian PLINK files, which did not alter the direction of the genetic differentiation for any trait (Supplementary Figure 61)

## Data Availability

All data produced in the present study are available upon reasonable request to the authors

---

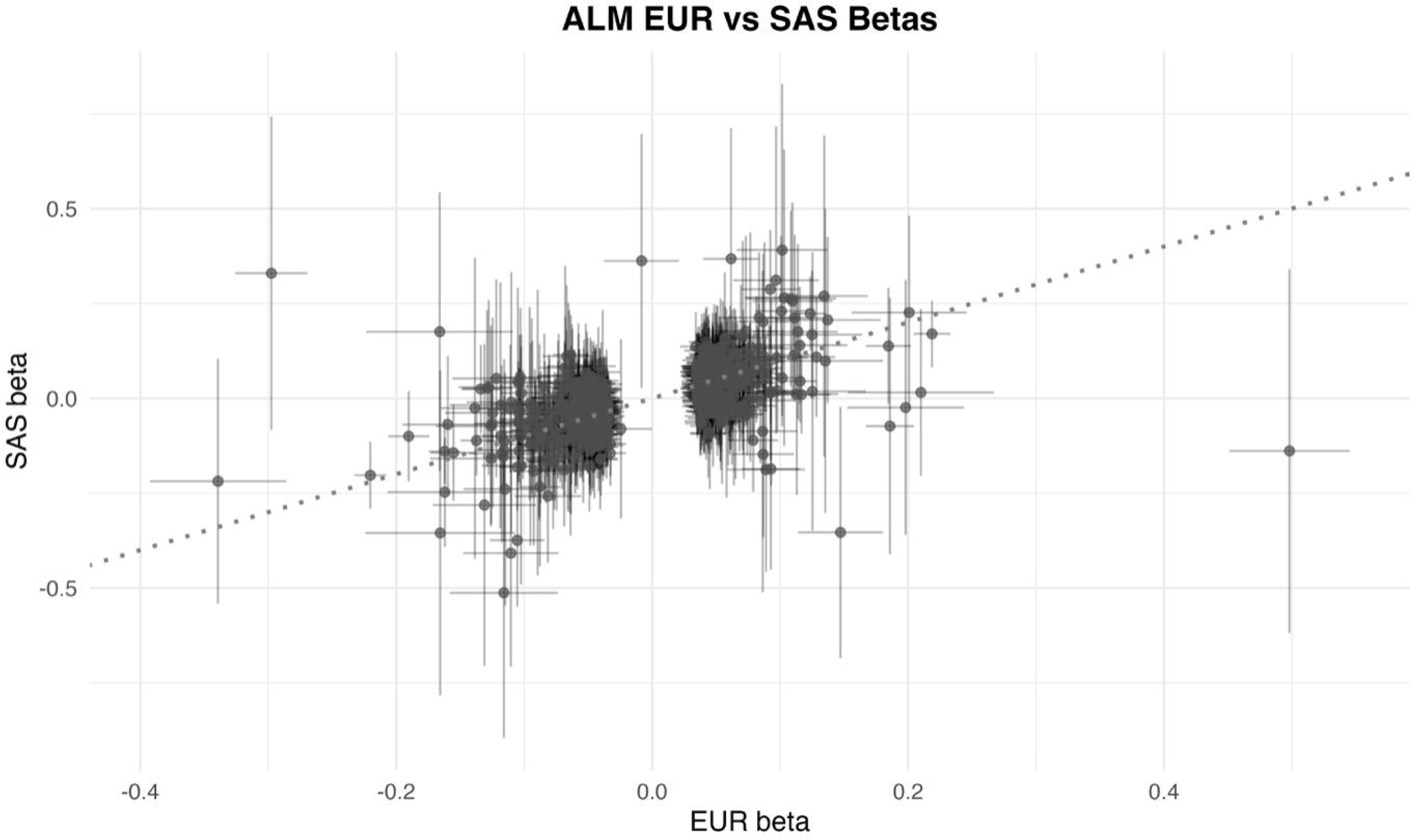

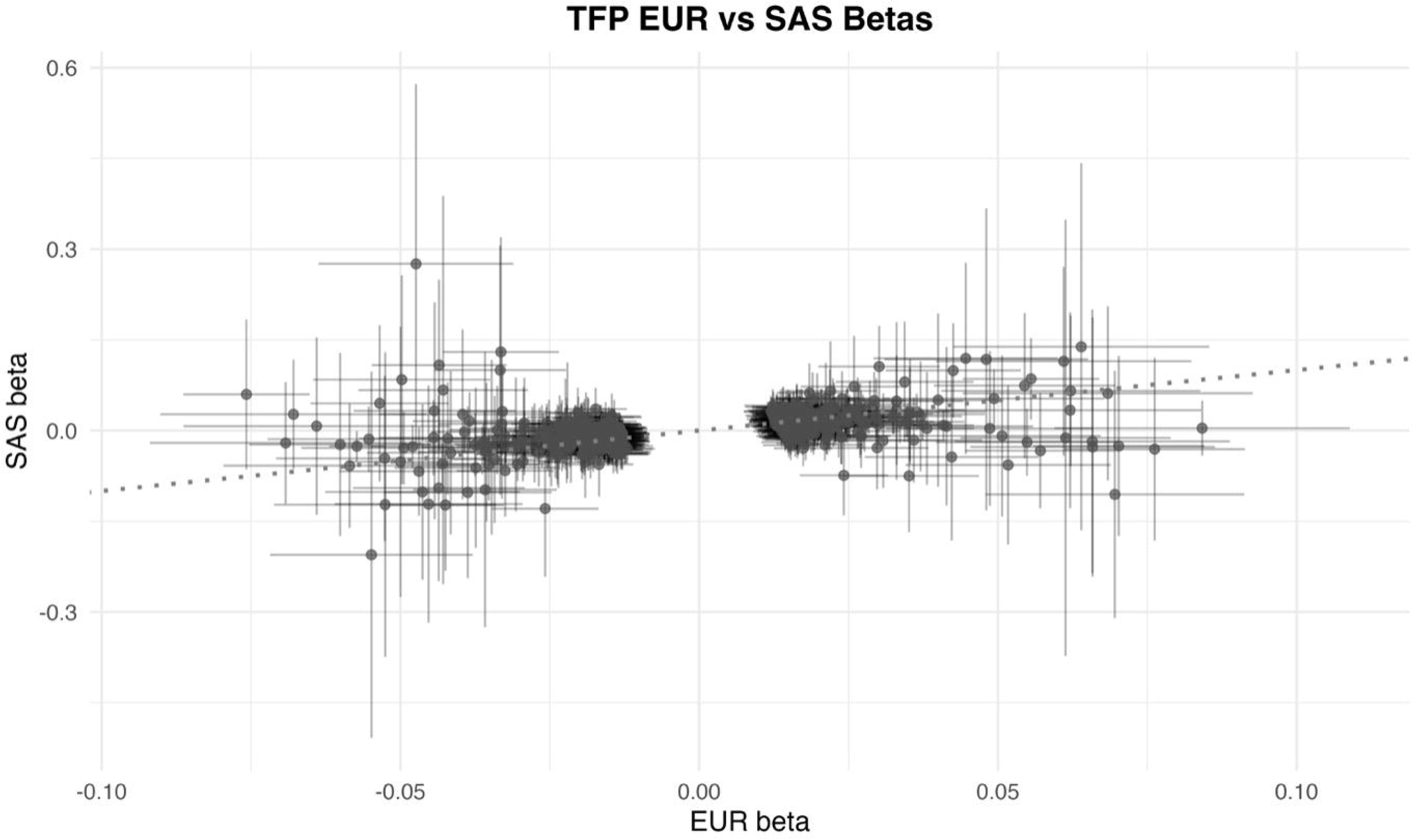

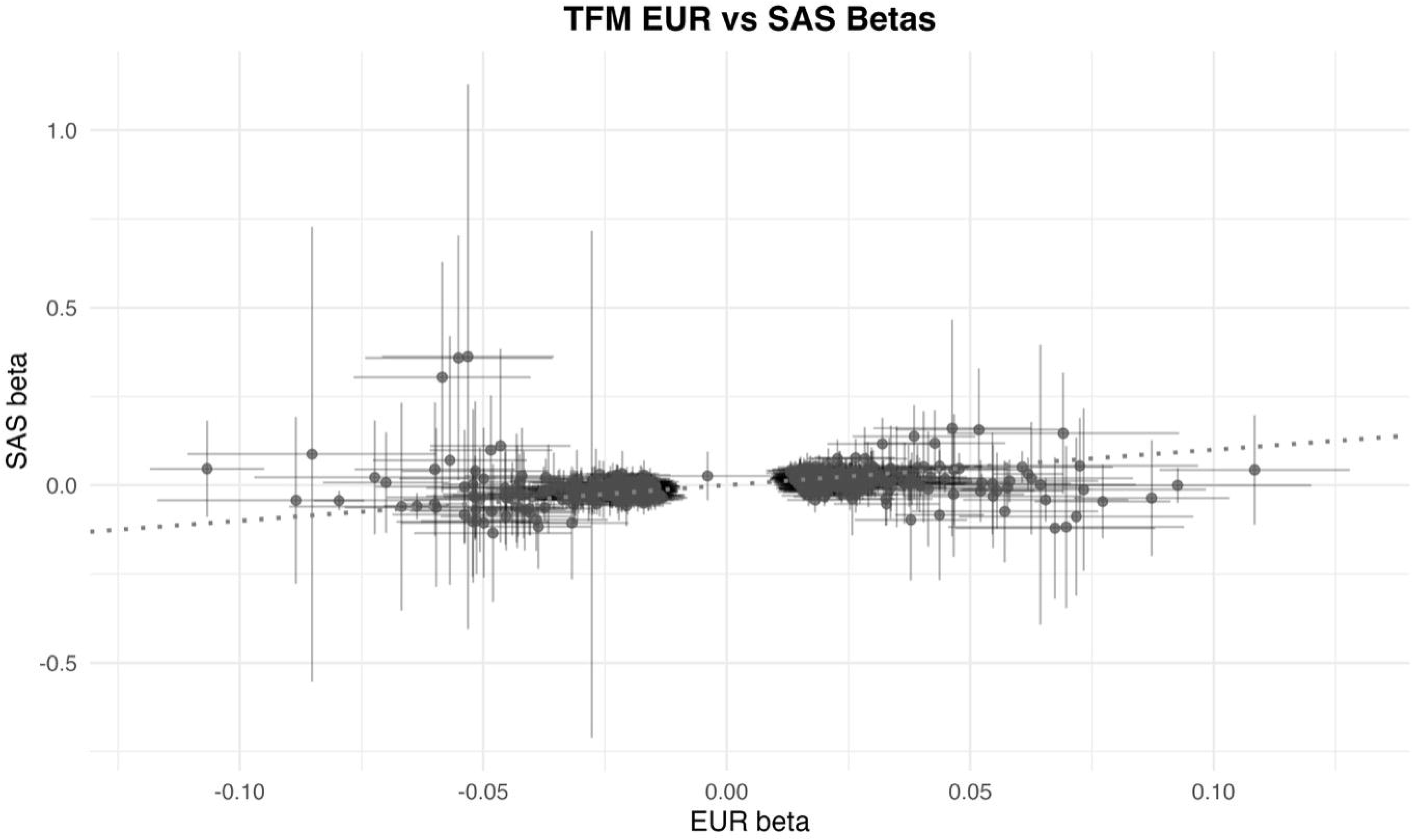

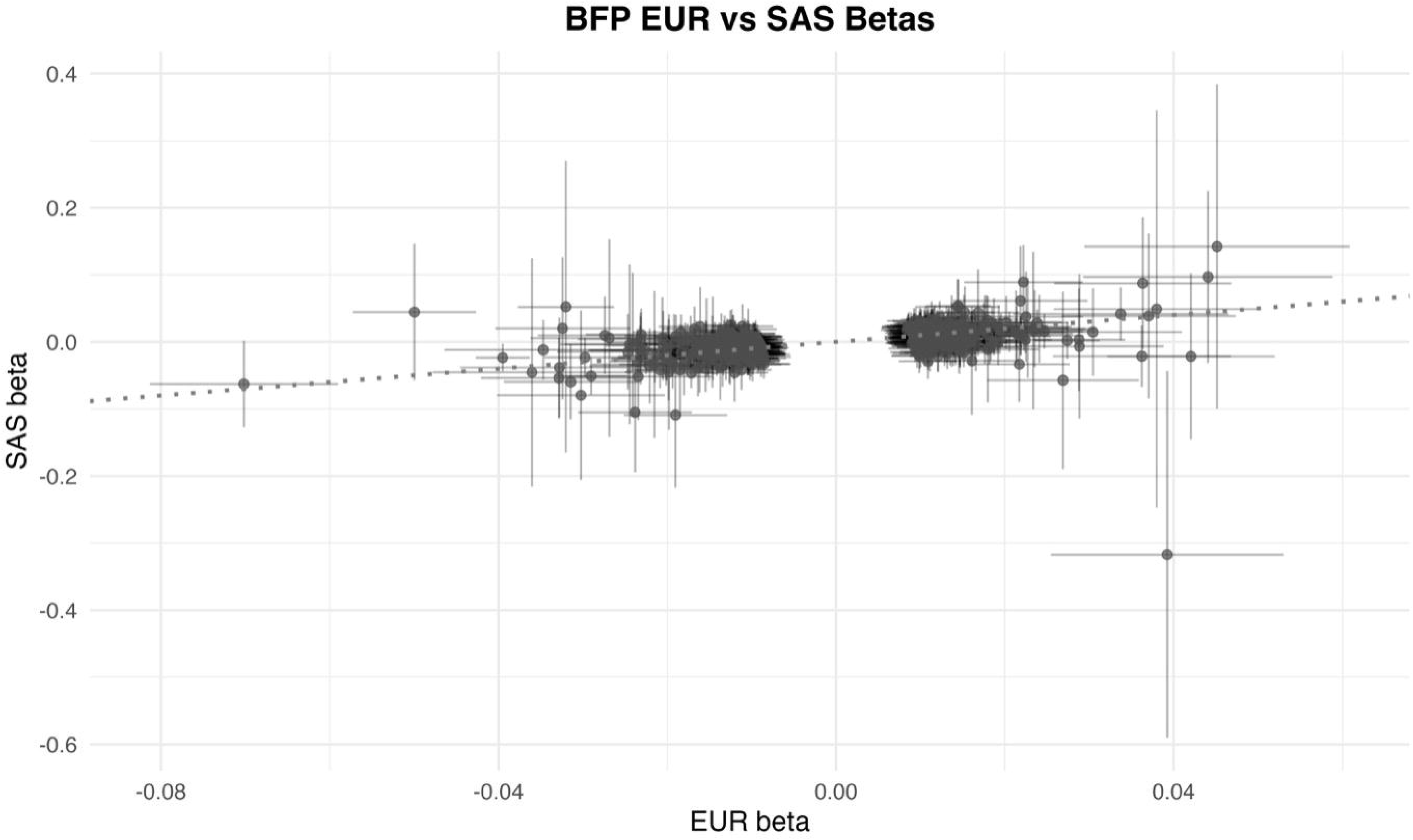

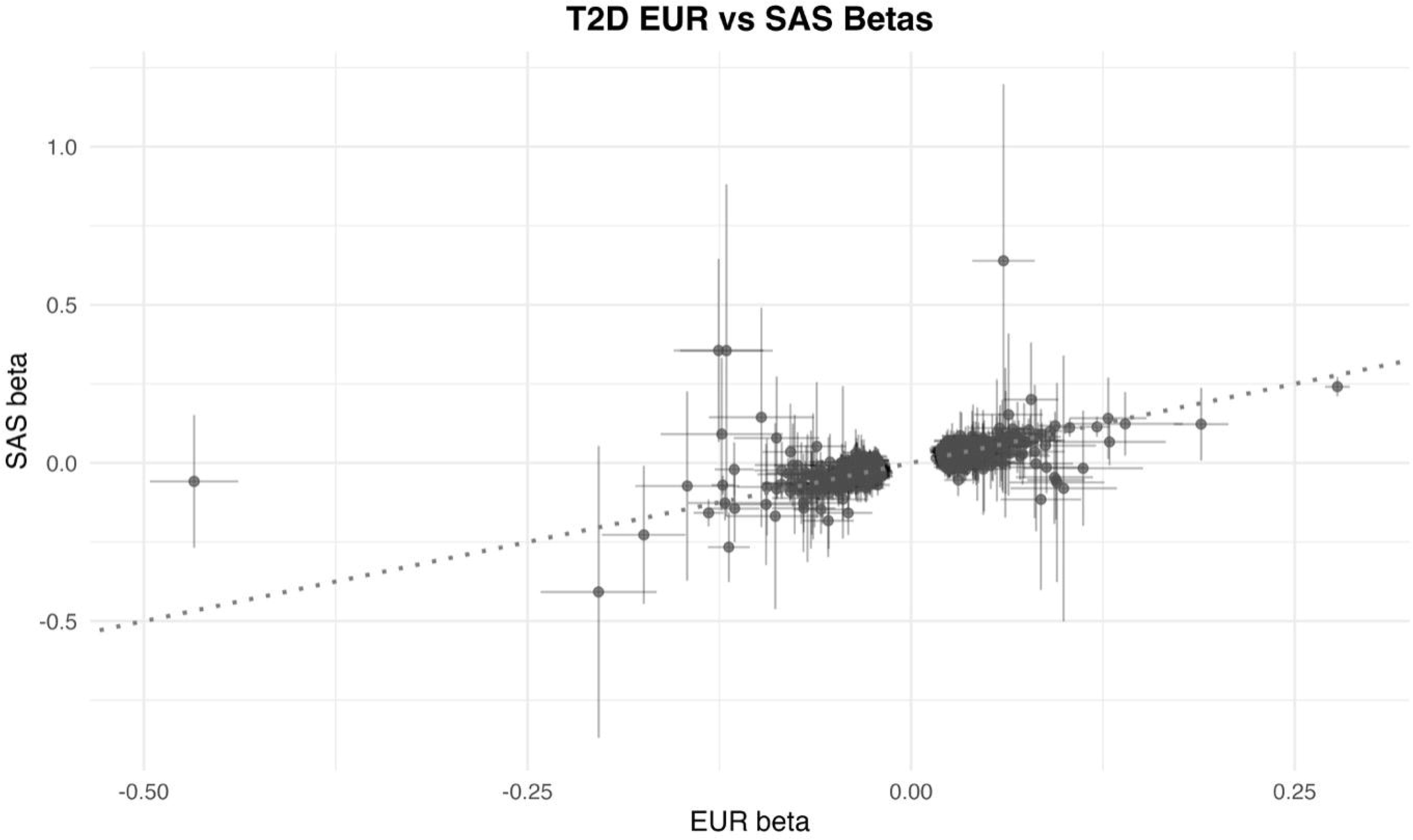

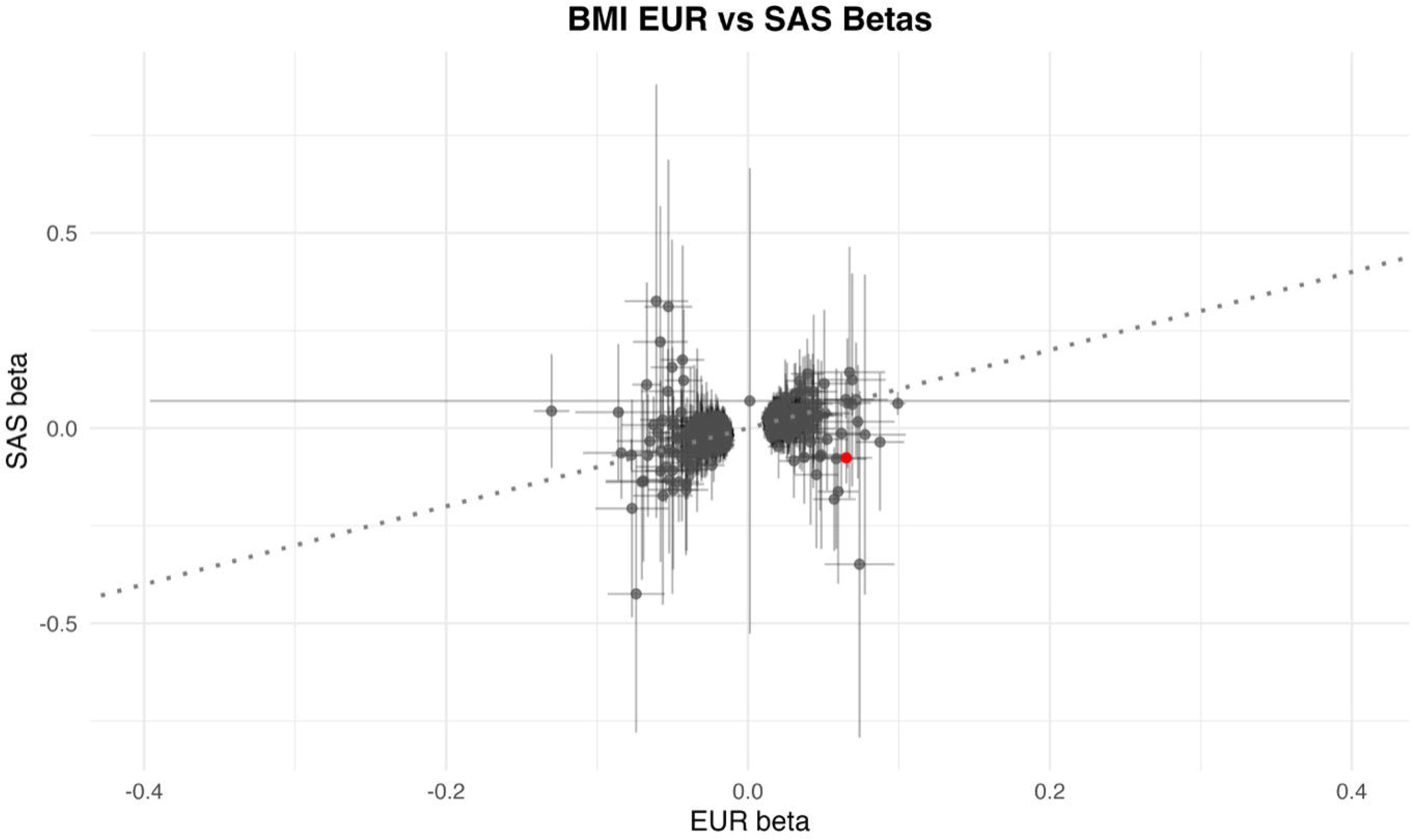

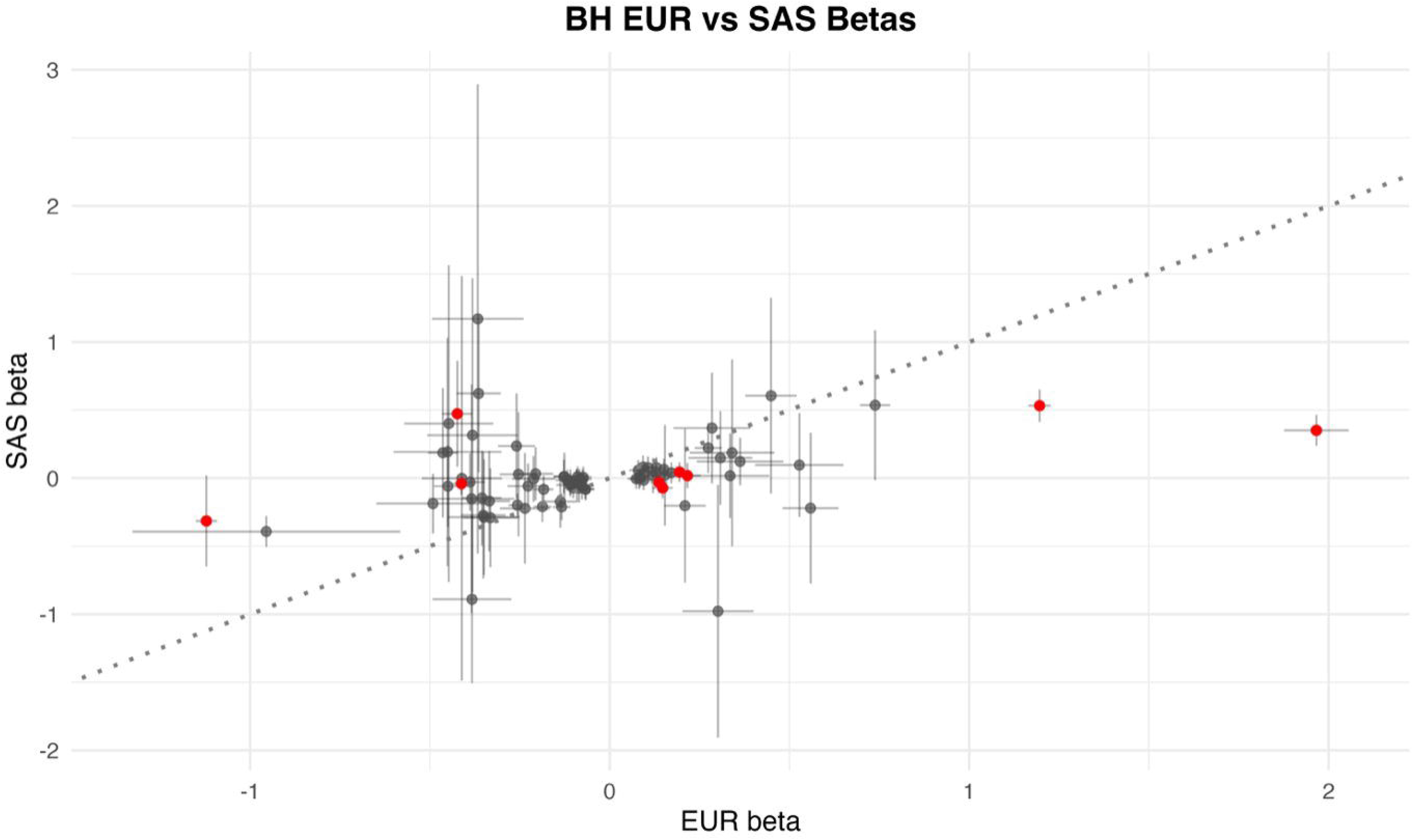

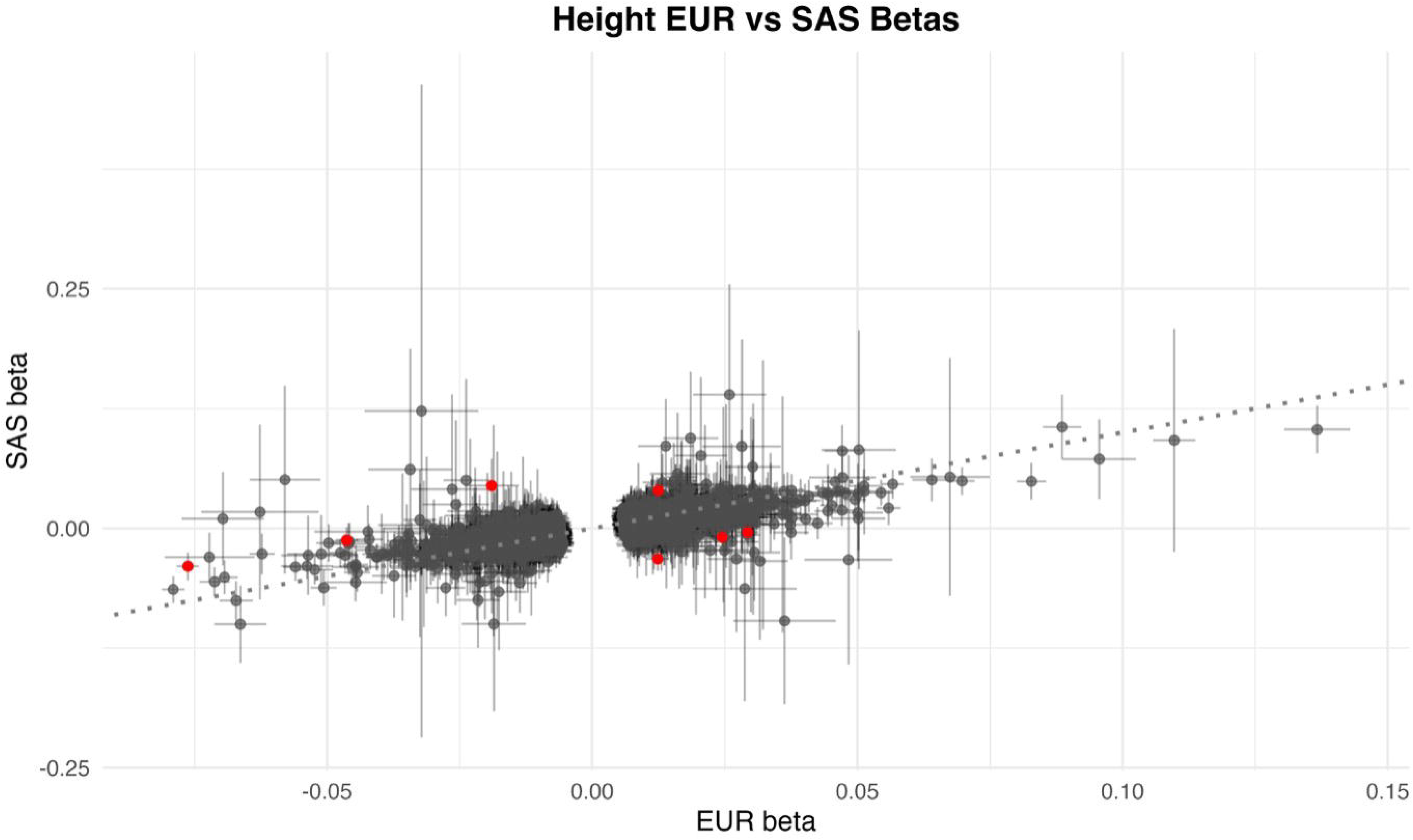

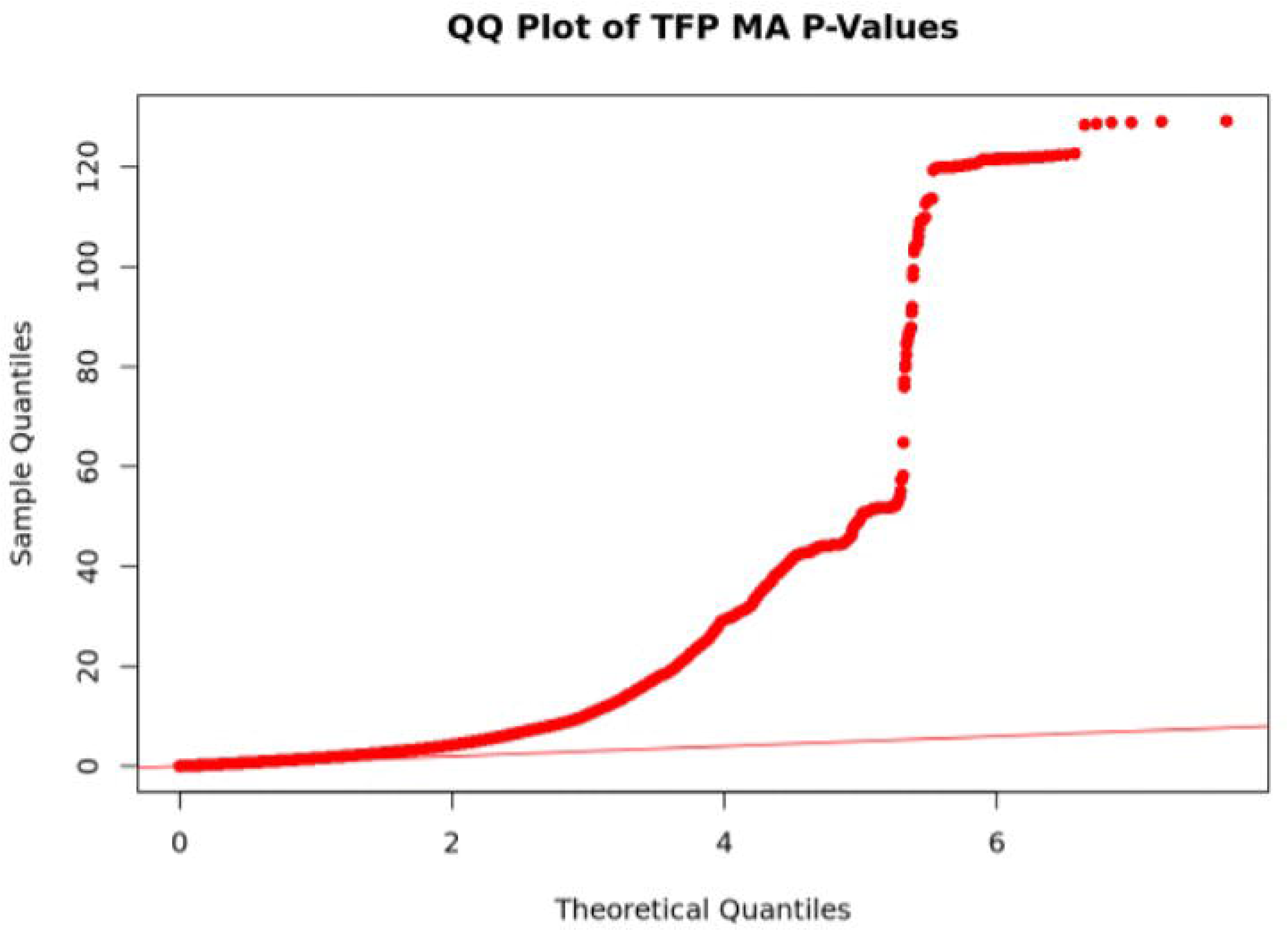

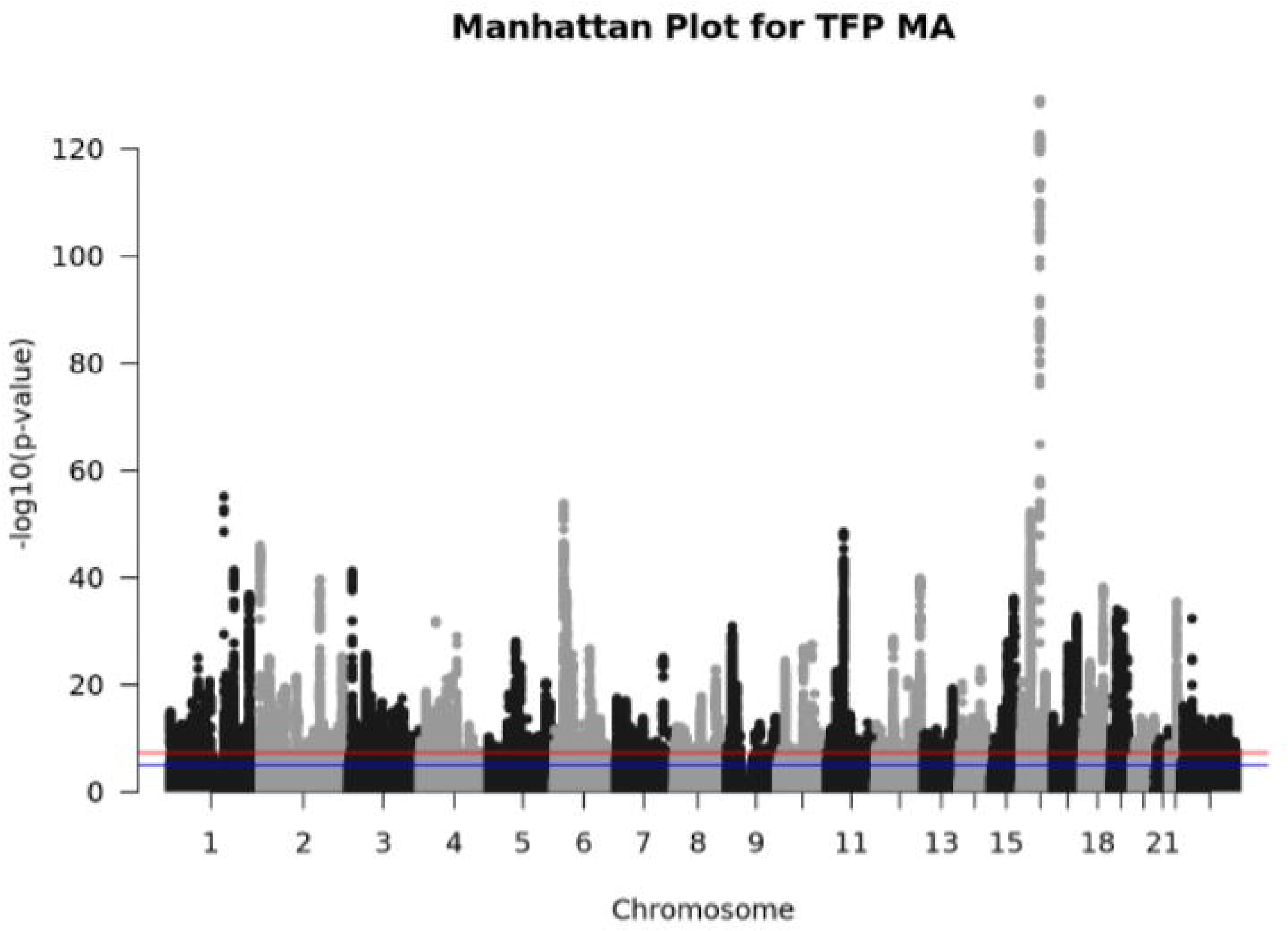

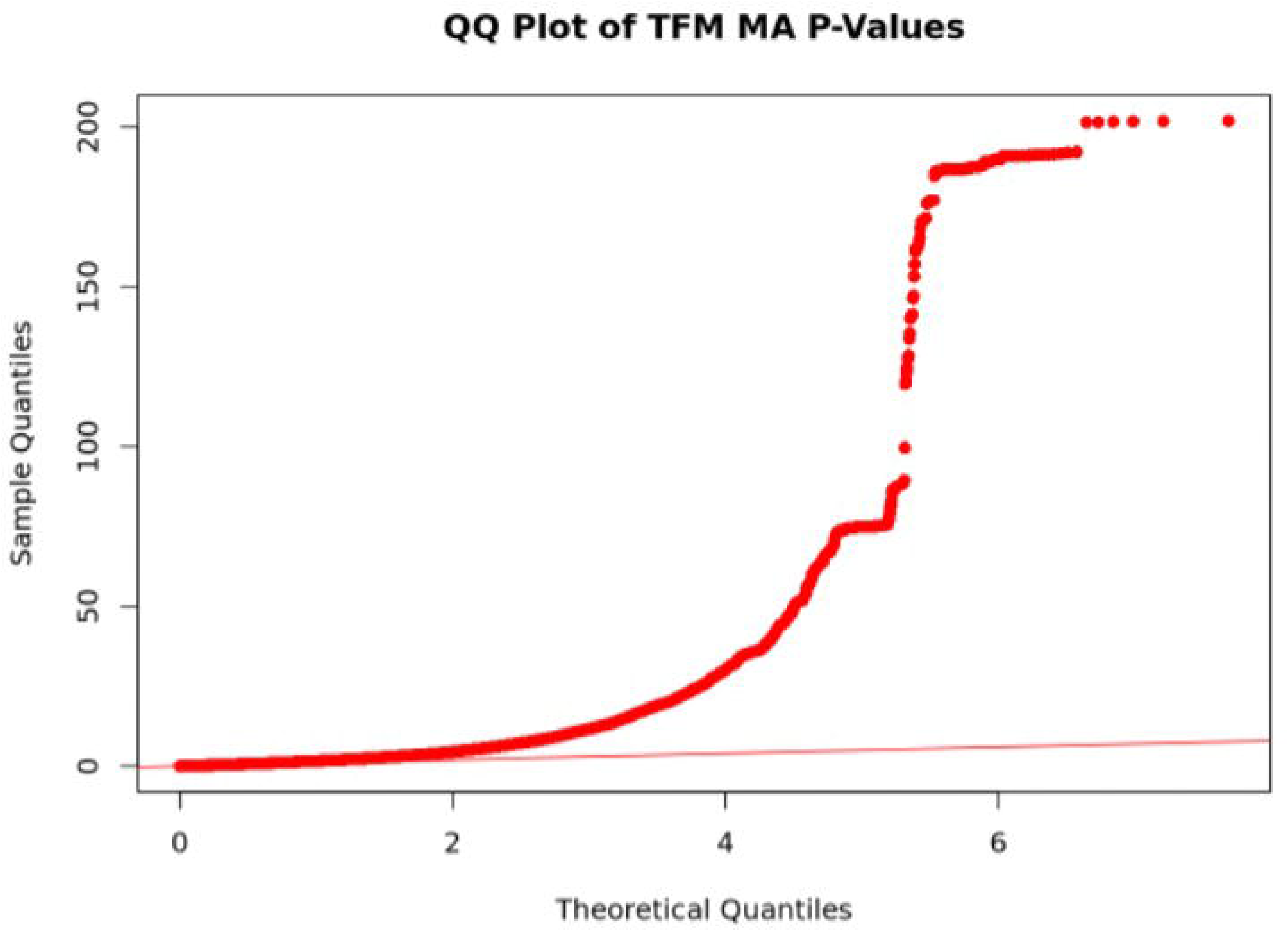

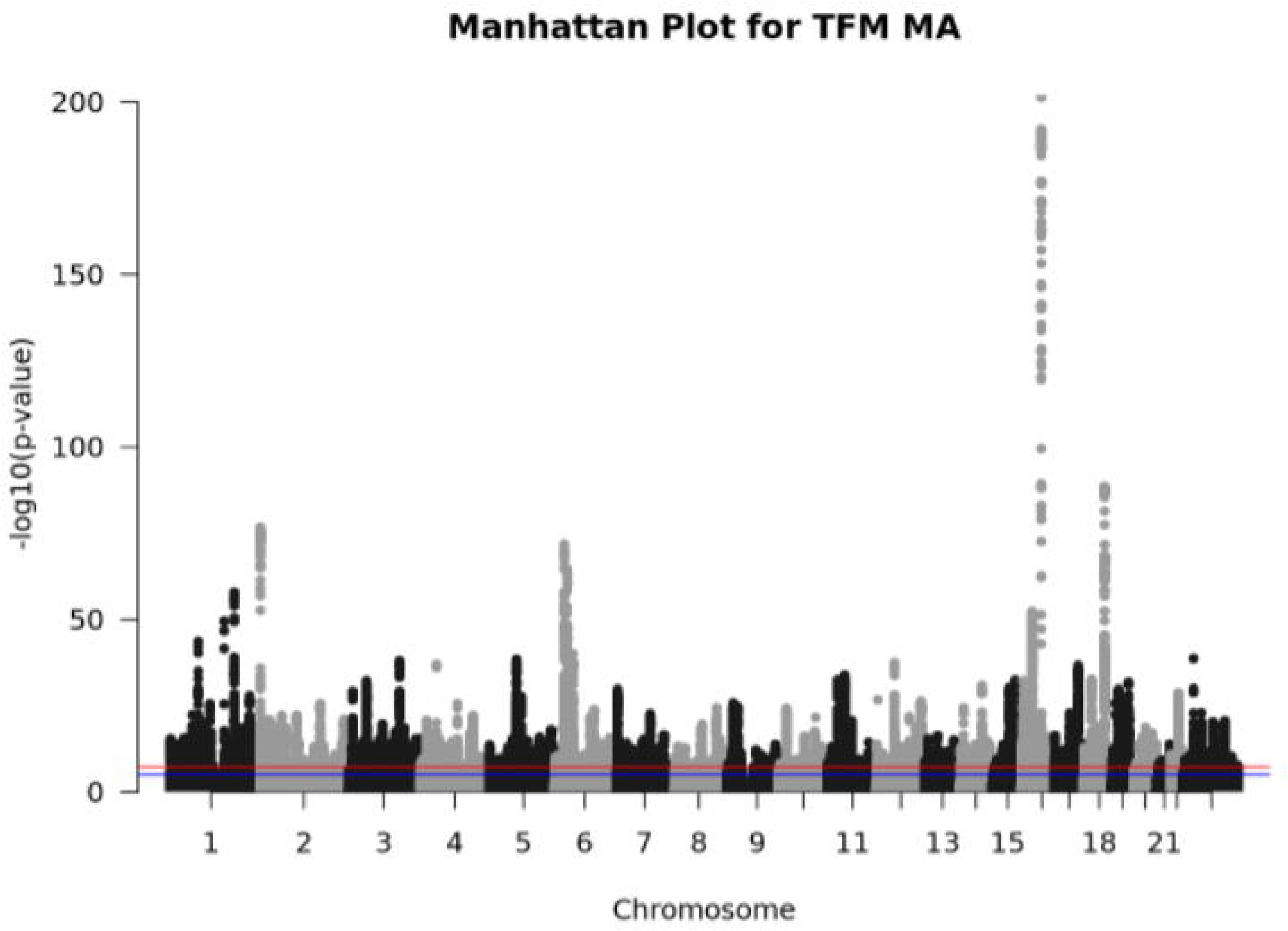

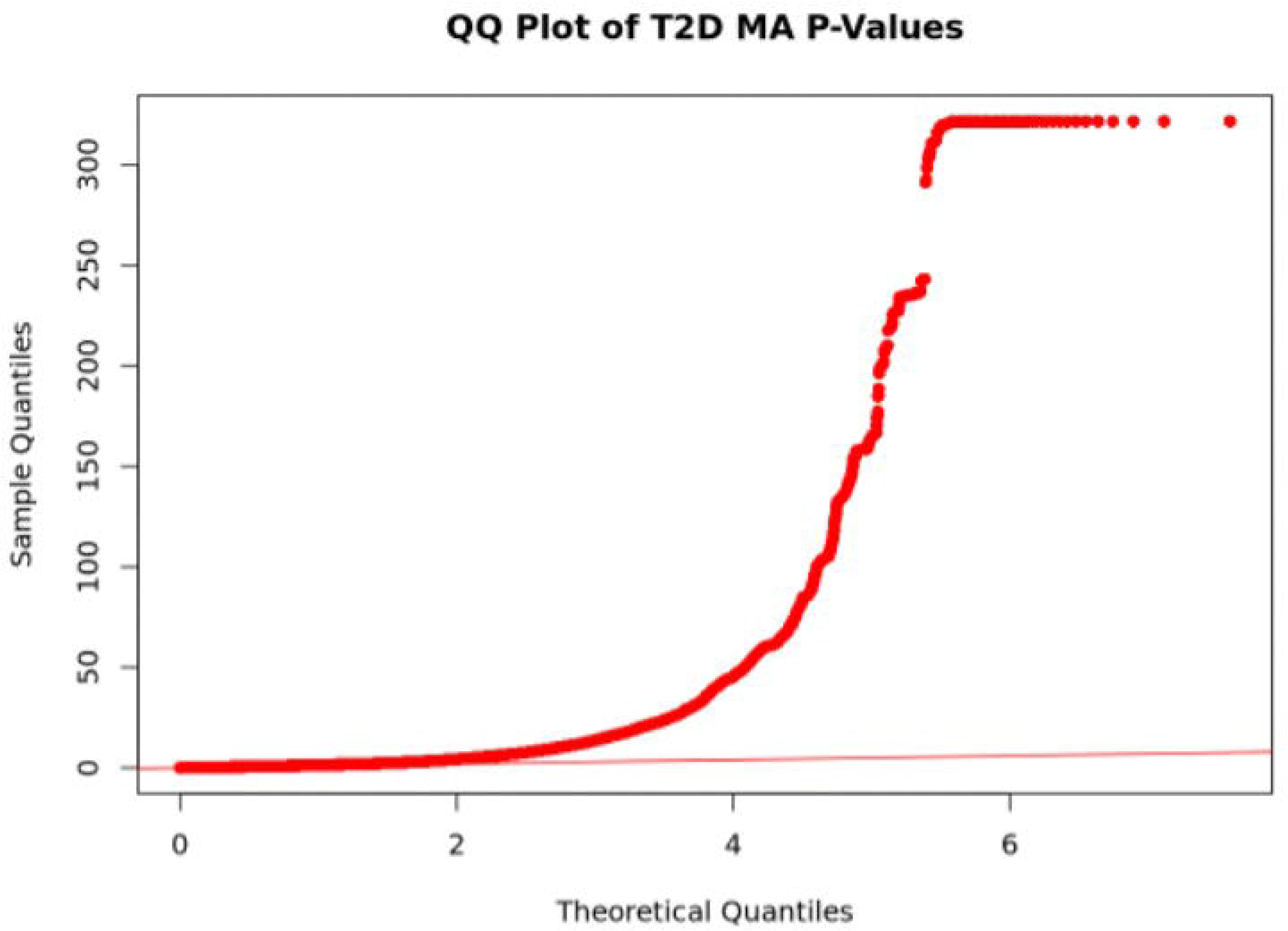

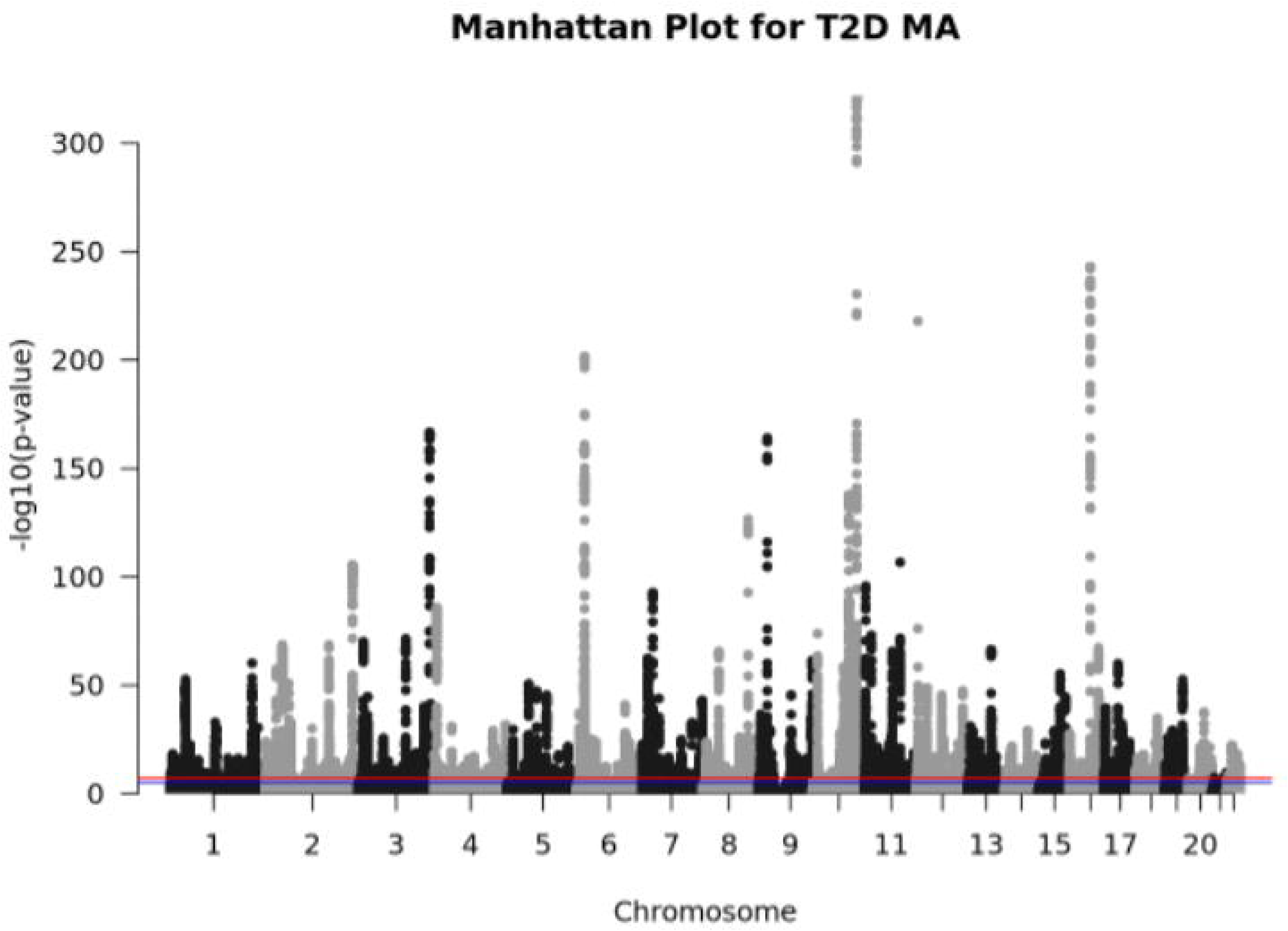

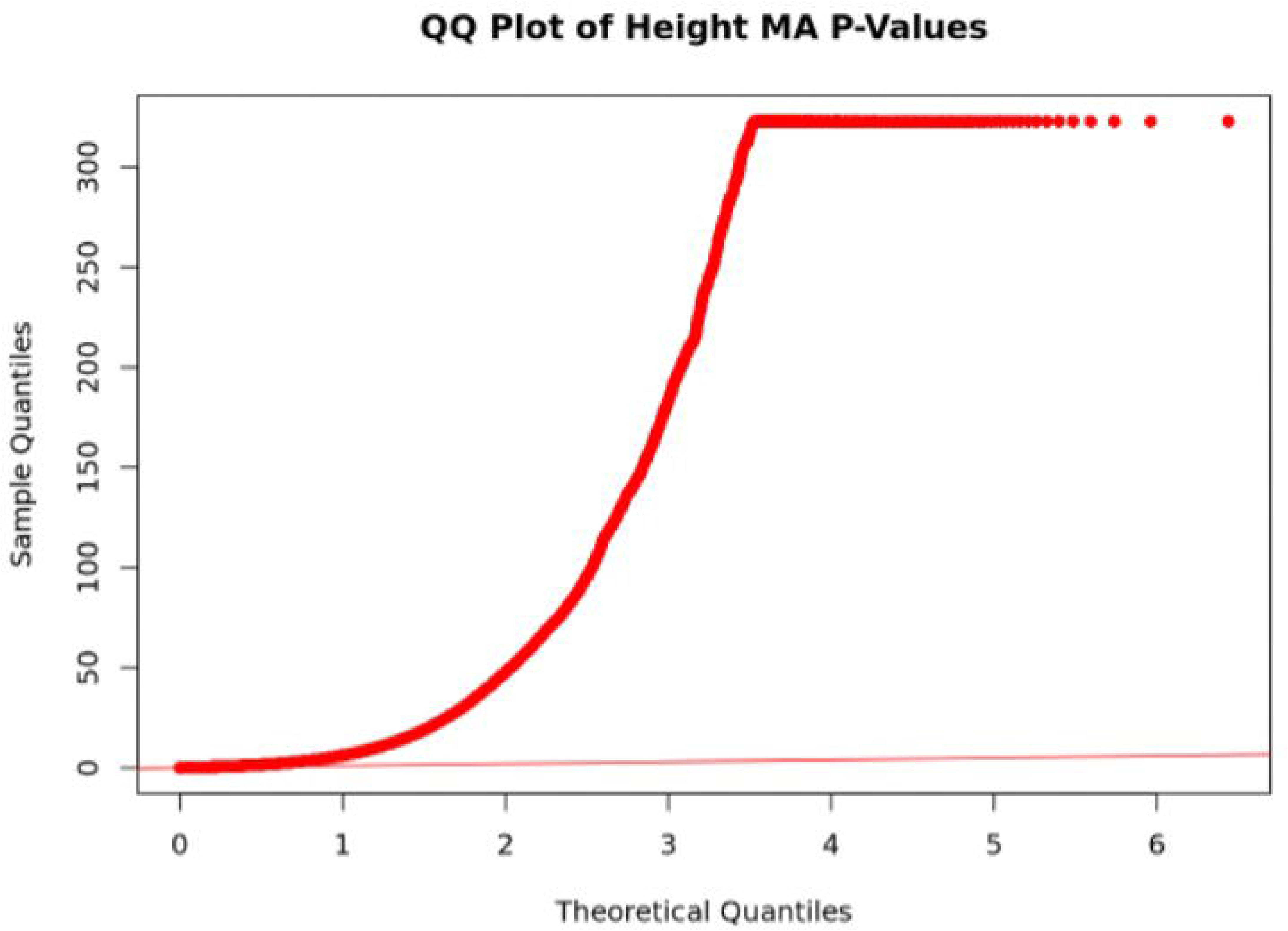

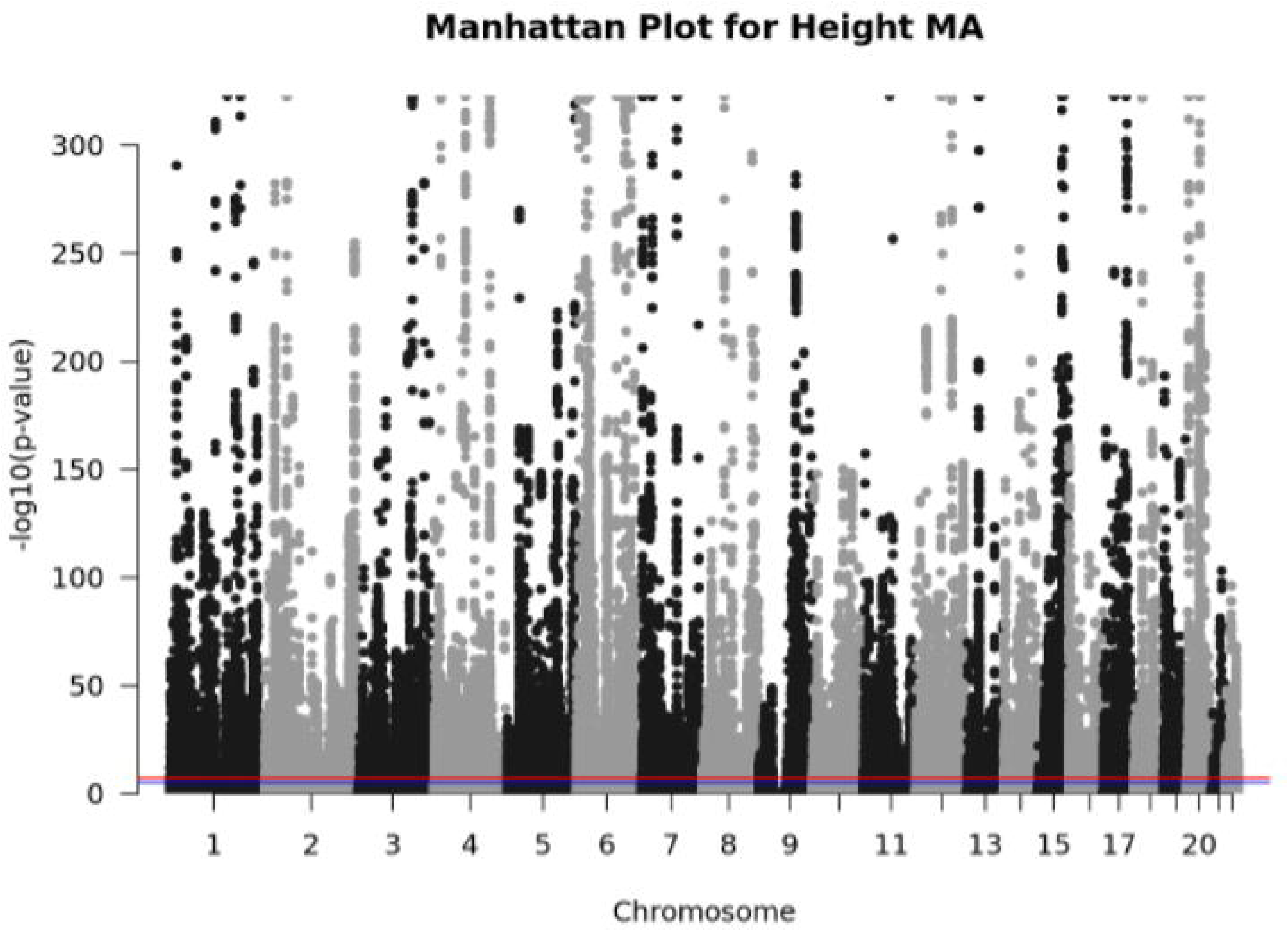

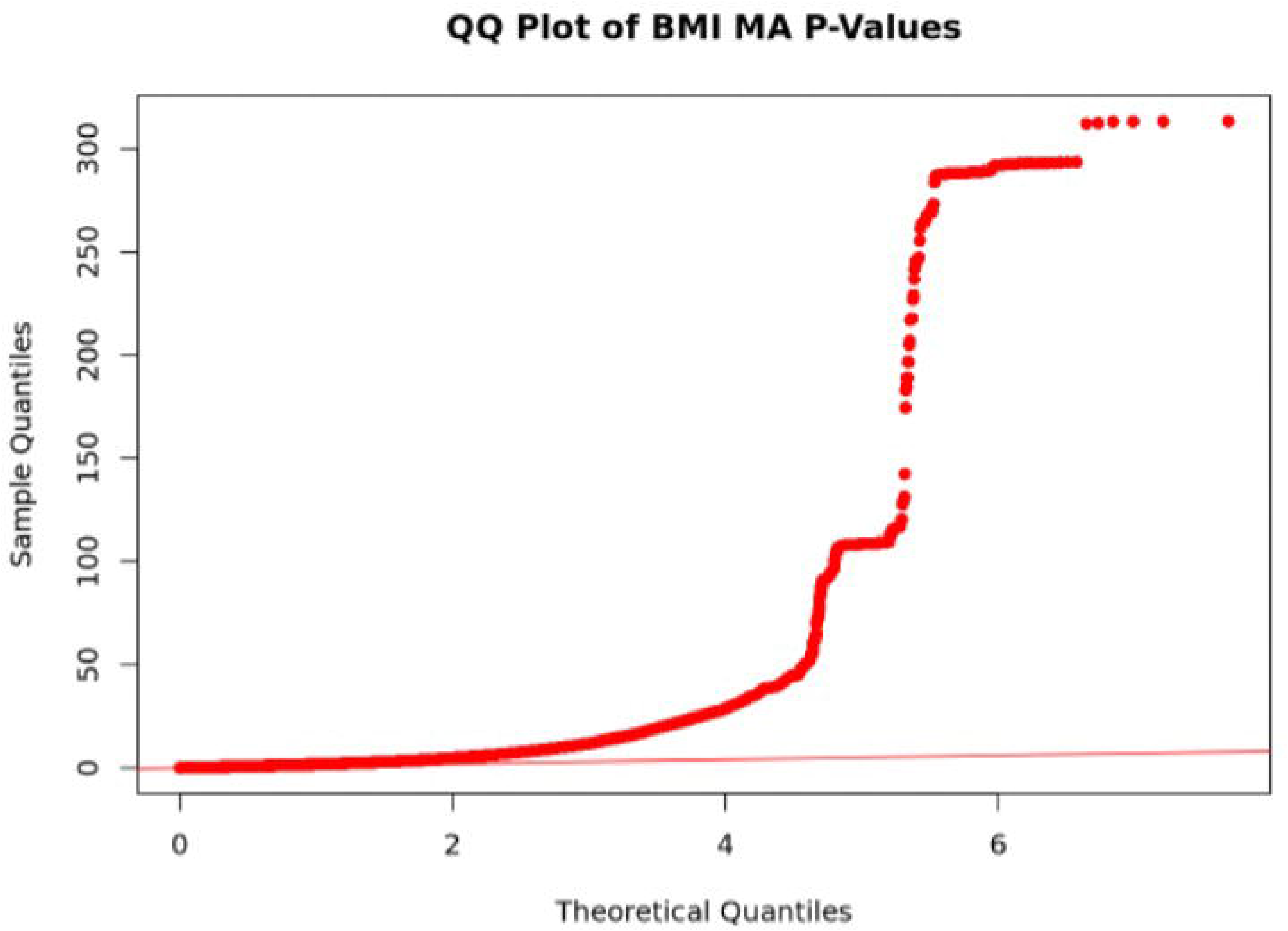

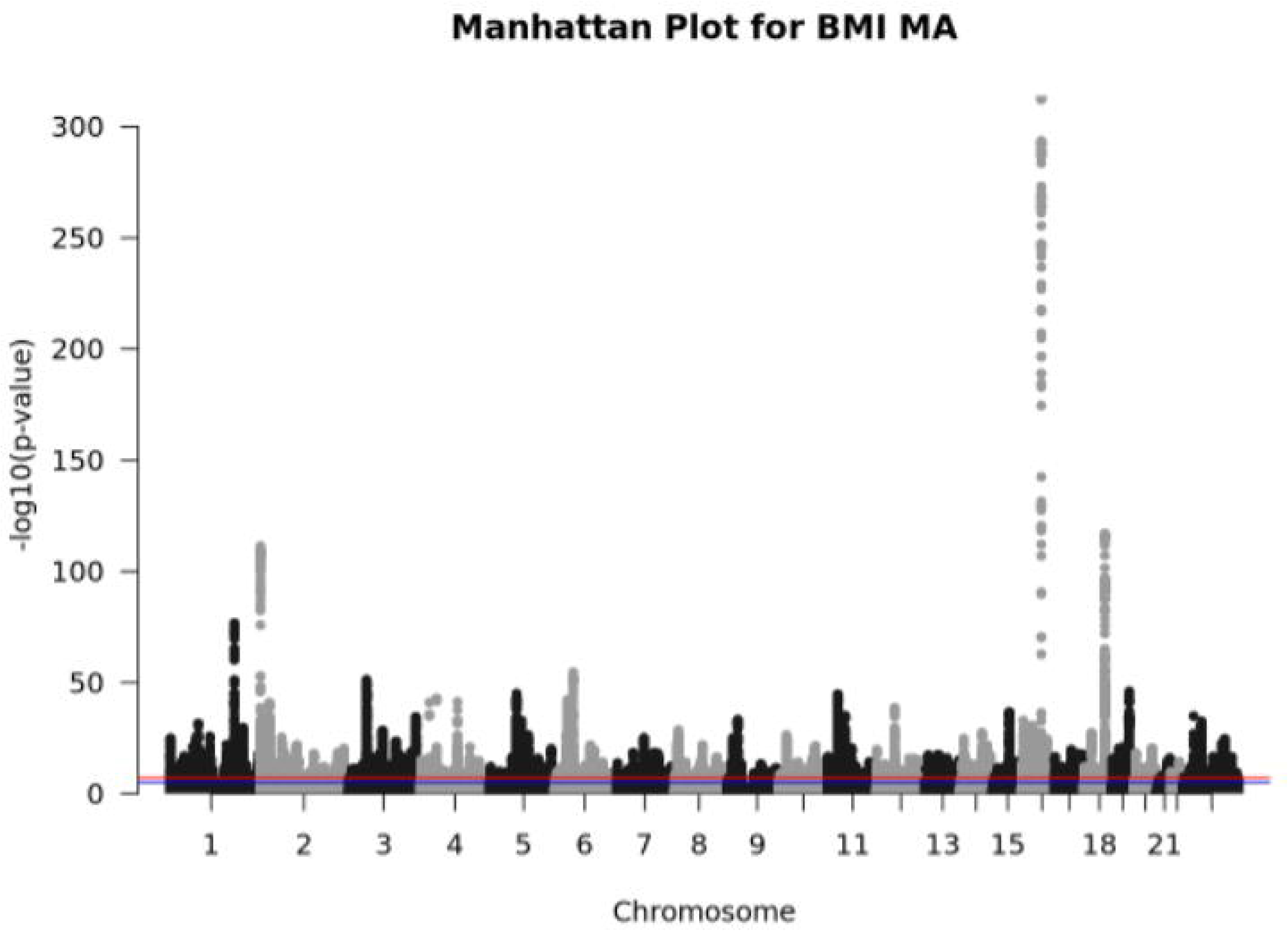

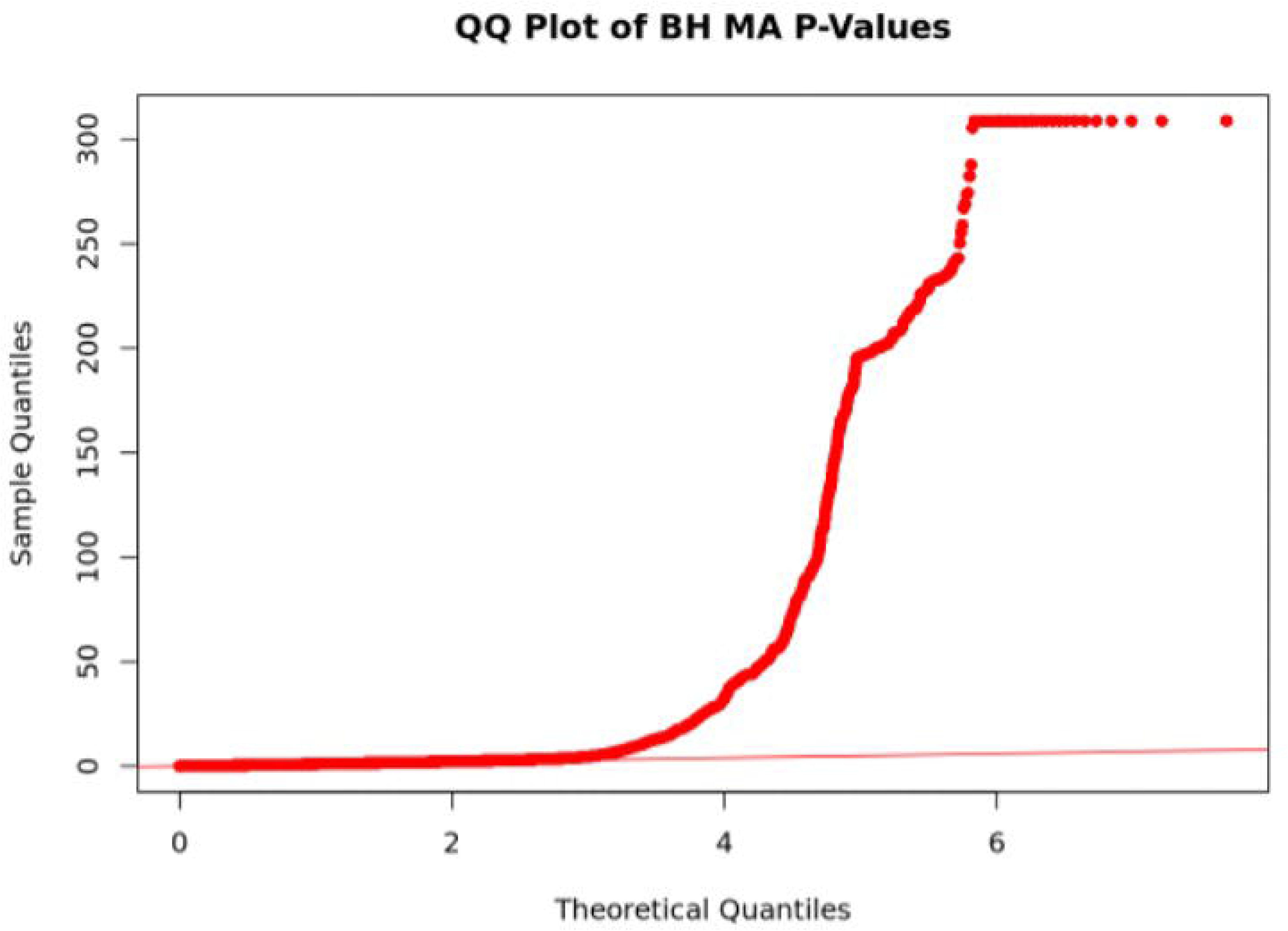

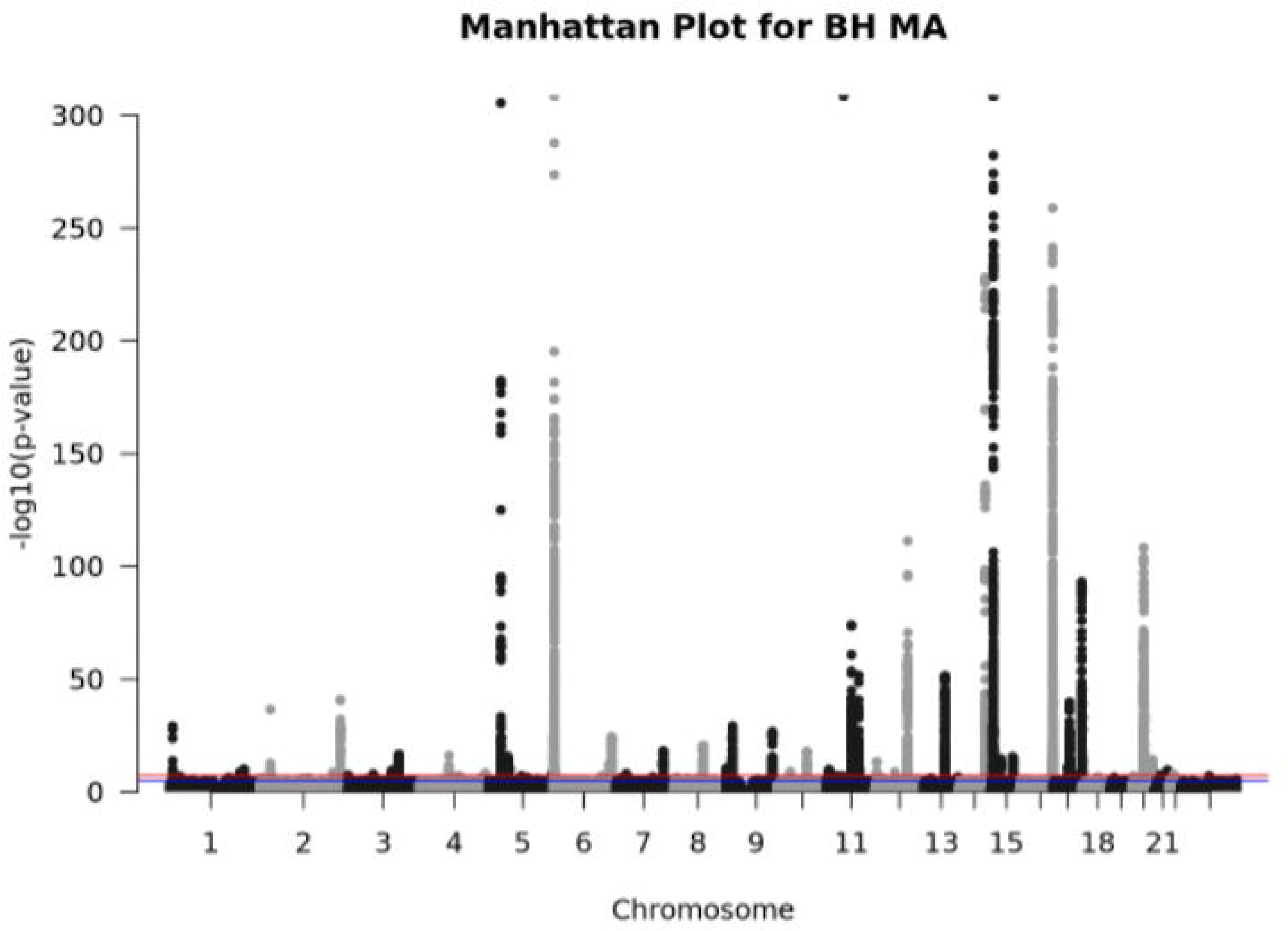

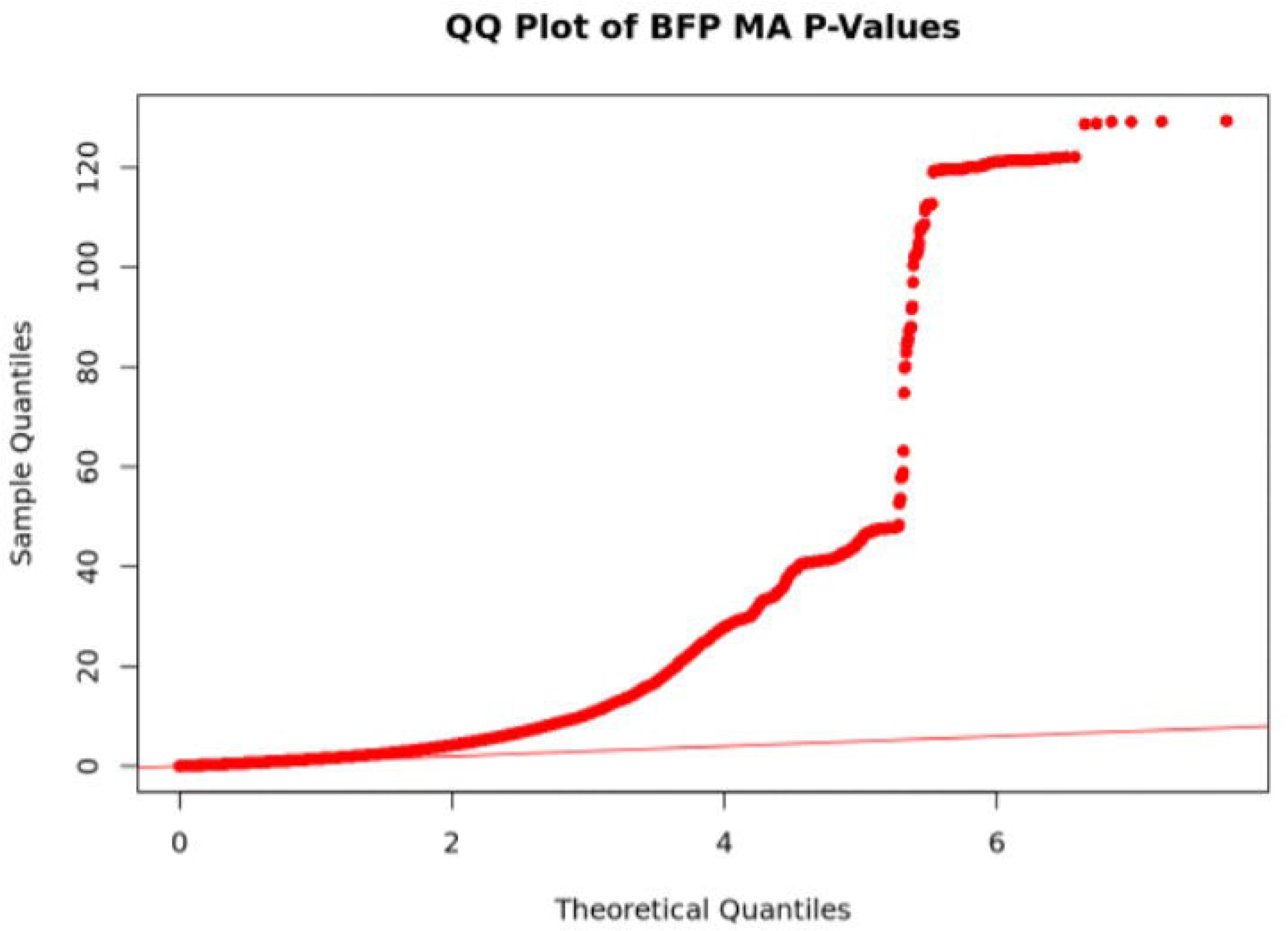

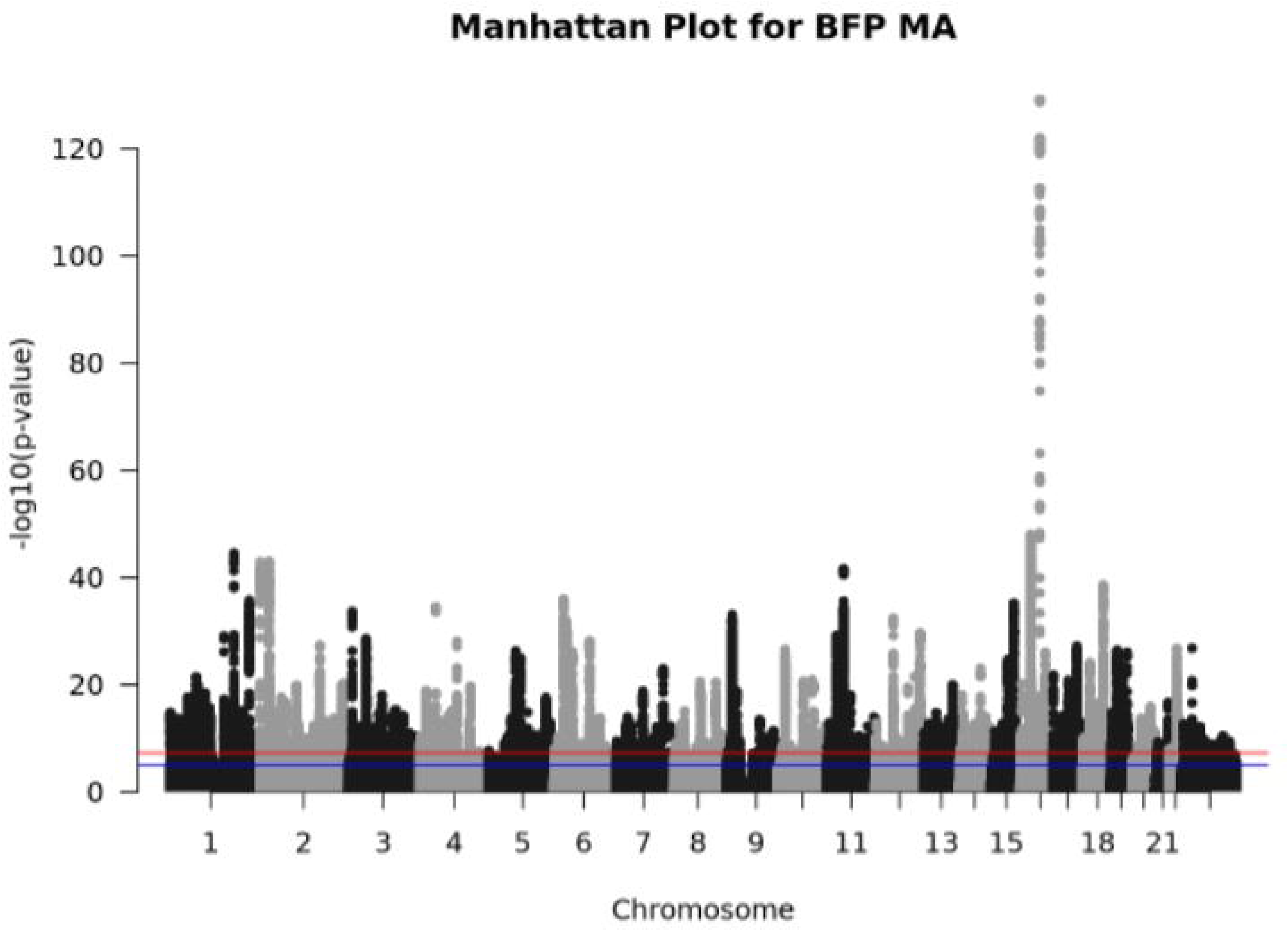

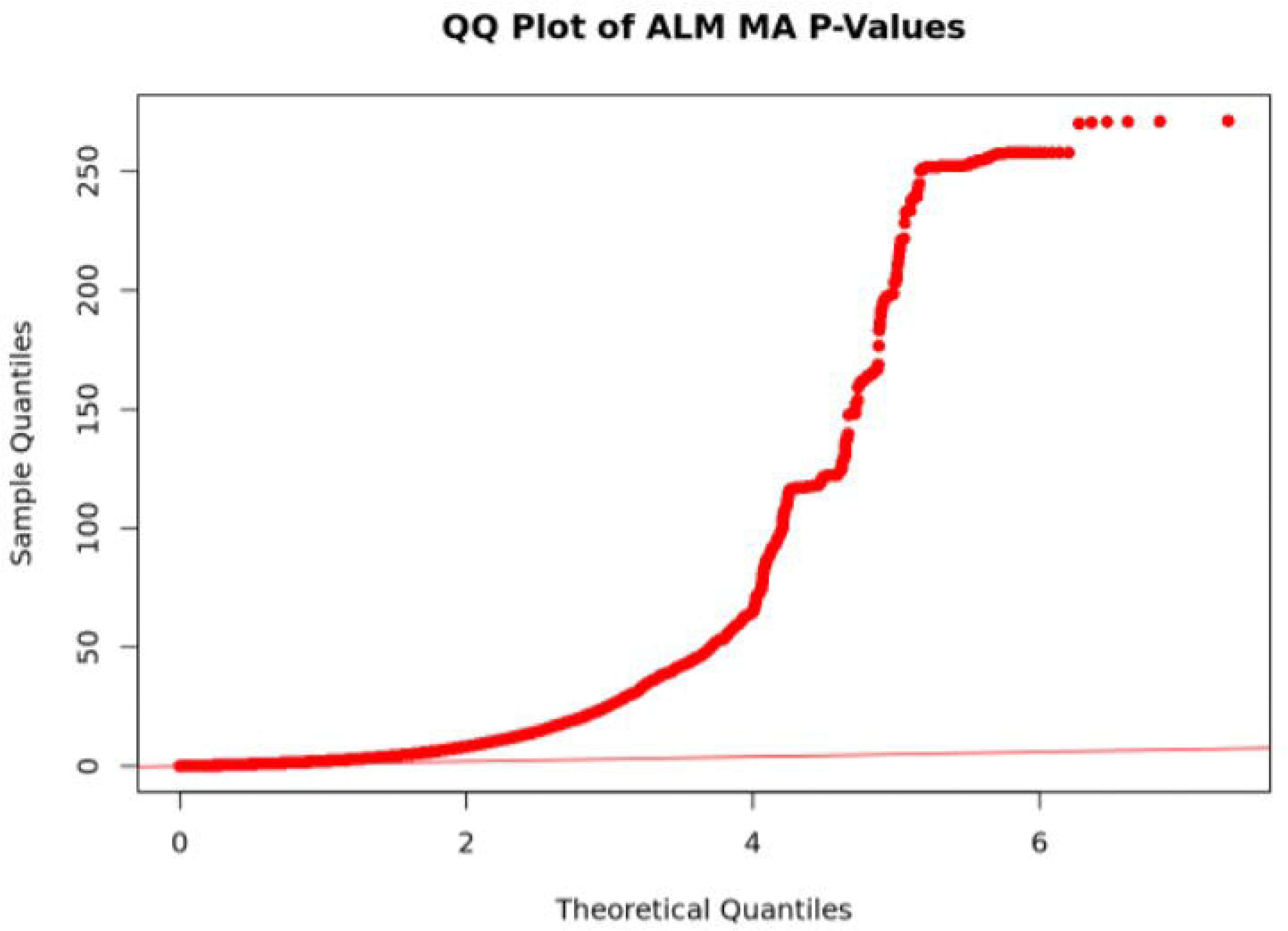

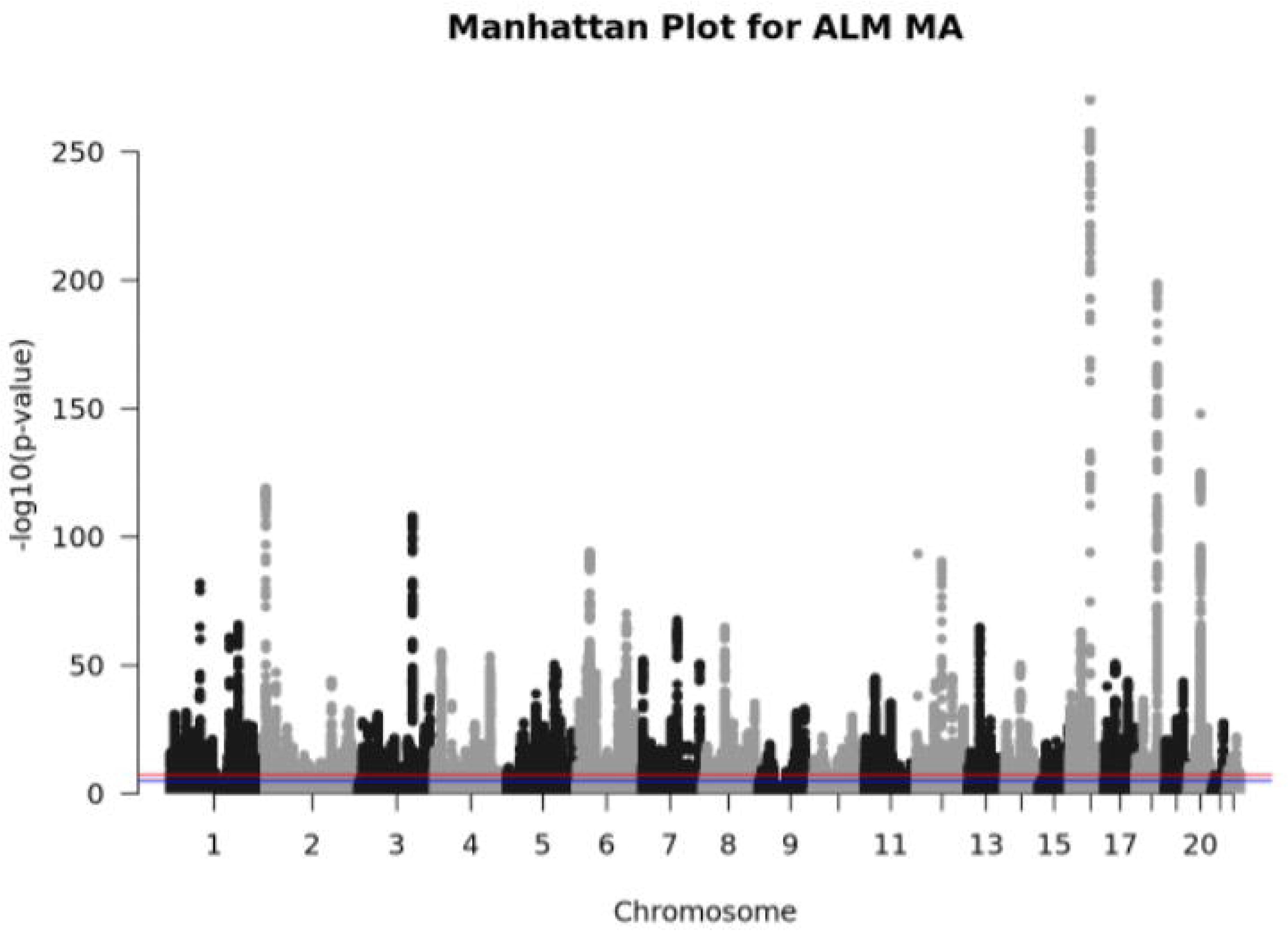

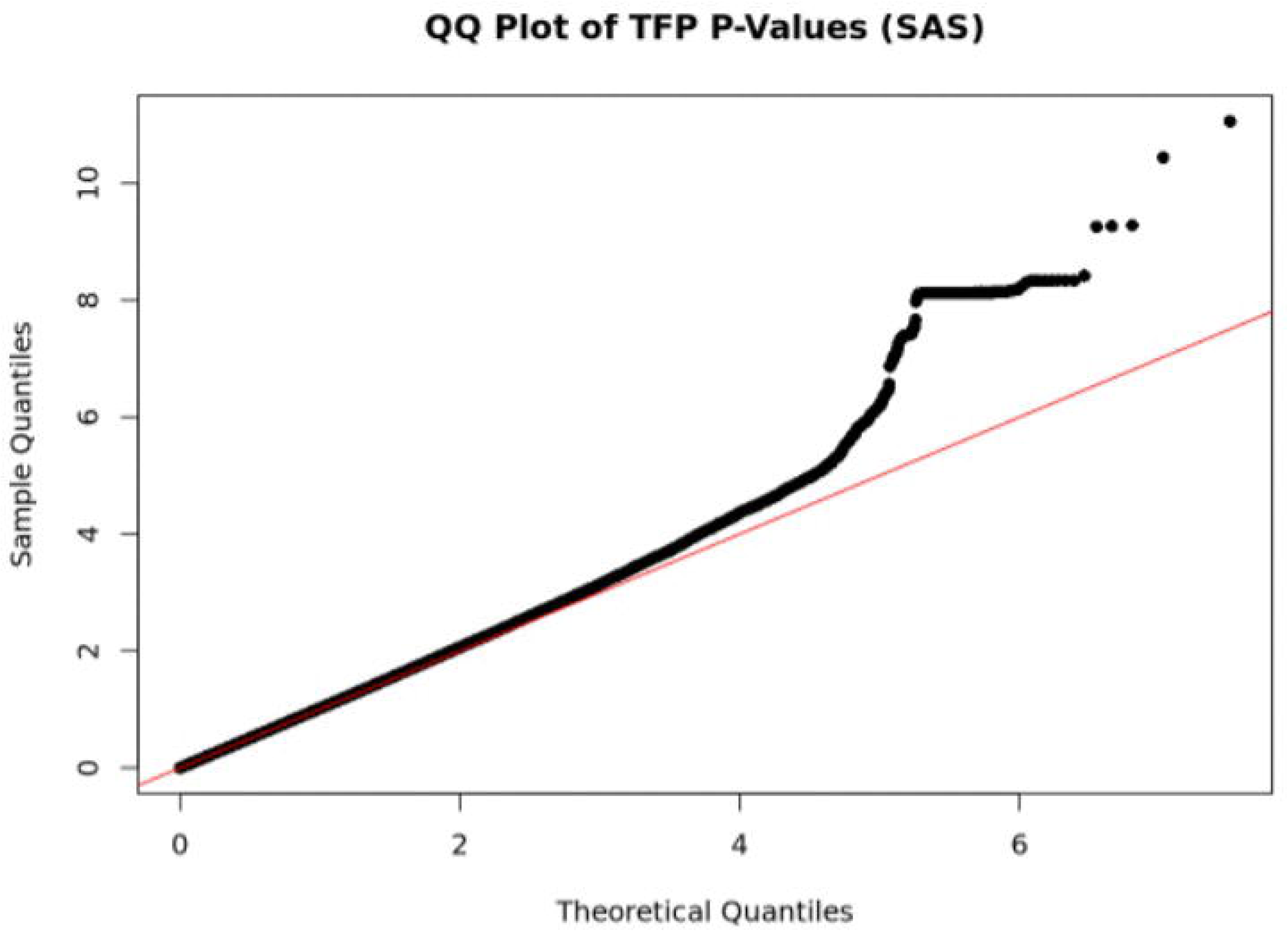

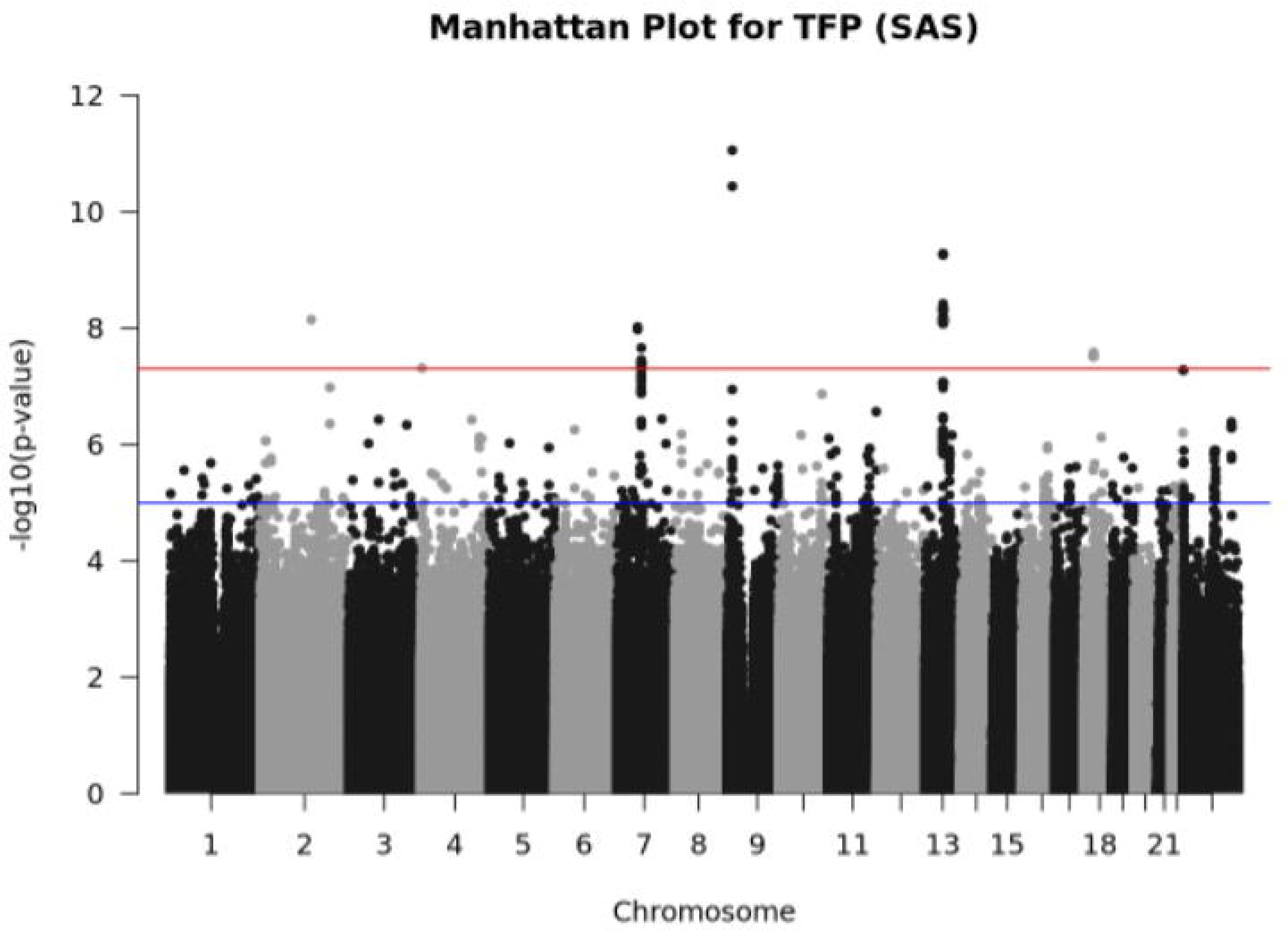

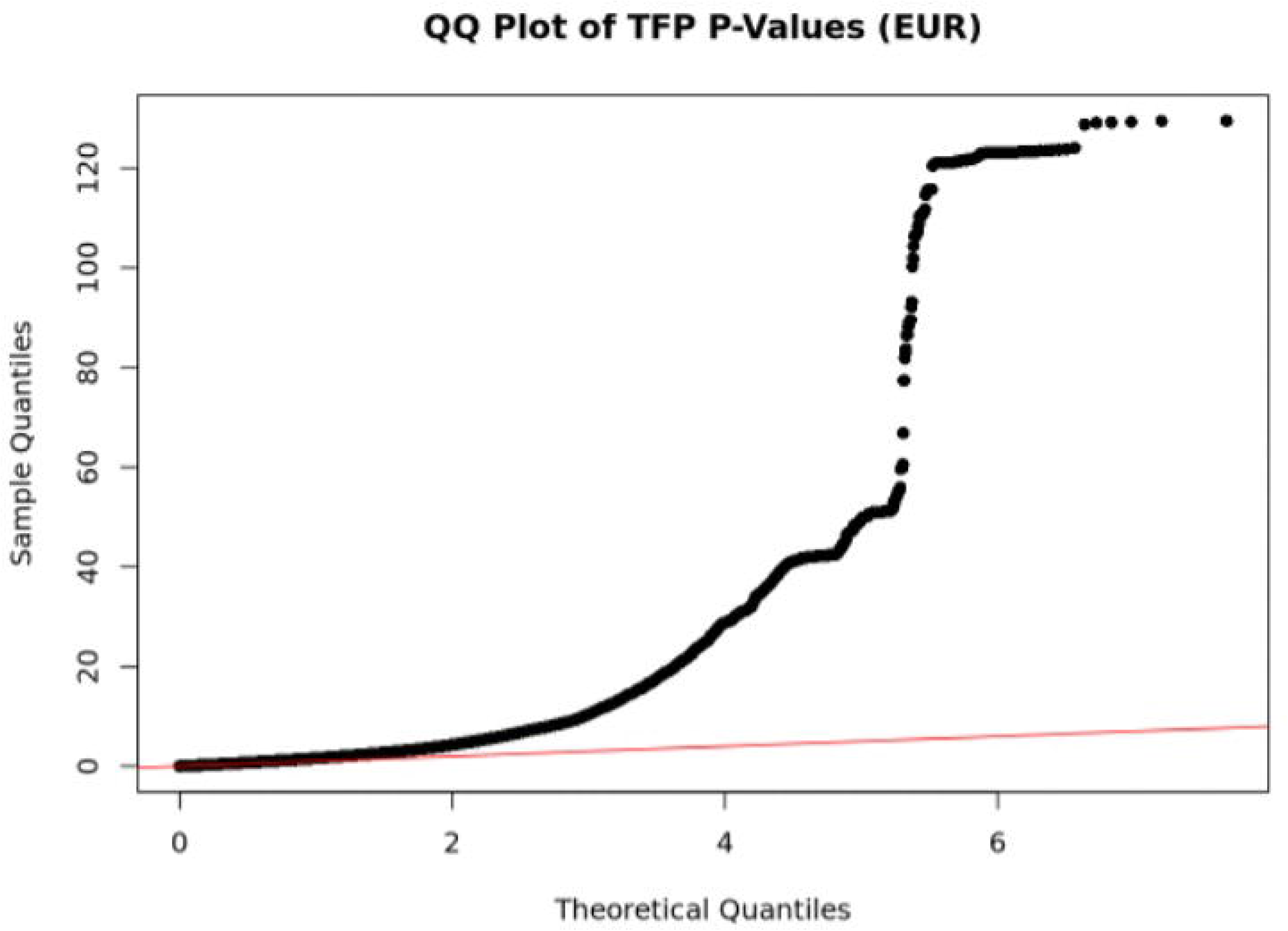

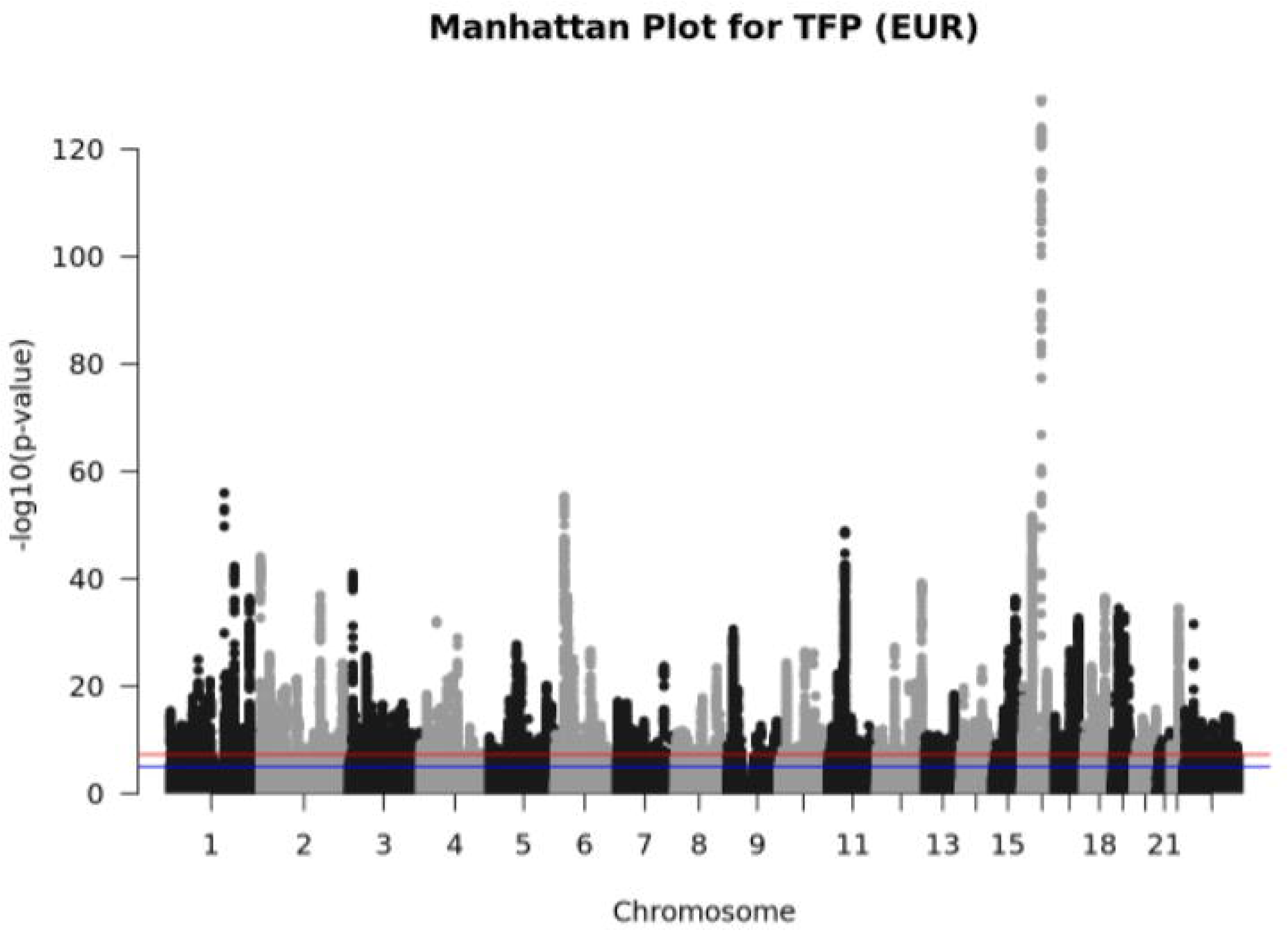

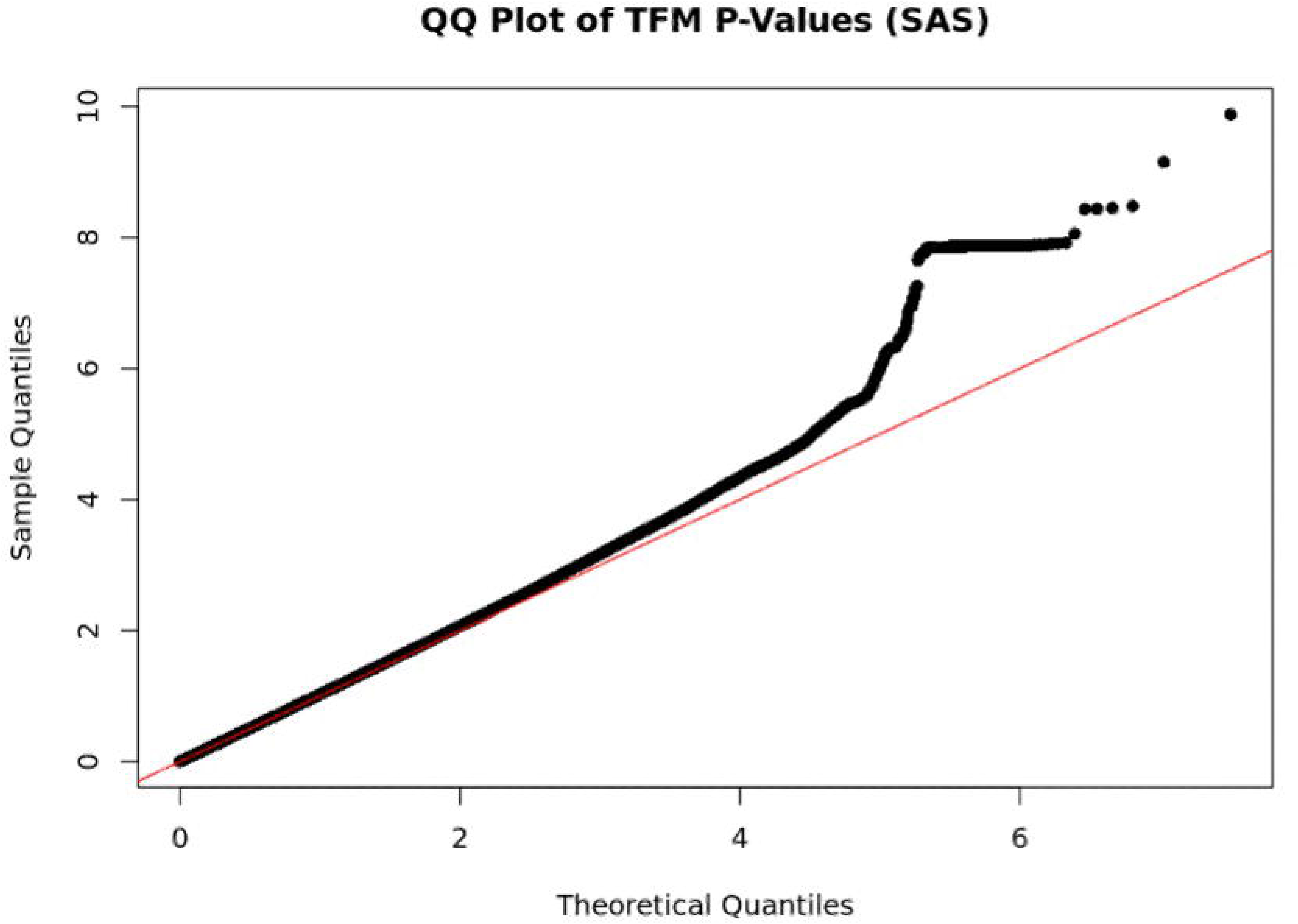

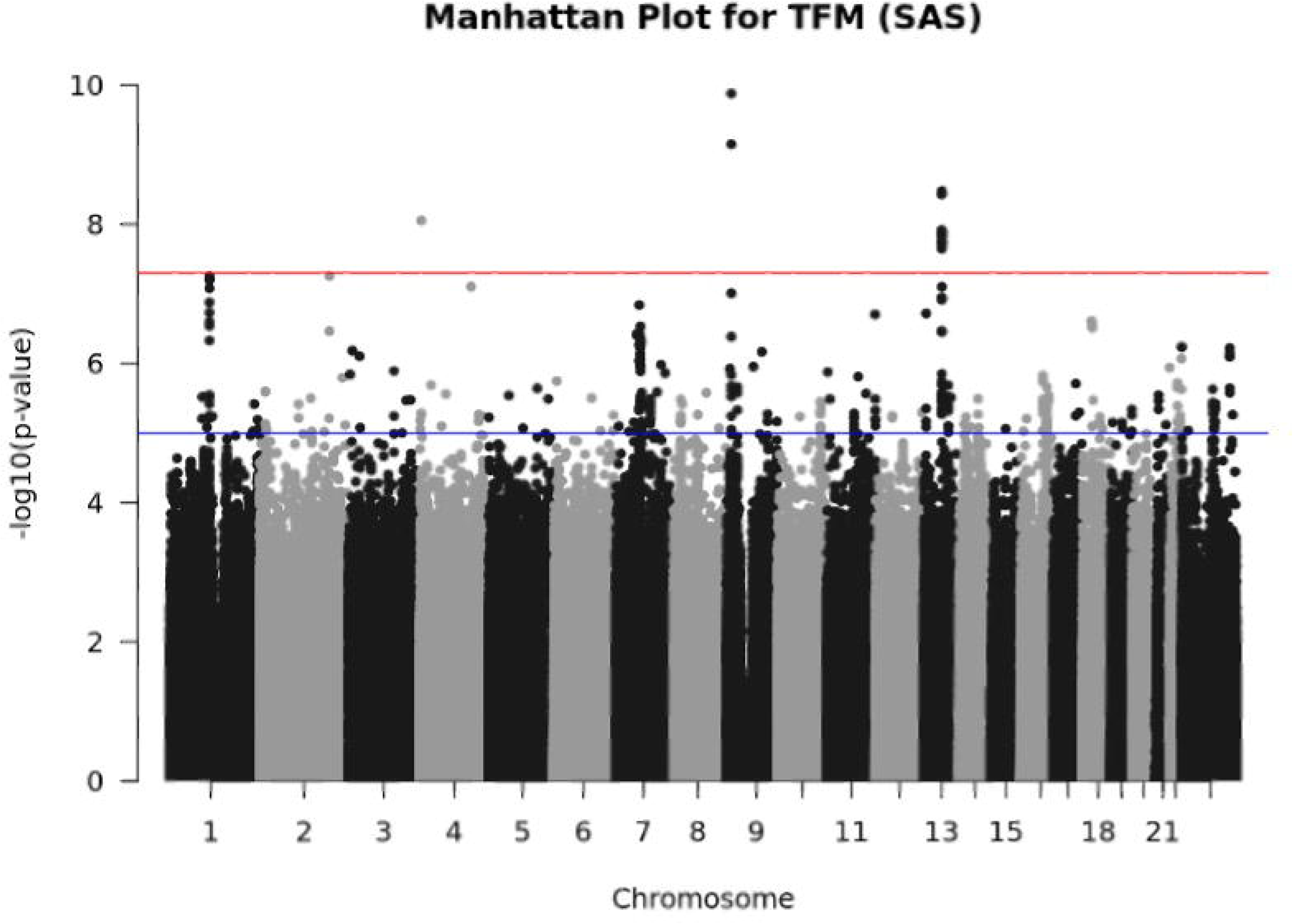

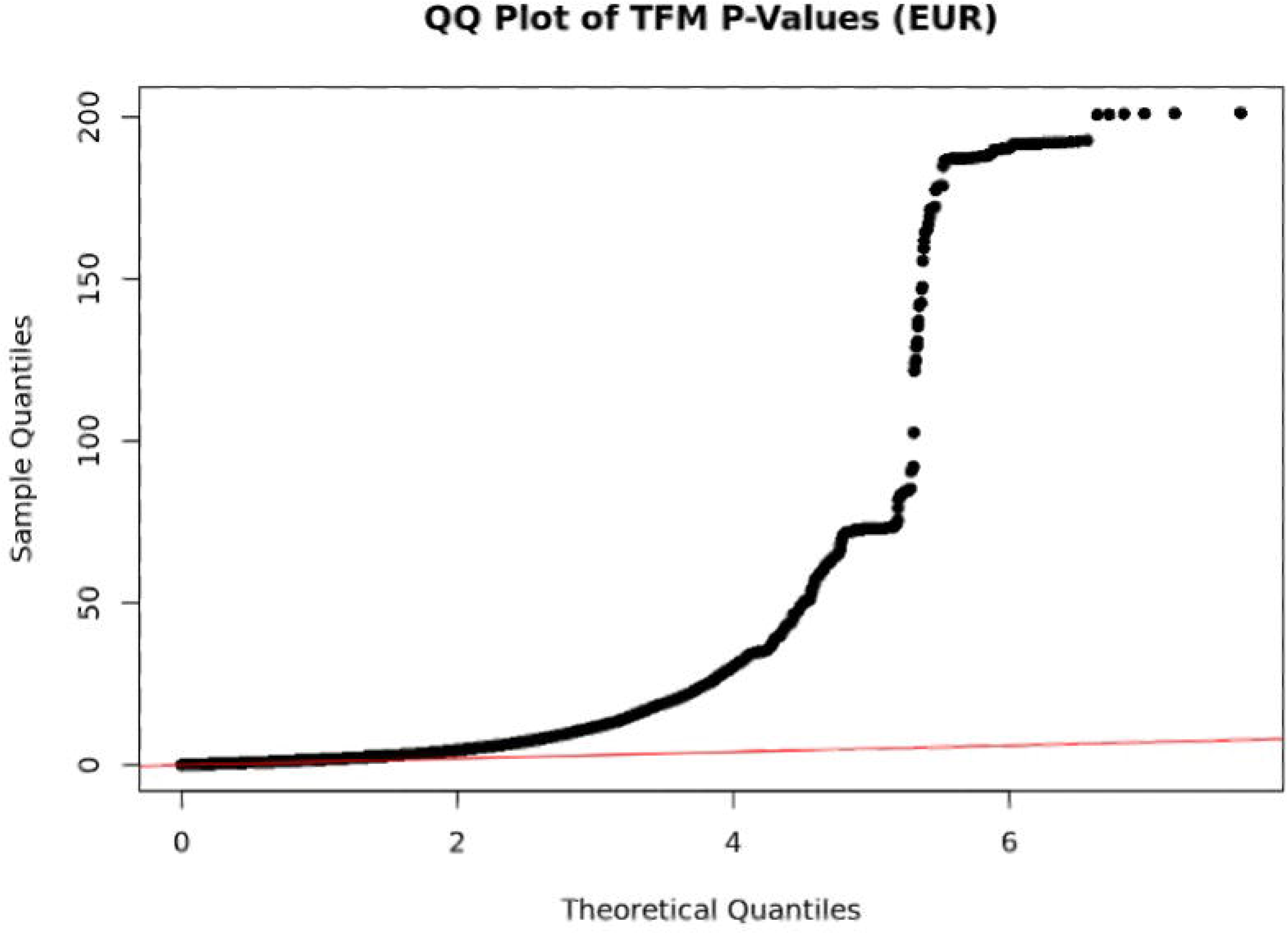

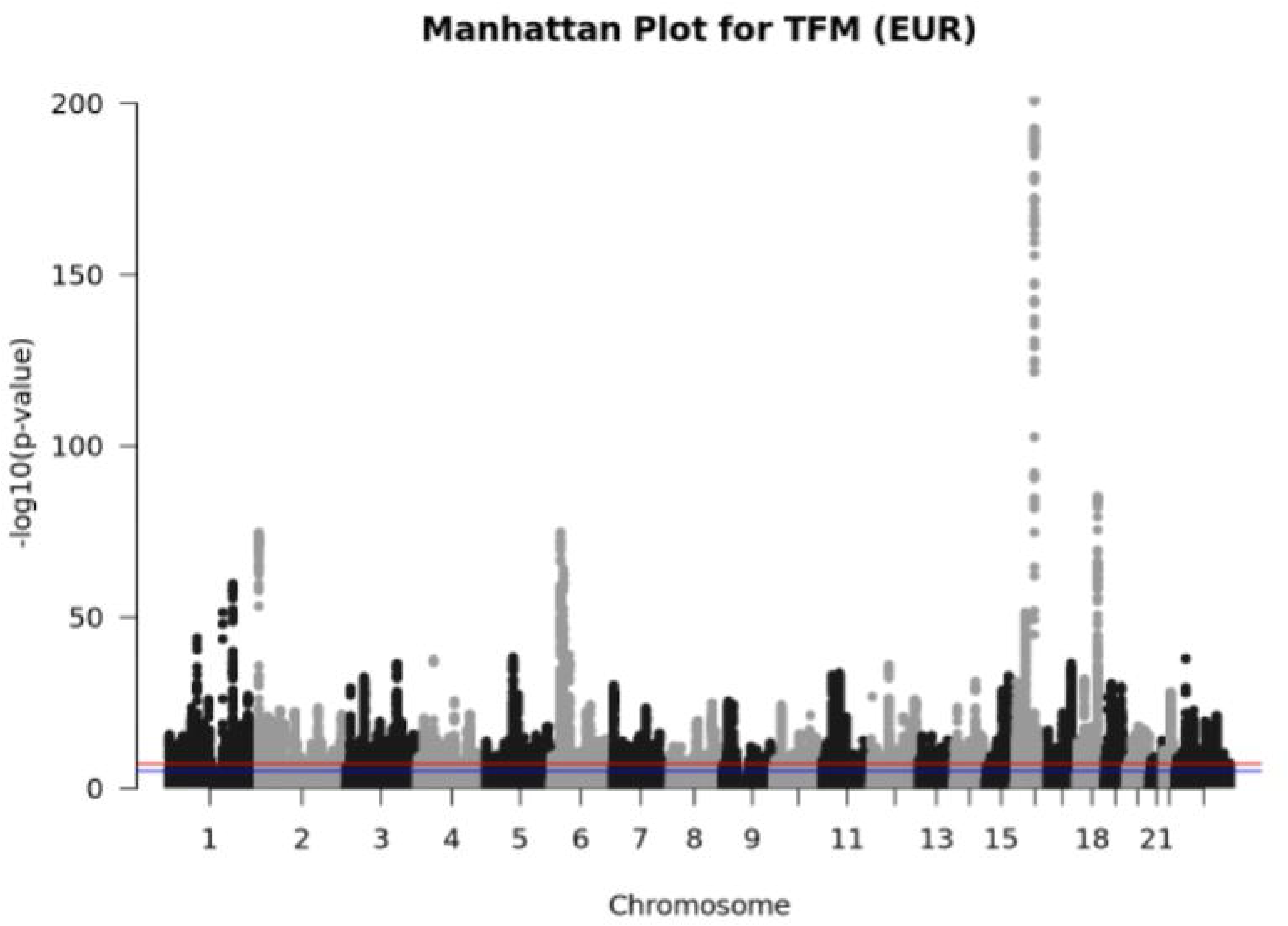

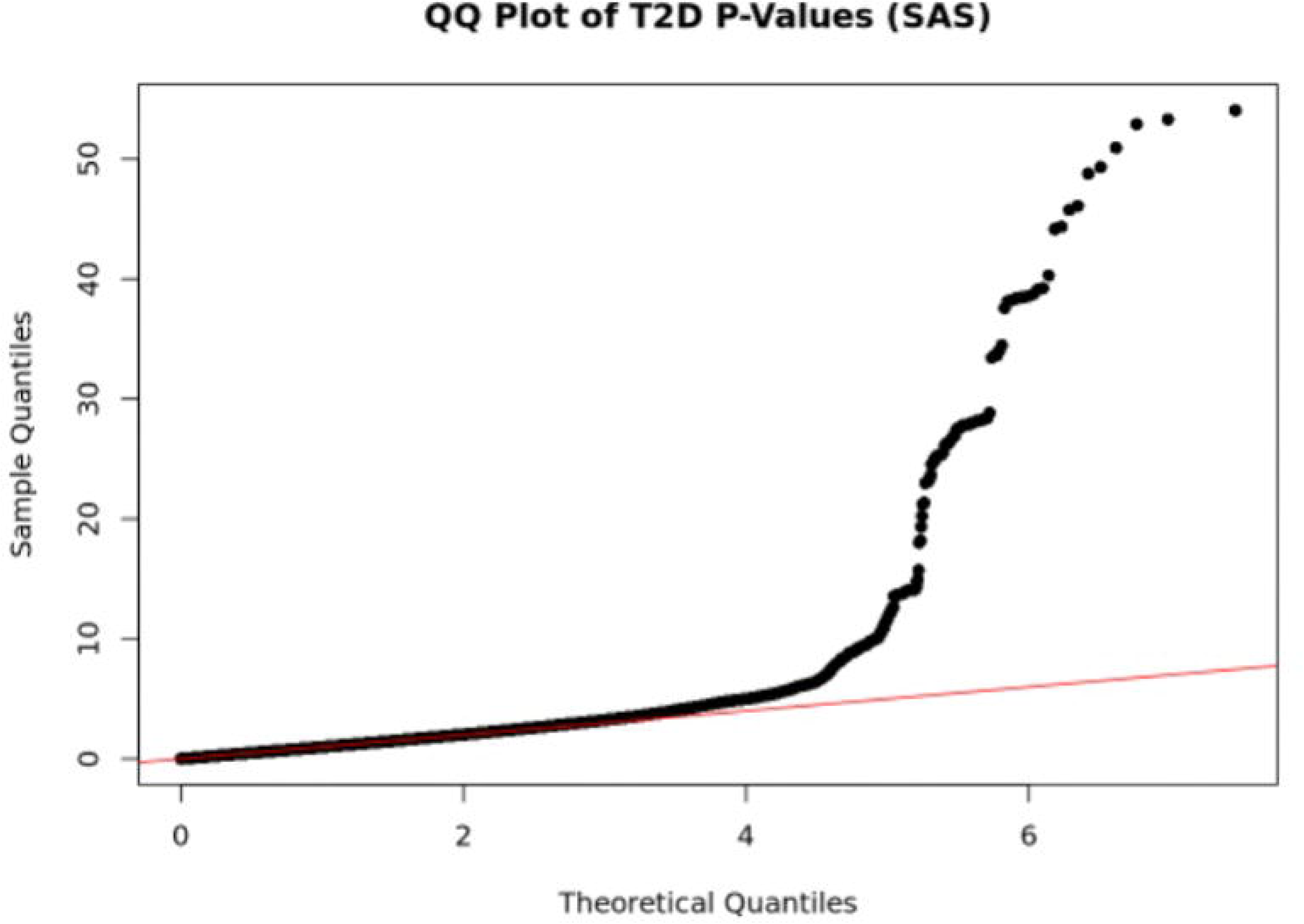

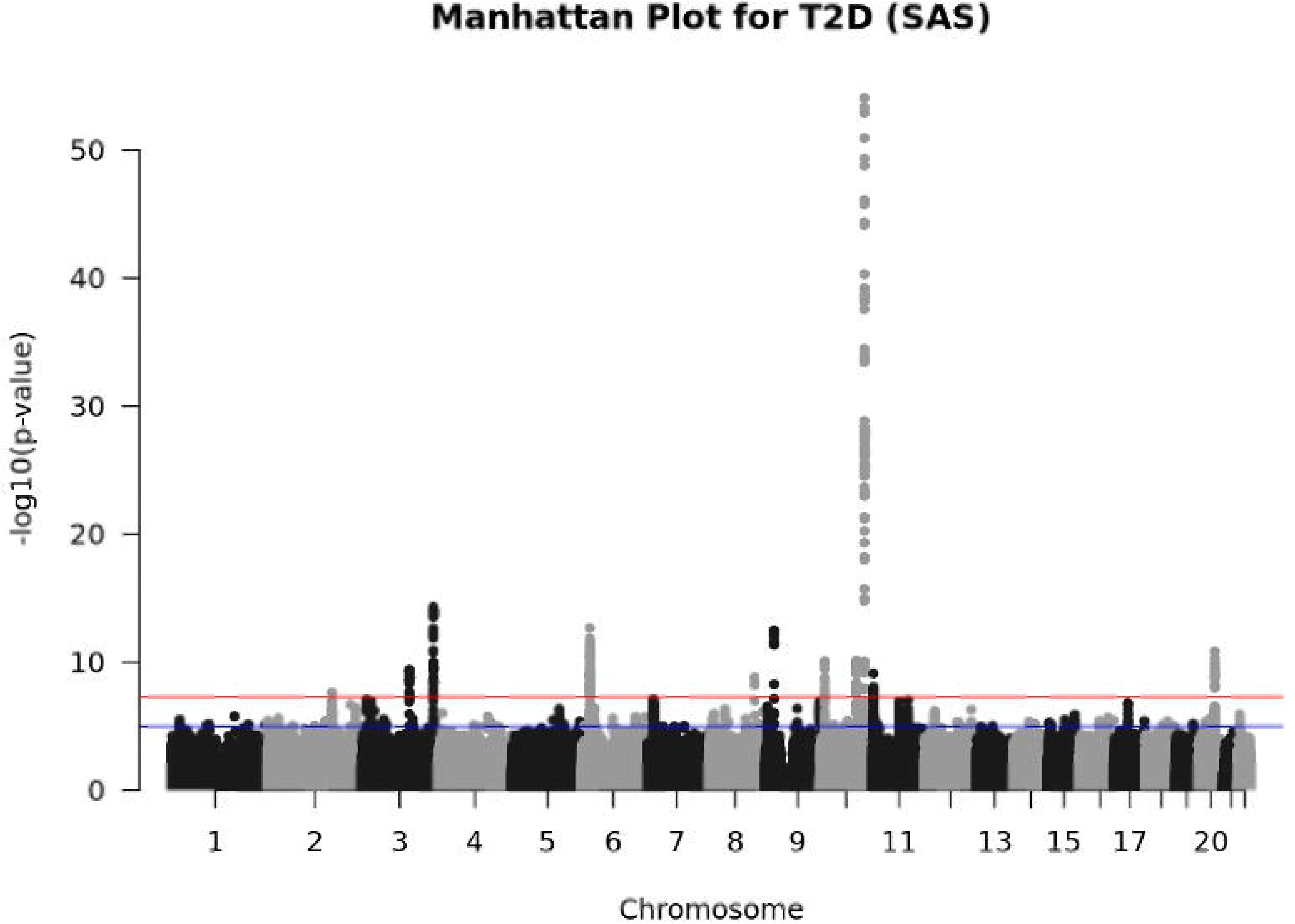

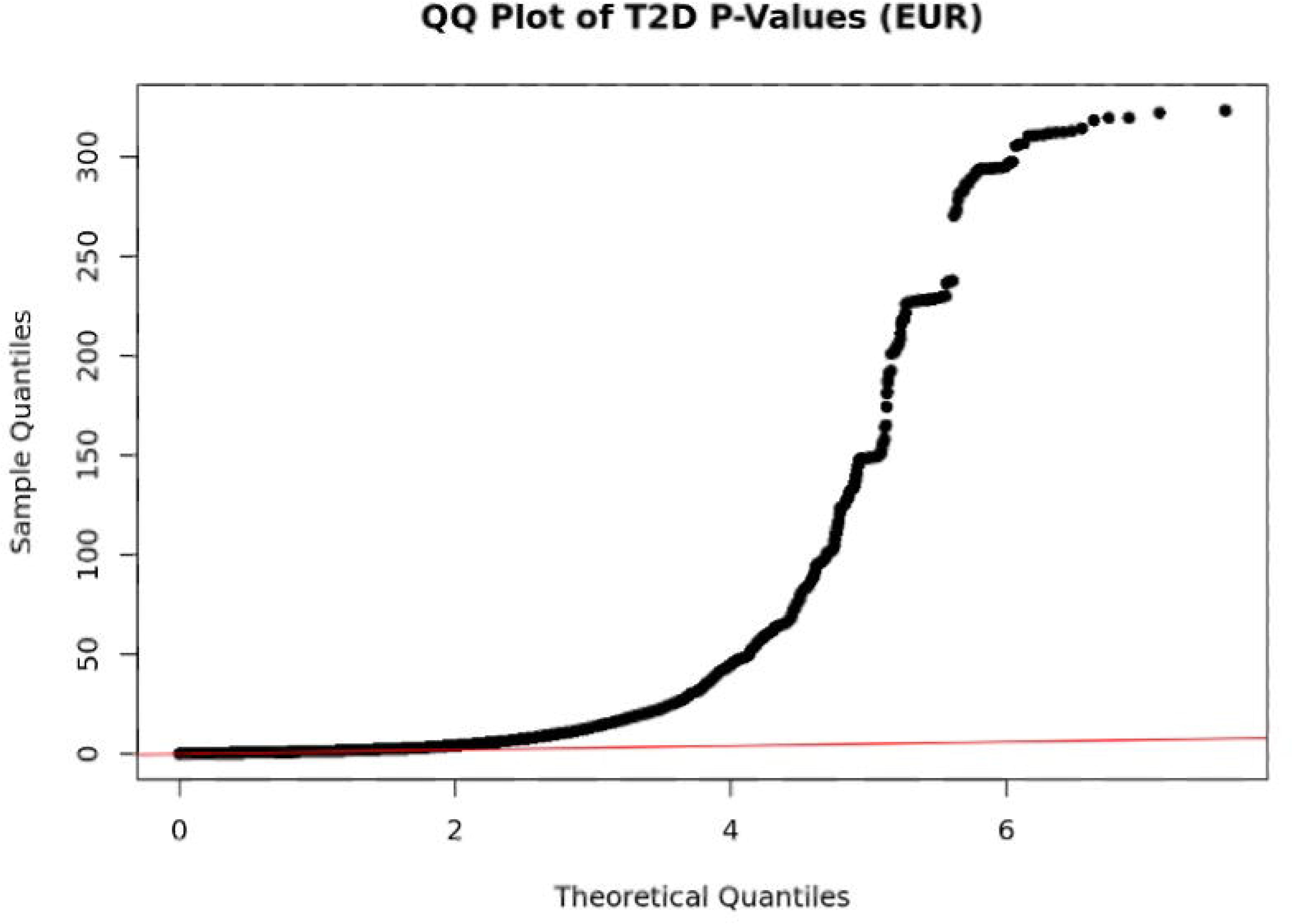

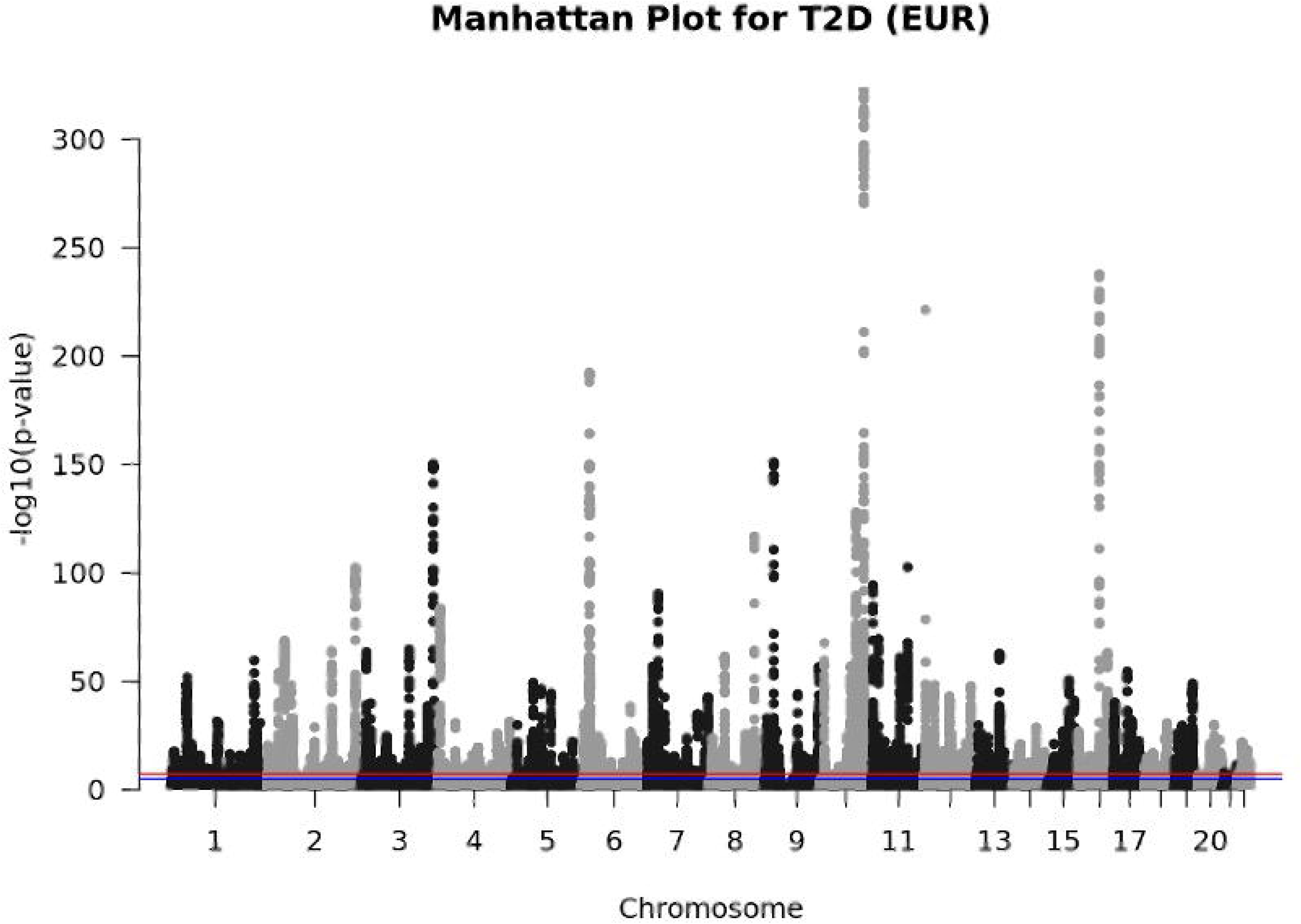

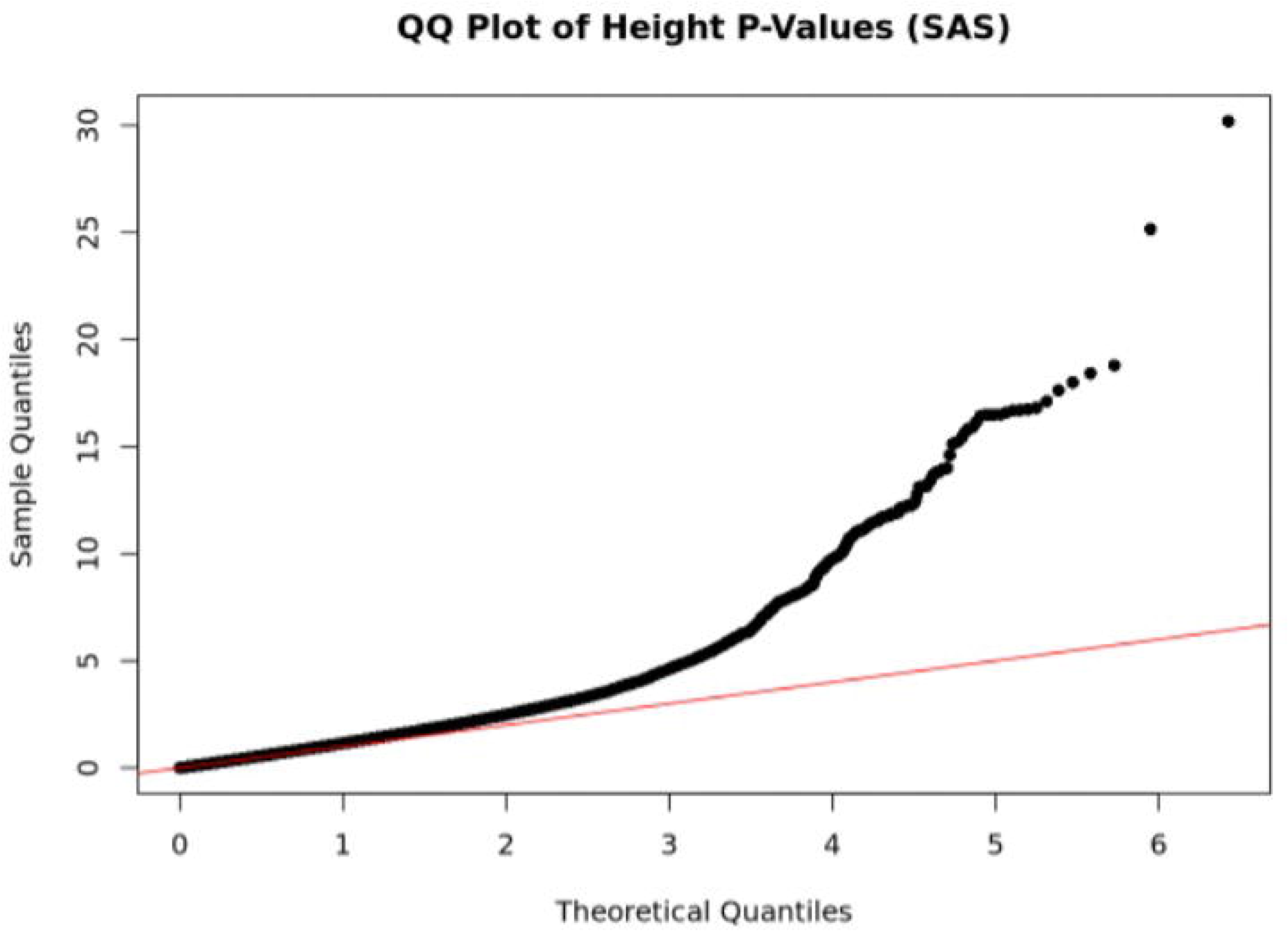

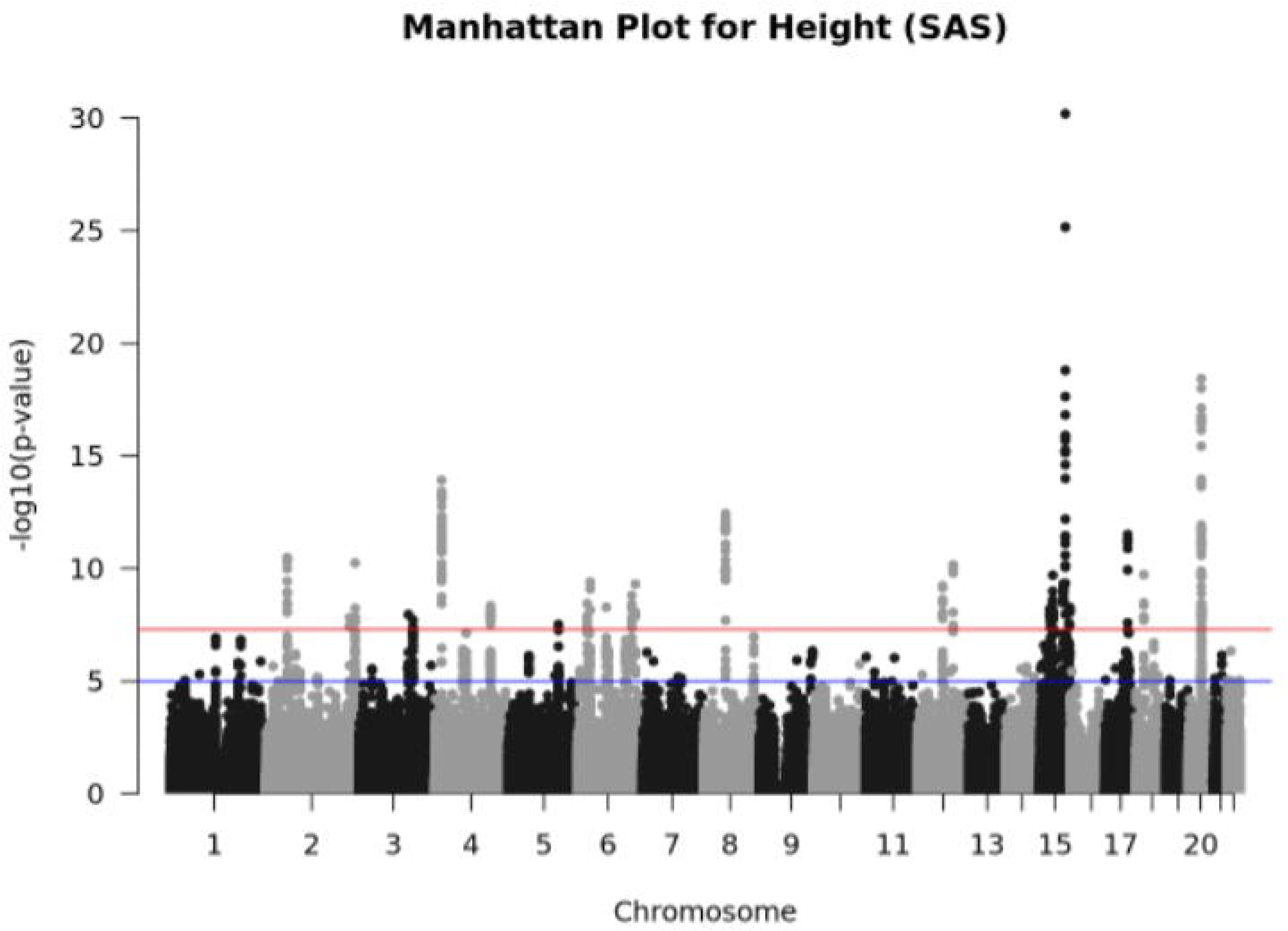

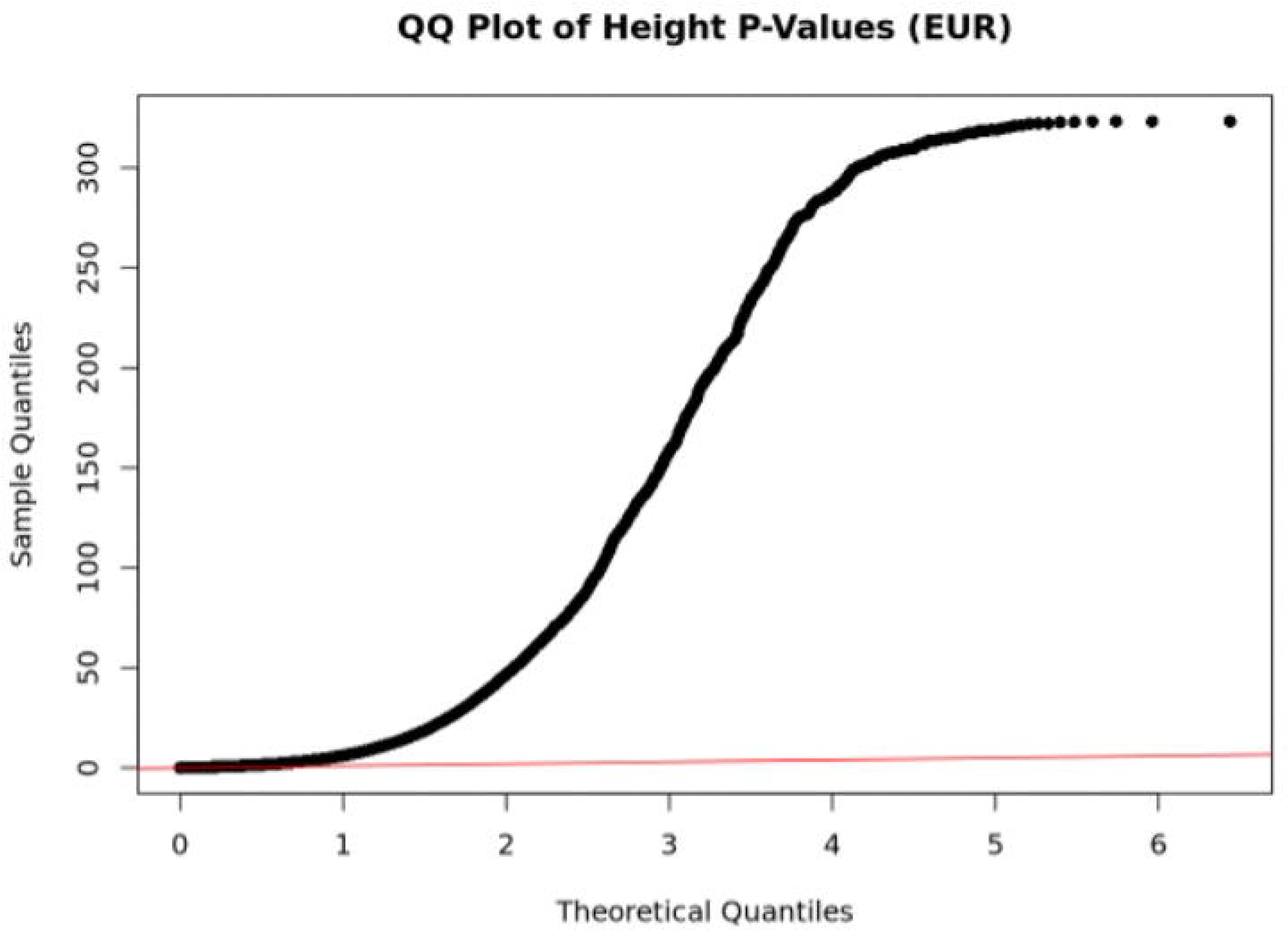

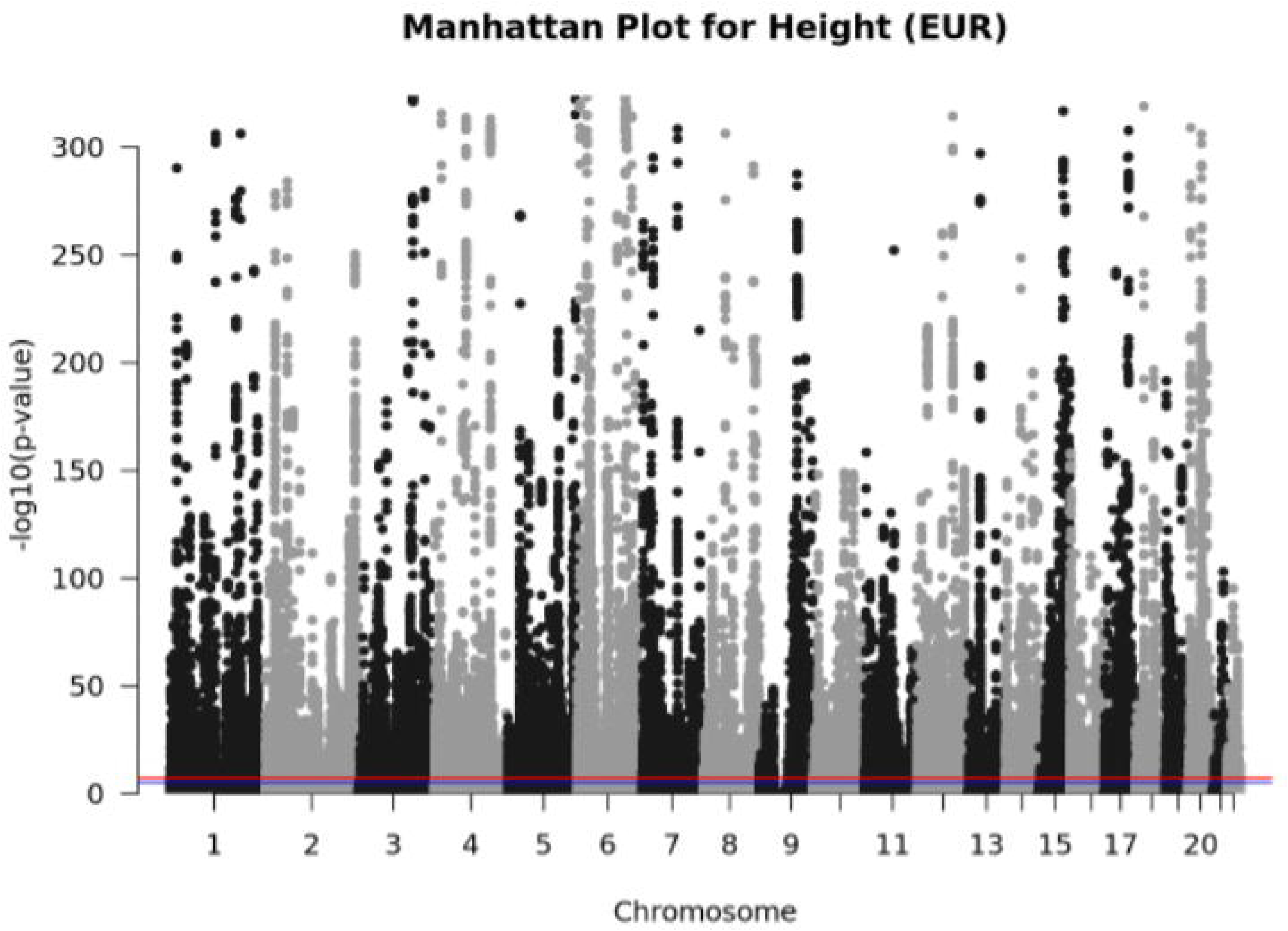

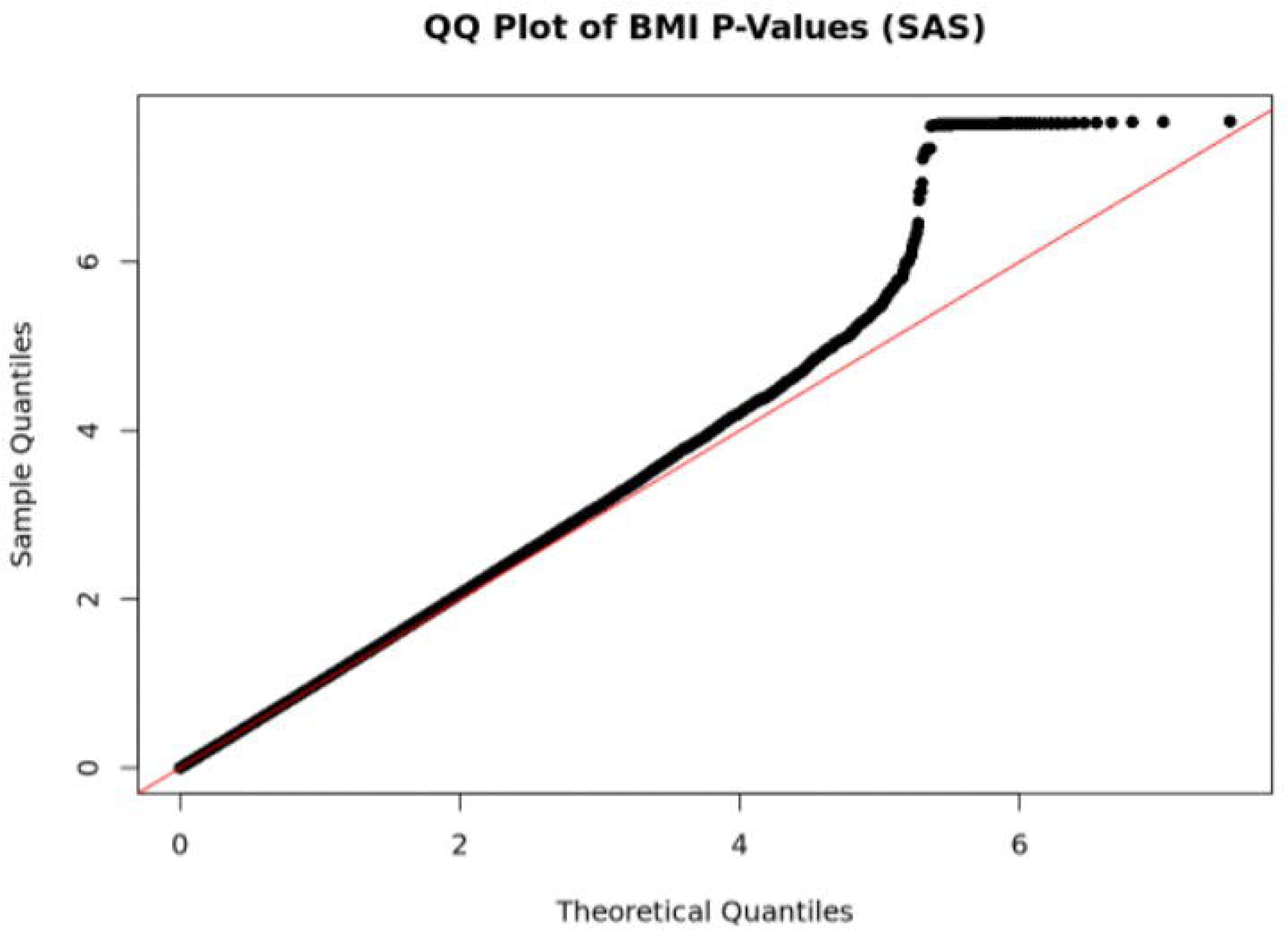

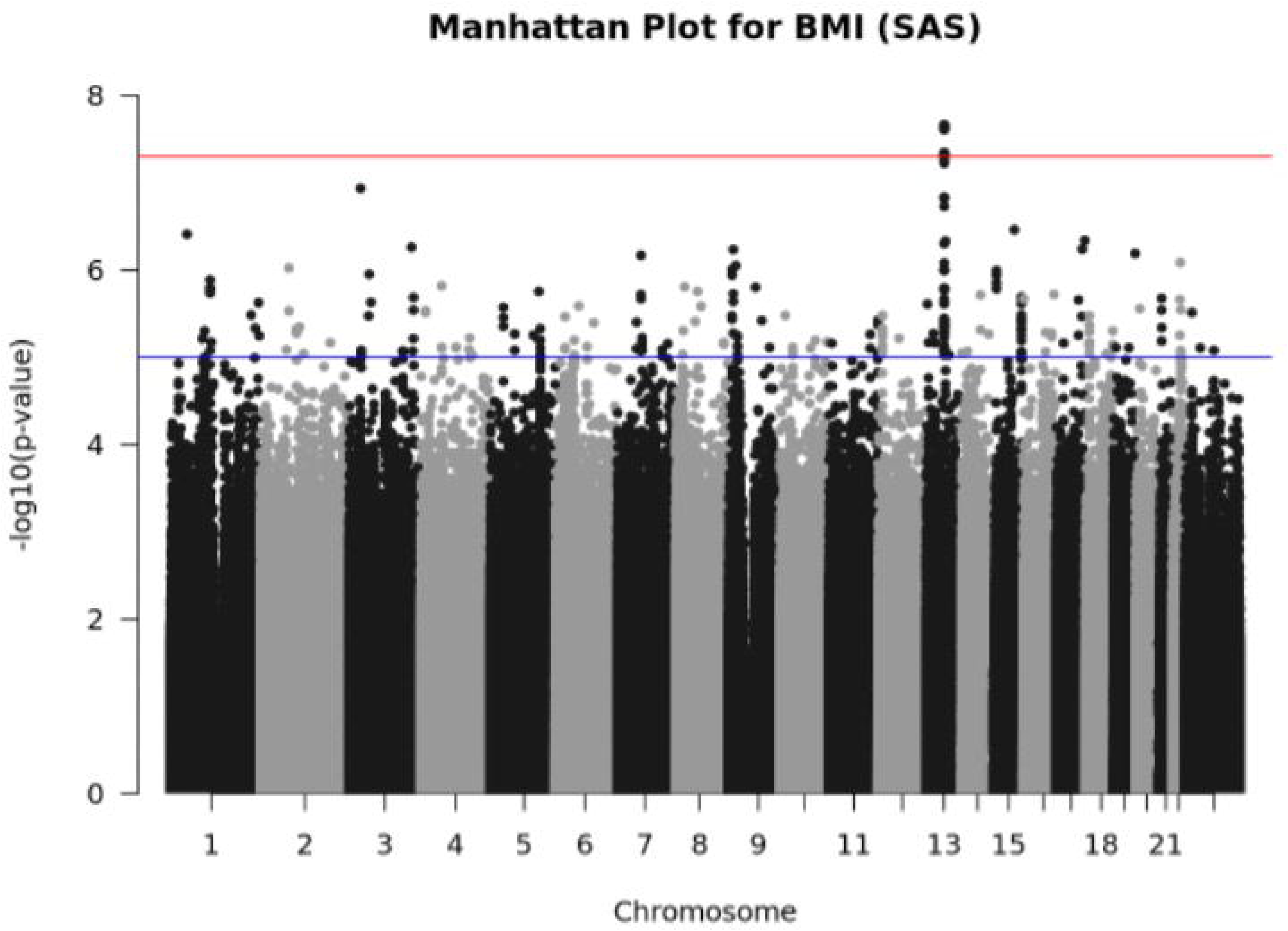

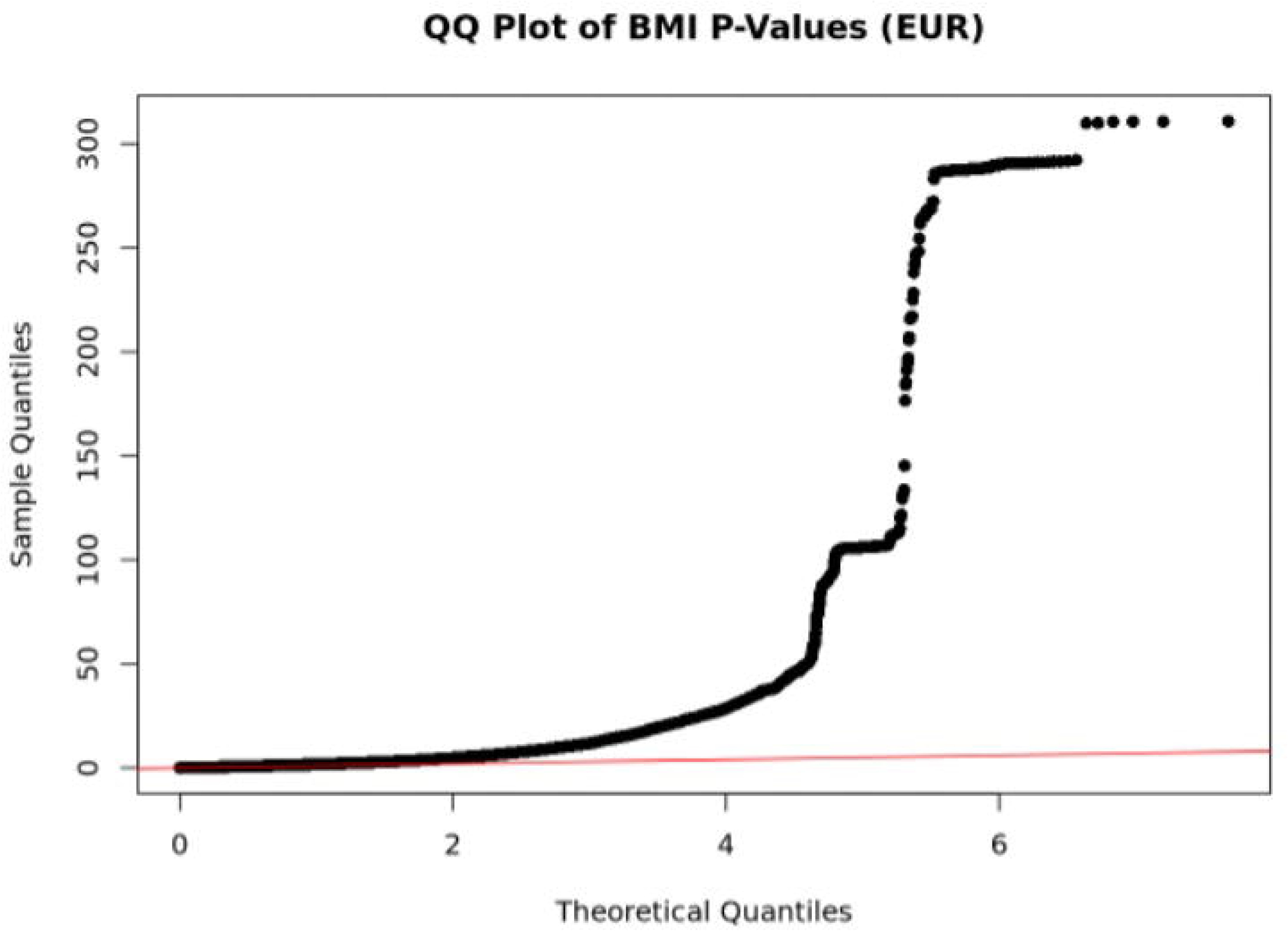

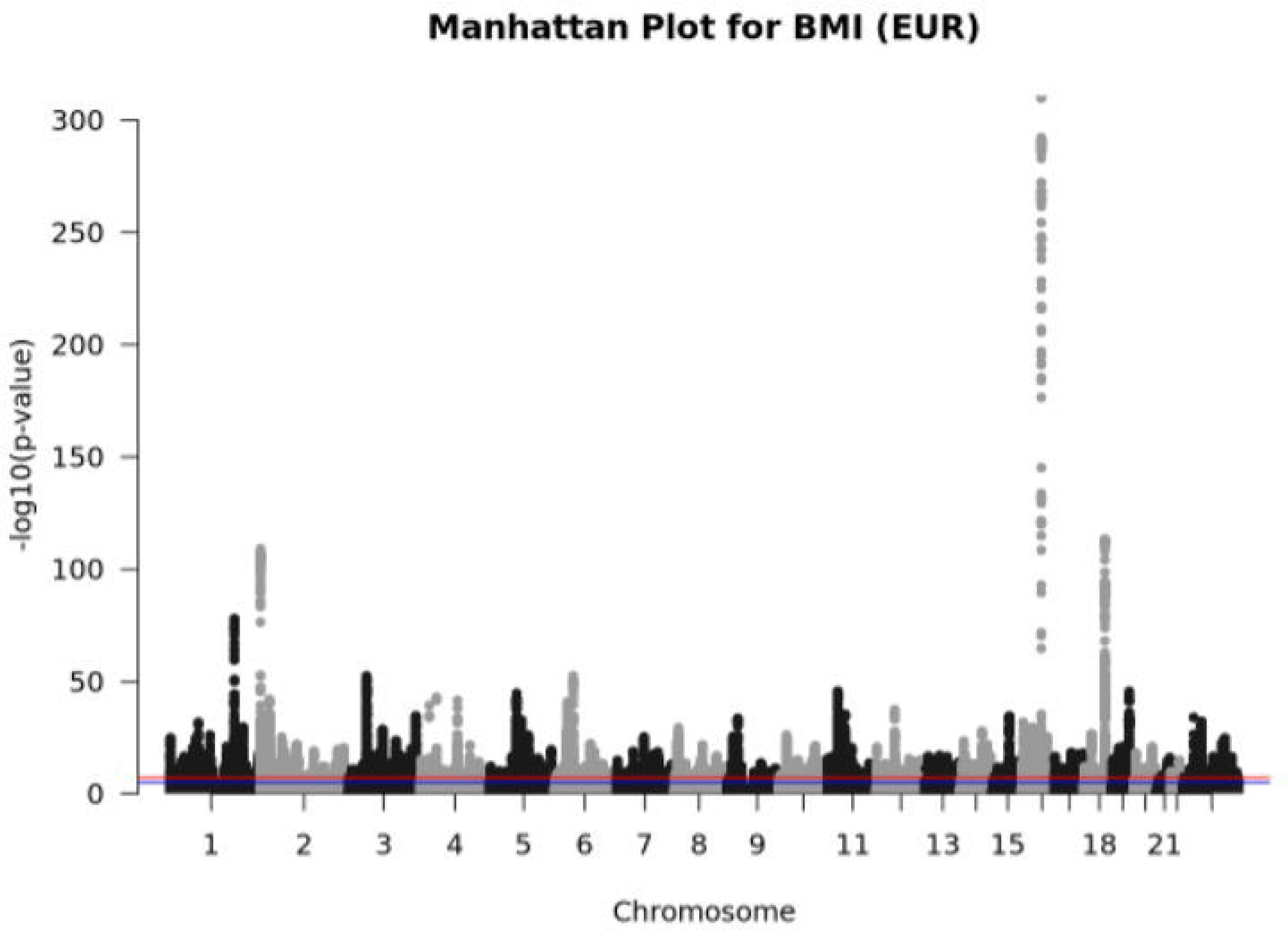

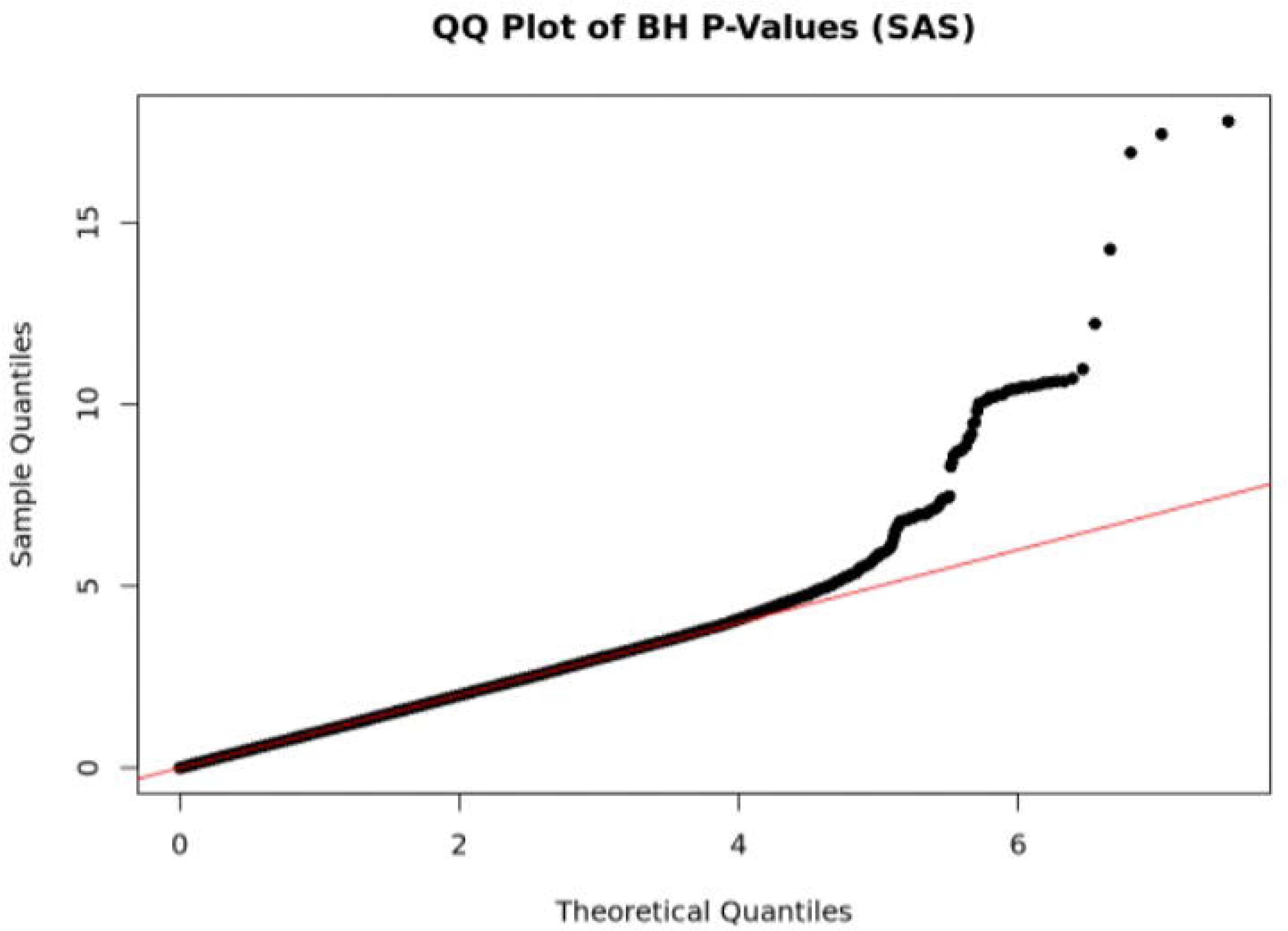

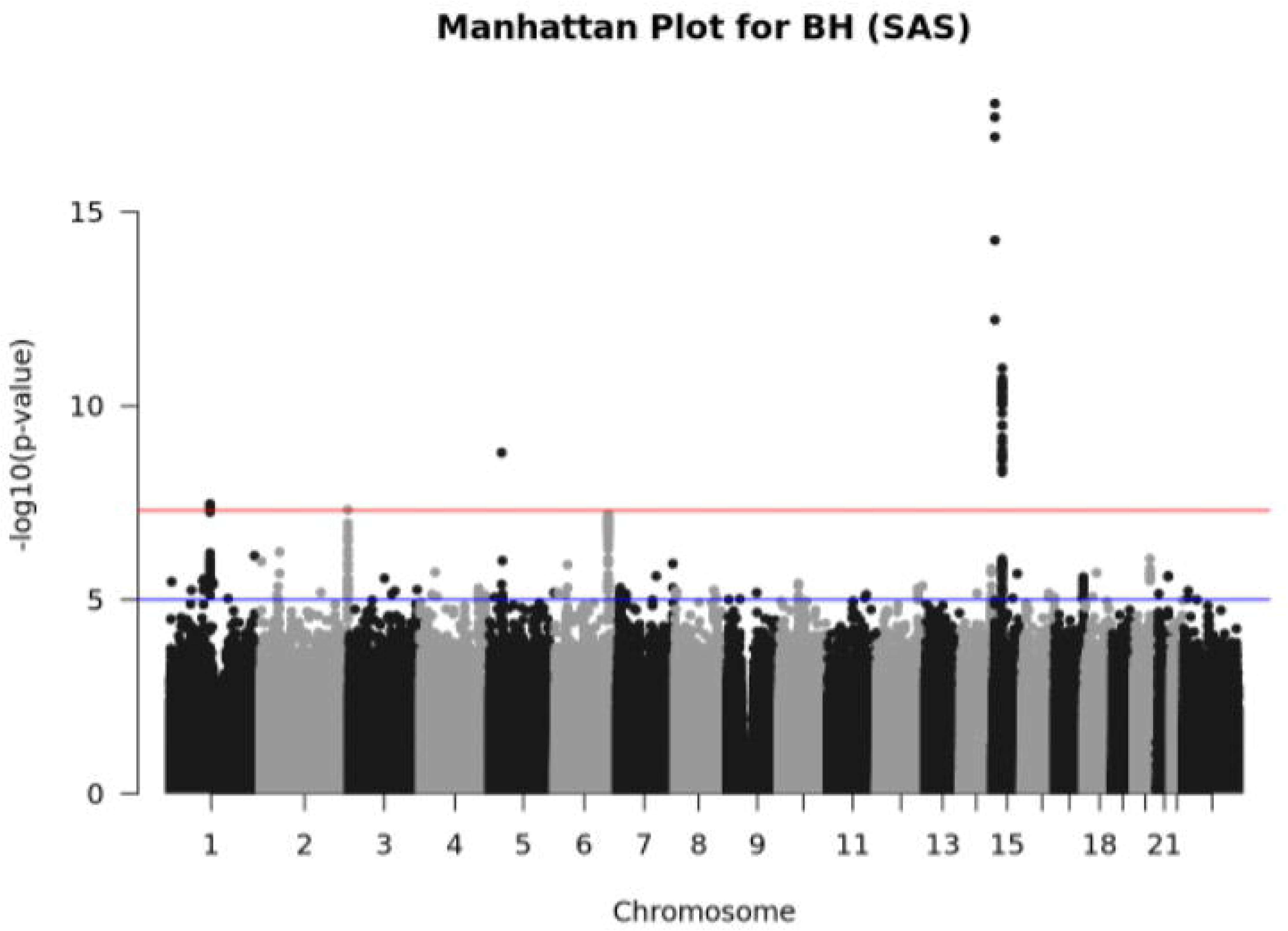

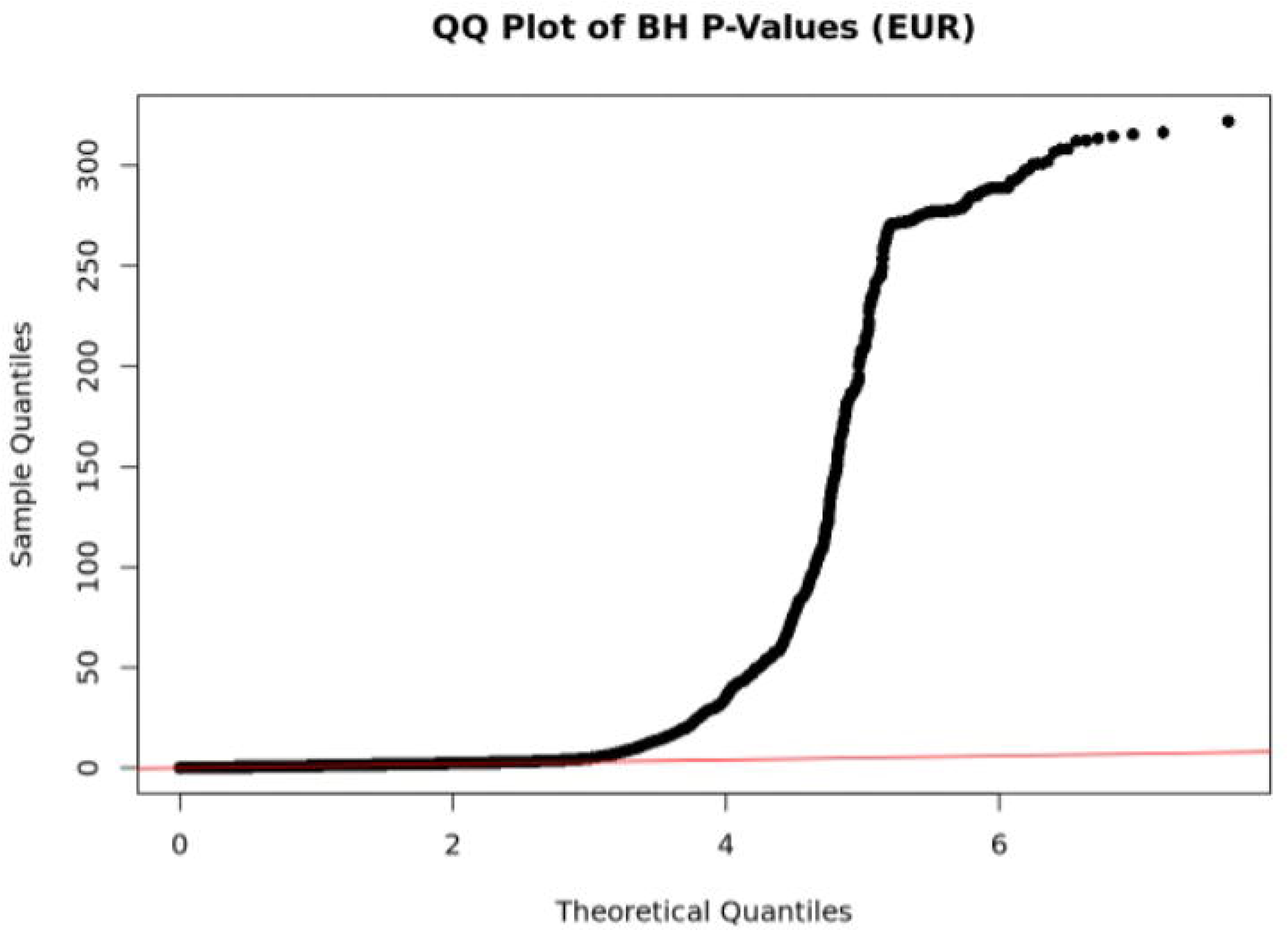

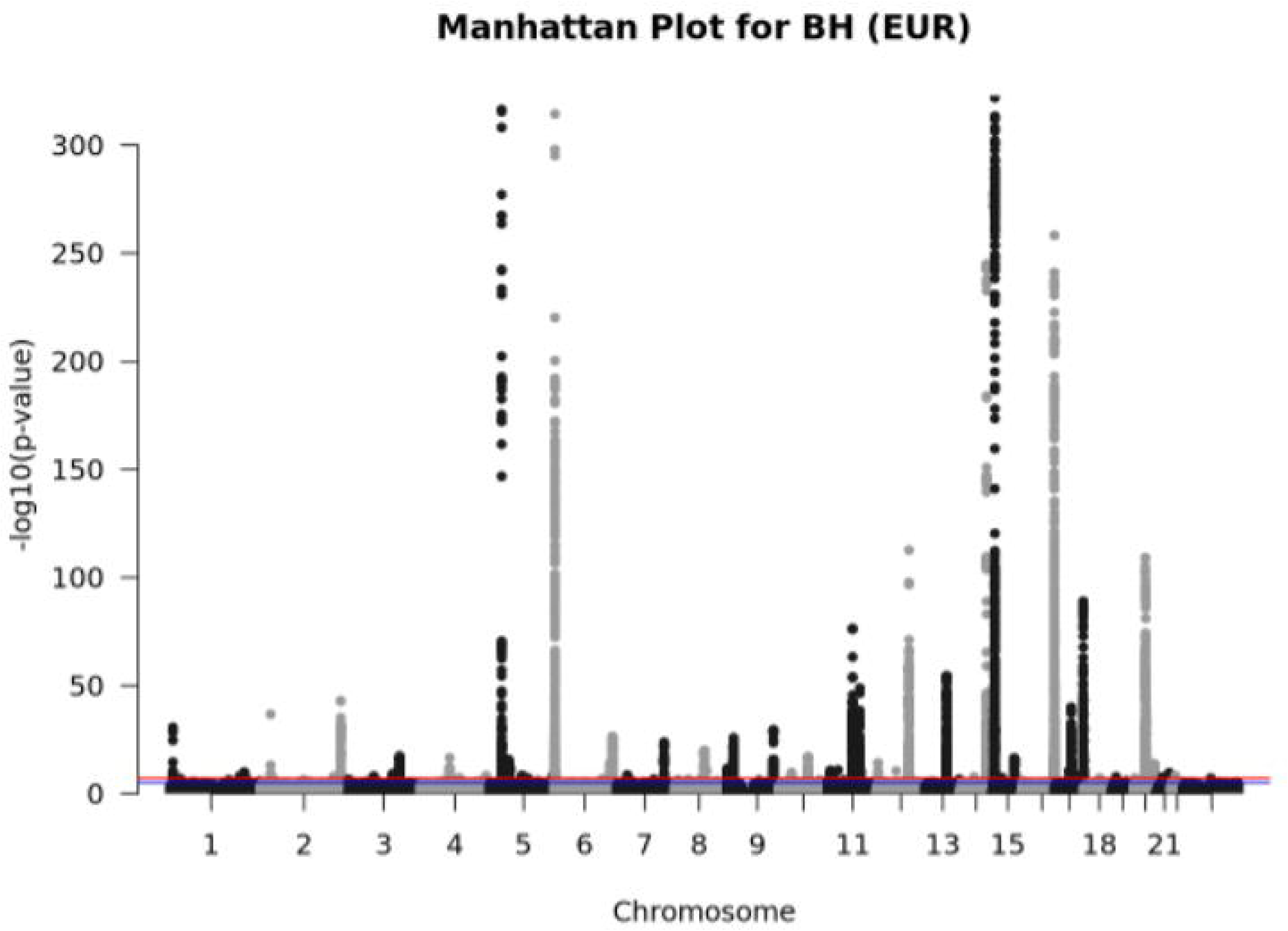

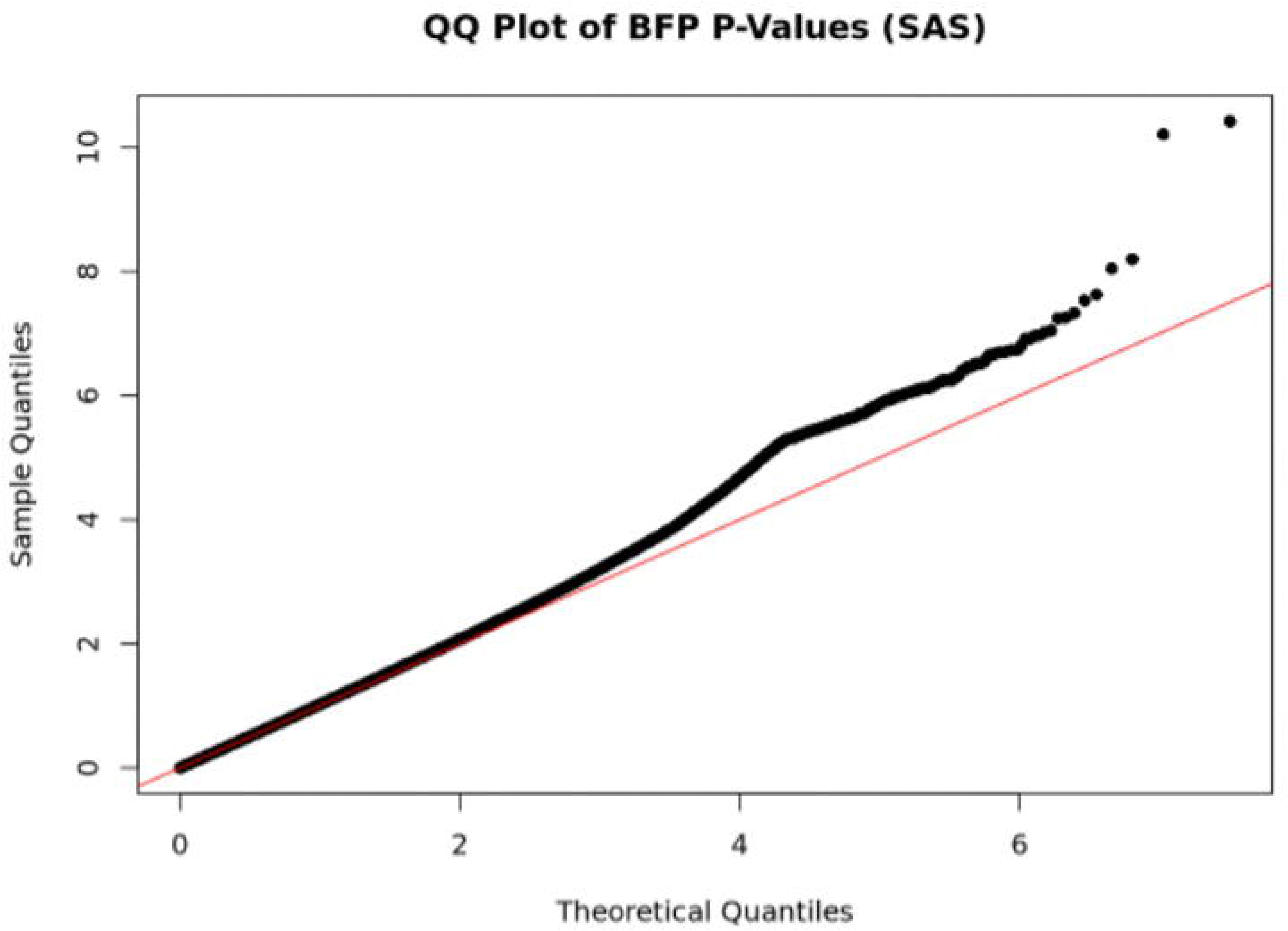

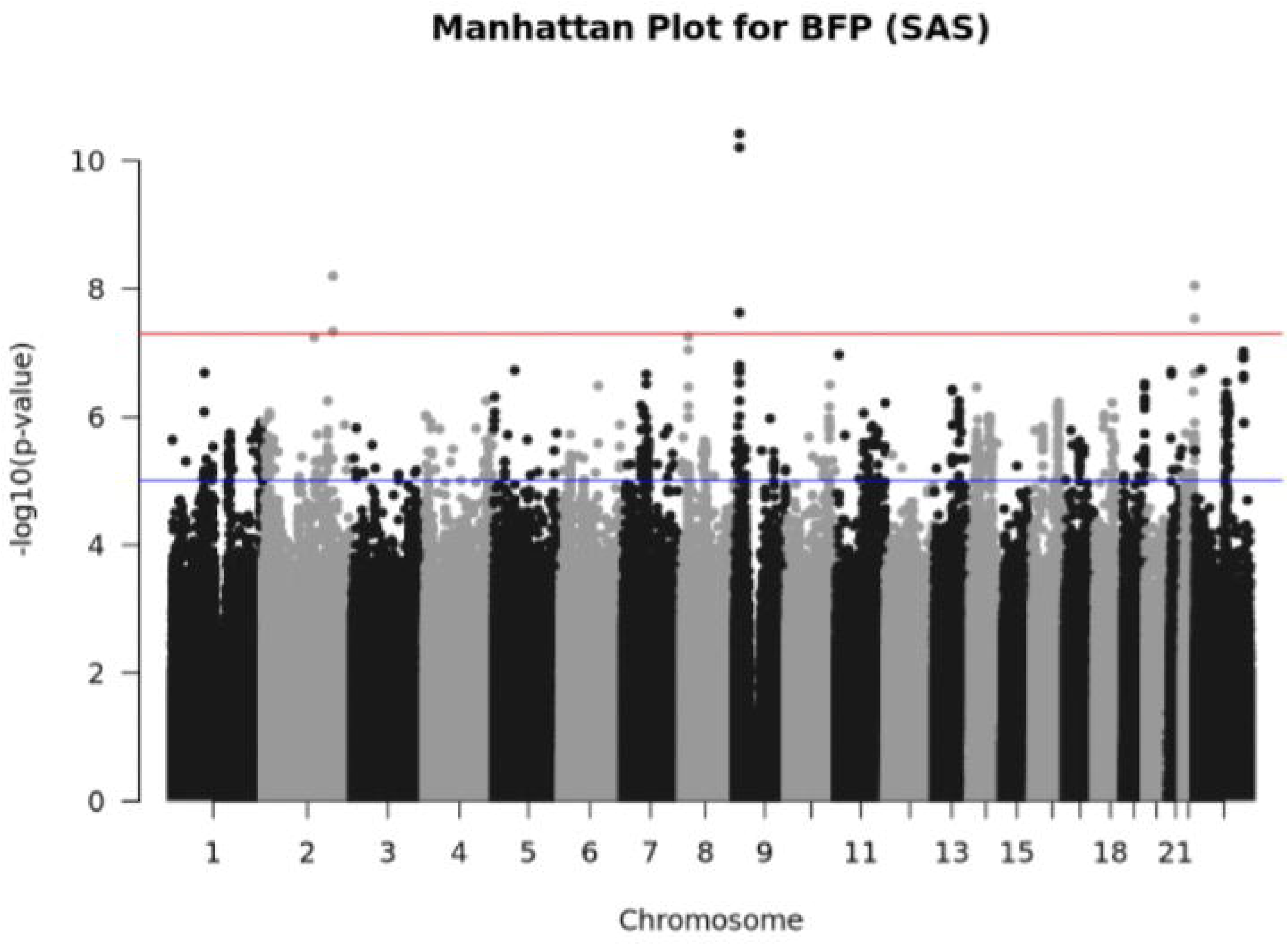

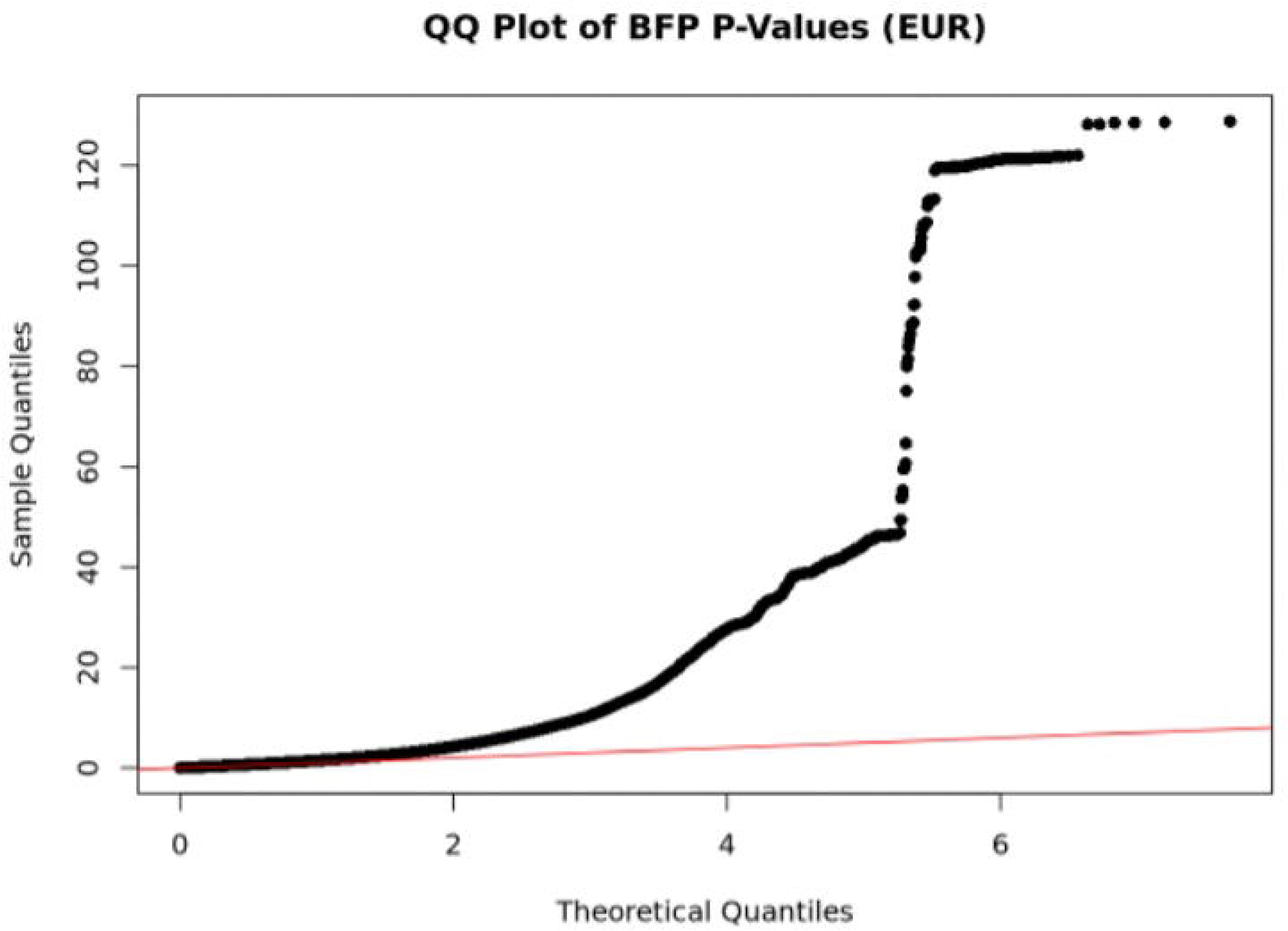

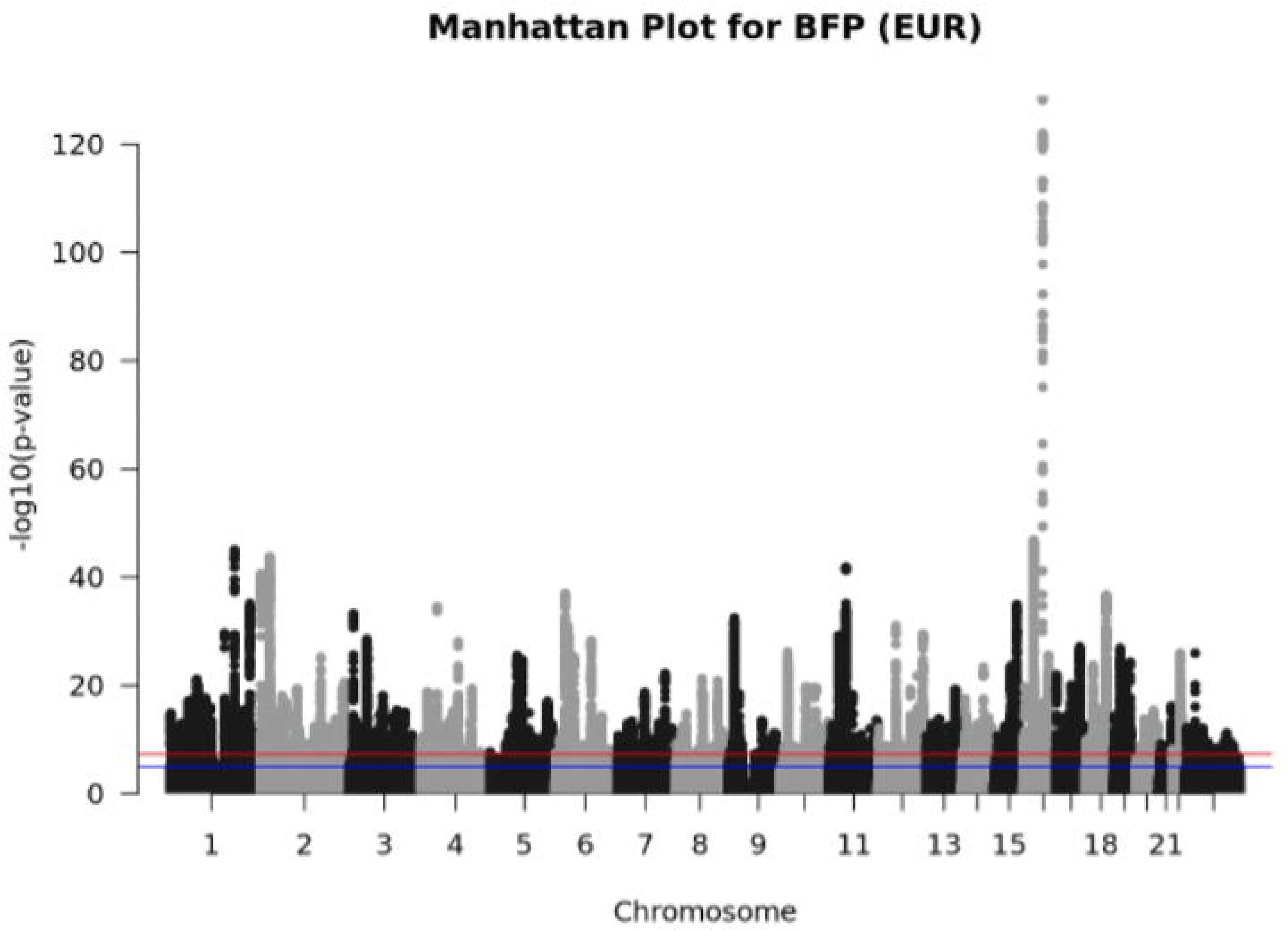

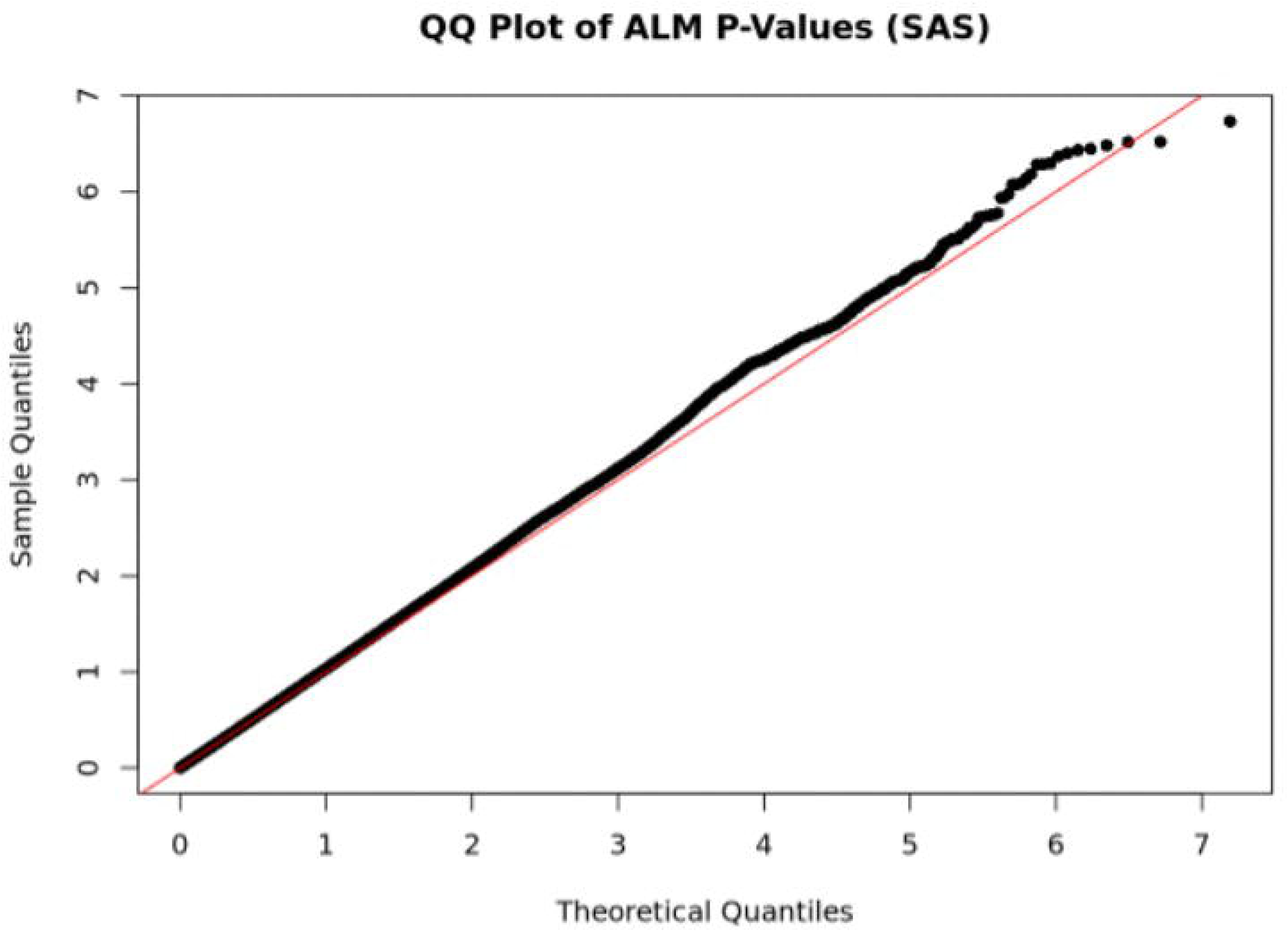

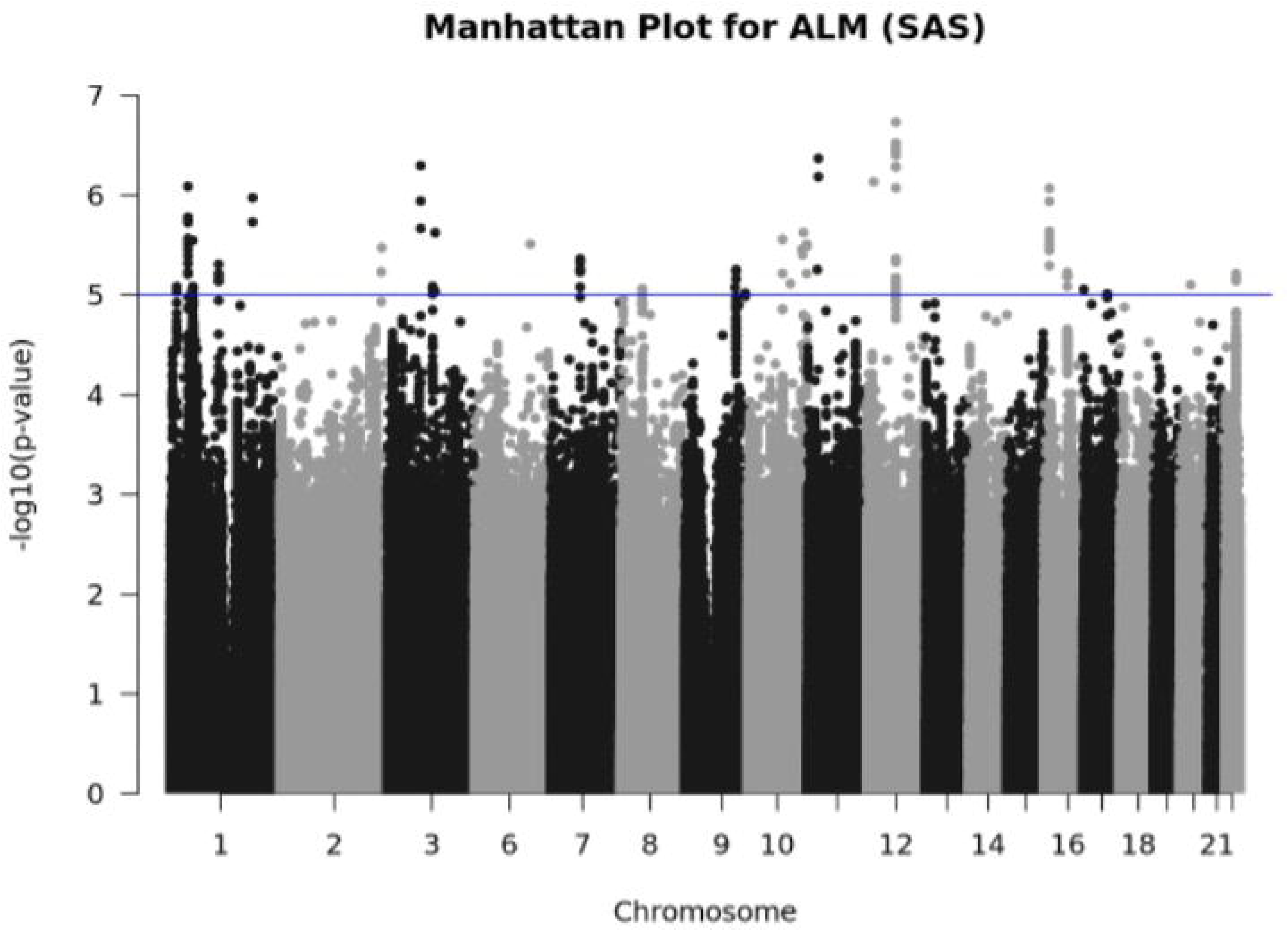

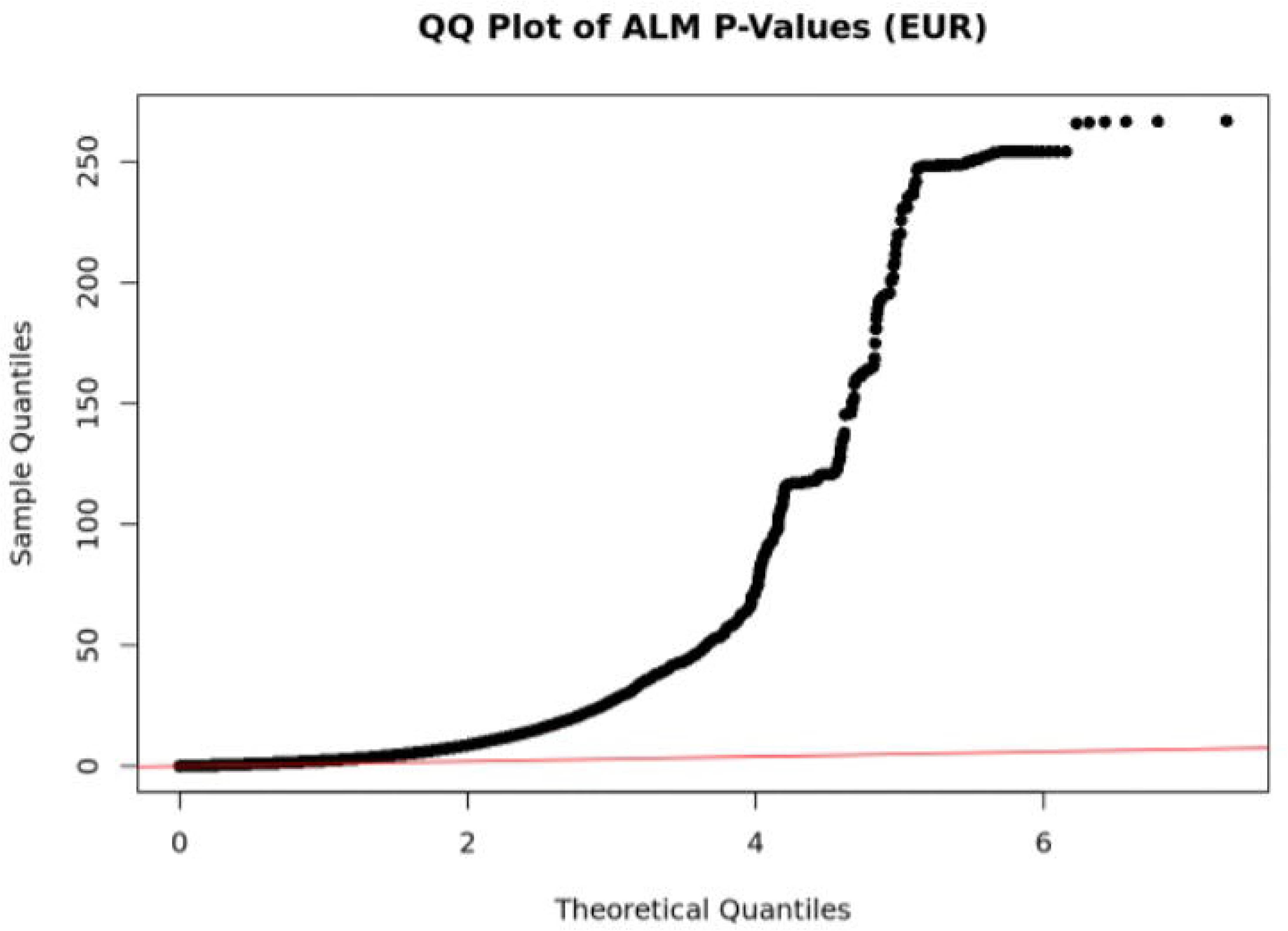

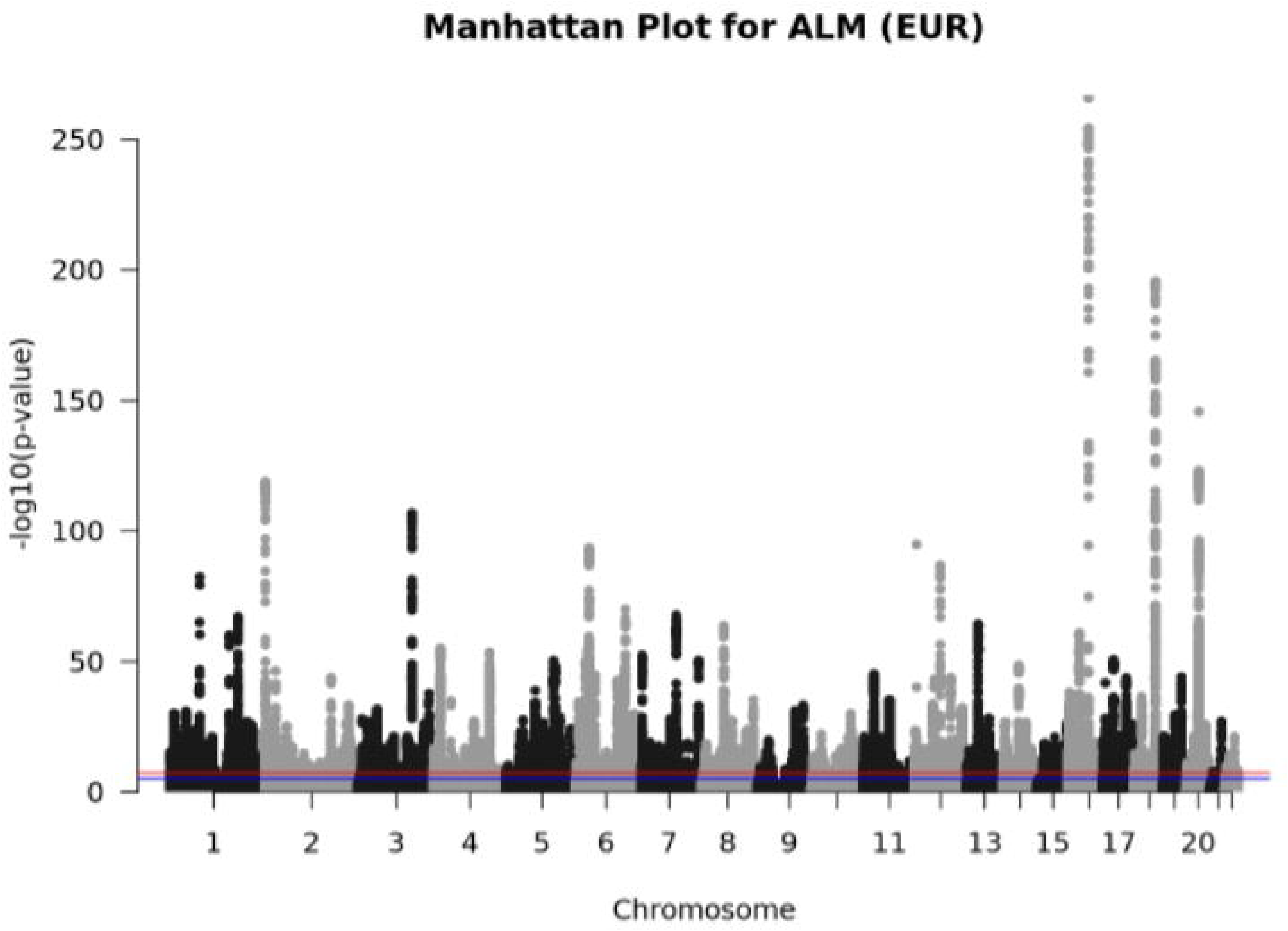

